# Neutralizing activity of Sputnik V vaccine sera against SARS-CoV-2 variants

**DOI:** 10.1101/2021.03.31.21254660

**Authors:** Satoshi Ikegame, Mohammed N. A. Siddiquey, Chuan-Tien Hung, Griffin Haas, Luca Brambilla, Kasopefoluwa Y. Oguntuyo, Shreyas Kowdle, Ariel Esteban Vilardo, Alexis Edelstein, Claudia Perandones, Jeremy P. Kamil, Benhur Lee

**Author notes:** Correspondence to: Benhur Lee. These authors contributed equally to the study (Satoshi Ikegame, Mohammed N. A. Siddiquey). Senior authors (Claudia Perandones, Jeremy P. Kamil, Benhur Lee). **Competing interests:** B.L. and K.Y.O. are named inventors on a patent filed by the Icahn School of Medicine for some of the materials used in this work. J.P.K. is a consultant for BioNTech (advisory panel on coronavirus variants).

## Abstract

The novel pandemic betacoronavirus, severe acute respiratory syndrome coronavirus 2 (SARS-CoV-2), has infected at least 120 million people since its identification as the cause of a December 2019 viral pneumonia outbreak in Wuhan, China. Despite the unprecedented pace of vaccine development, with six vaccines already in use worldwide, the emergence of SARS-CoV-2 ‘variants of concern’ (VOC) across diverse geographic locales suggests herd immunity may fail to eliminate the virus. All three officially designated VOC carry Spike (S) polymorphisms thought to enable escape from neutralizing antibodies elicited during initial waves of the pandemic. Here, we characterize the biological consequences of the ensemble of S mutations present in VOC lineages B.1.1.7 (501Y.V1) and B.1.351 (501Y.V2). Using a replication-competent EGFP-reporter vesicular stomatitis virus (VSV) system, rcVSV-CoV2-S, which encodes S from SARS coronavirus 2 in place of VSV-G, and coupled with a clonal HEK-293T ACE2 TMPRSS2 cell line optimized for highly efficient S-mediated infection, we determined that only 1 out of 12 serum samples from a cohort of recipients of the Gamaleya Sputnik V Ad26 / Ad5 vaccine showed effective neutralization (IC_90_) of rcVSV-CoV2-S: B.1.351 at full serum strength. The same set of sera efficiently neutralized S from B.1.1.7 and showed only moderately reduced activity against S carrying the E484K substitution alone. Taken together, our data suggest that control of some emergent SARS-CoV-2 variants may benefit from updated vaccines.

## INTRODUCTION

In the 15 months since its emergence in late 2019 ^1^, SARS-CoV-2 has caused over 131 million confirmed COVID-19 cases worldwide, leading to at least 2.85 million deaths ^2^. SARS-CoV-2 is closely related to two other zoonotic betacoronaviruses, MERS-CoV and SARS-CoV, that also cause life-threatening respiratory infections ^3^.

This global health emergency has spurred the development of COVID-19 preventive vaccines at an unprecedented pace. Six are already authorized for human use across the globe ^4–9^. These vaccines focus on the SARS-CoV-2 spike protein (S), due to its critical roles in cell entry. Indeed, the presence of serum neutralizing antibodies directed at S correlate strongly with protection against COVID-19 ^10,11^. Although these six vaccines are efficacious, the recent emergence of novel SARS-CoV-2 variants has reignited concerns that the pandemic may not be so easily brought under control.

In December 2020, the United Kingdom reported the sudden emergence of a novel SARS-CoV-2 lineage, termed B.1.1.7 (501Y.V1, VOC 202012/01), which was designated as the first SARS-CoV-2 variant of concern (VOC). The lineage had rapidly increased in prevalence since first being detected in November 2020 ^12^. Its genome showed an unusually high number of non-synonymous substitutions and deletions, including eight in the S gene, suggesting a substantial degree of host adaptation that may have occurred during prolonged infection of an immunocompromised person ^13^. The B.1.1.7 lineage has now been shown to exhibit enhanced transmissibility ^14^ as well as an increased case fatality rate ^15,16^.

Soon afterwards, two additional SARS-CoV-2 VOC, B.1.351 and P.1, were reported from S. Africa and Brazil, respectively, which each showed substantial escape from neutralizing antibodies elicited by first wave pandemic viruses, leading to documented cases of re-infection ^17–19^. The S genes of B.1.351 and P.1 viruses each carry a number of mutations, but include three in the receptor binding domain (RBD) that are particularly notable, the S: N501Y substitution, found in B.1.1.7, alongside polymorphisms at positions 417 and 484, K417N/T and E484K. S: E484K had already been identified in multiple independent laboratories to confer escape from convalescent sera and monoclonal antibodies ^20–22^. As expected, the P.1 and B.1.351 variants escape or resist neutralization by first wave convalescent sera, as well as antibodies elicited by COVID-19 vaccines ^23–27^.

Although the P.1 and B.1.351 lineages are dominant in Brazil and S. Africa, unlike B.1.1.7 they have not increased greatly in number in the United States since originally being detected here. In contrast, the E484K polymorphism is recurrently emergent, and is found in a number of other lineages that are increasing in the U.S. and other countries. For example, a B.1.526 sub-lineage carrying E484K in recent weeks has expanded more rapidly than B.1.1.7 ^28,29^, which may be indicative of the ability of S: E484K variants to penetrate herd immunity. The P.2 lineage, originally detected in Rio de Janeiro, carries only the E484K mutation in the RBD and has spread to other parts of South America, including Argentina ^30^.

The six COVID-19 vaccines currently in use around the world employ different strategies, and do not all incorporate the two proline substitutions that “lock” S into the pre-fusion conformer. Vaccines that do not utilize pre-fusion “locked” S are expected to produce lower levels of neutralizing antibodies, and hence may be less efficacious against infection, even if they do protect against severe COVID-19^31,32^. Indeed, a two-dose regimen of the AstraZeneca ChAdOx1 based vaccine, which does not use a “locked” S, did not protect against mild-to-moderate COVID-19 in S. Africa, where 93% of COVID-19 cases in trial participants were caused by the B.1.351 variant ^33^. Like the AstraZeneca ChAdOx1 vaccine, the Sputnik V vaccine (Gam-COVID-Vac) is based on adenovirus vectored expression of a native S sequence, rather than a pre-fusion “locked” S ^34^. Although the Sputnik V vaccine has a reported vaccine efficacy of 91.6% in the interim analysis of Phase 3 trials held in Russia between Sept 7 and Nov 24, 2020, none of the VOC mentioned above nor independent lineages containing the E484K mutation were prevalent in Russia during this time period. Since the Sputnik vaccine is now in use not only in Russia, but also in countries like Argentina, Mexico, and Hungary, where some of the VOC and emerging lineages bearing the E484K mutation are more widespread, it is critical to assess the neutralizing activity of Sputnik vaccine elicited antibody responses against these cognate VOC and mutant spikes.

This study characterizes the neutralization activity of sera from a dozen Sputnik V vaccine recipients in Argentina. Our work was spurred by Argentina’s nascent genomic surveillance efforts, which detected multiple independent lineages with S: E484K (B.1.1.318 and P.2) and/or S: N501Y substitutions (B.1.1.7 and P.1) in common, just as Argentina had started rolling out its vaccination campaign, which commenced on Dec 29, 2020. Here, we generated isogenic replication-competent vesicular stomatitis virus bearing the prevailing wild-type (WT=D614G) SARS-CoV-2 S (rcVSV-CoV2-S), or the B.1.1.7, B.1.351 or E484K mutant S and used them in a robust virus neutralization assay. Our results show that Sputnik V vaccine sera effectively neutralized S: WT and S: B.1.1.7. viruses, albeit with highly variable titers. The same sera, however, exhibited moderate and markedly reduced neutralization titers, respectively, against S: E484K and S: B.1.351. Analyses of dose response curves indicate that S: B.1.351 exhibits resistance to neutralizing sera in a manner that is qualitatively different from the E484K mutant. Taken together, our data argue that surveillance of the neutralizing activity elicited by vaccine sera will be necessary on an ongoing basis. Viral neutralization assays can indicate which SARS-CoV-2 variants are likely capable of transmission in the face of vaccine elicited immunity, and whether updated vaccines will be needed to control their emergence and spread.

## RESULTS

### Robust reverse genetics for generating replication-competent VSV expressing SARS-CoV-2 Spike proteins

Several groups have now generated replication-competent VSV expressing SARS-CoV-2 spike in place of VSV-G (rcVSV-CoV2-S)^35–37,38^. These rcVSV-CoV2-S can be used in BSL-2 compatible virus neutralization assays (VNAs), which correlate very well with VNAs using live SARS-CoV-2 (Spearman’s r > 0.9 across multiple studies). rcVSV-CoV2-S has been assessed as a candidate vaccine ^37,39^, and used in forward genetics experiments to generate antibody escape mutants or perform comprehensive epitope mapping studies ^40,20,38^. Indeed, the now concerning E484K mutation, present in many variants of concern (VOC), was identified as an antibody escape mutation using rcVSV-CoV-2-S ^20,38^.

However, many groups passage their rcVSV-CoV-2-S extensively in Vero cells after the initial rescue, either to generate higher titer stocks and/or to remove confounding components such as the vaccinia virus expressing T7-polymerase and/or transfected VSV-G, both of which were deemed necessary for efficient rescue ^38^. Serial passage of rcVSV-CoV-2-S in Vero cells invariably leads to mutations in the S1/S2 furin cleavage site, as well as truncations in the cytoplasmic tail of the S protein ^39^. The latter promotes S incorporation into VSV without compromising the conformational integrity of the ectodomain, whereas the former is problematic when assessing the neutralization sensitivity and structure-function phenotype of Spike VOC with multiple mutations that likely have complex epistatic interactions.

To generate rcVSV-CoV2-S containing different variants or mutants on demand, without the need for extensive passaging, we developed a robust reverse genetics system and VNA which leverages the cell lines we previously developed for a standardized SARS-CoV-2 VNA that correlates well with live virus neutralization ^41^. Salient improvements include the addition of a hammerhead ribozyme immediately upstream of the 3’ leader sequence which cleaves *in cis* to give the exact 3’ termini, the use of a codon-optimized T7-polymerase which alleviates the use of vaccinia-driven T7-polymerase, and a highly permissive and transfectable 293T-ACE2+TMPRSS2 clone (F8-2) ^41^ (Extended Data Fig S1). A 6-plasmid transfection into F8-2 cells results in GFP+ cells 2-3 days post-transfection (dpt), which turn into foci of syncytia by 4-5 dpt indicating virus replication and cell-to-cell spread (Fig. 1A). Transfer of F8-2 cell supernatant into interferon-defective Vero-TMPRSS2 cells allowed for rapid expansion of low-passage viral stocks that maintain only the engineered Spike mutations. Clarified viral supernatants from Vero-TMPRSS2 cells were aliquoted, sequenced verified, then titered on F8-2 cells to determine the linear range of response (Fig. 1B).

**Figure 1.**
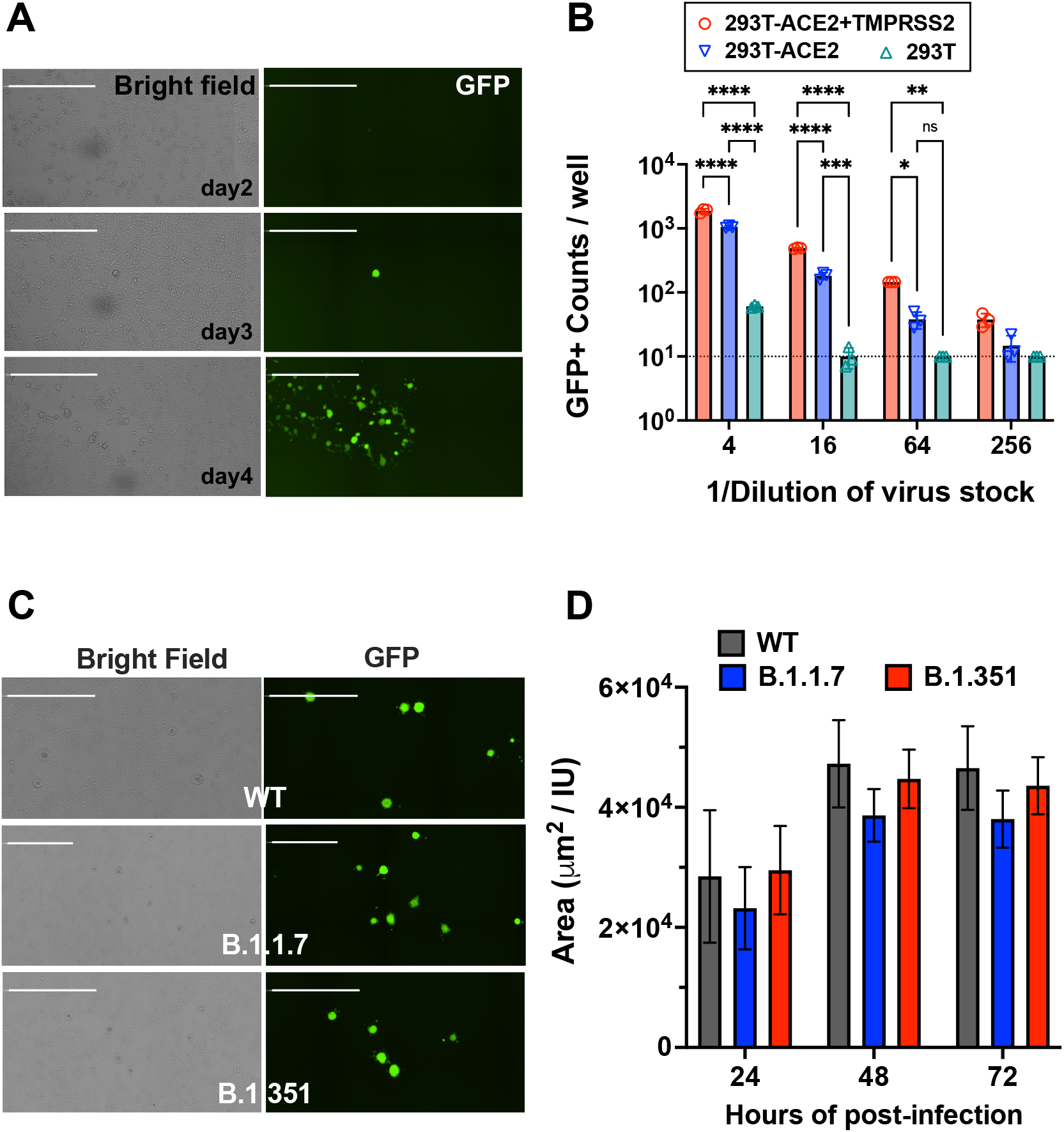
Replication-competent VSV bearing wild-type and variant SARS-CoV-2 spike (rcVSV-CoV2-S). **(A)** Representative images of *de novo* generation of rcVSV-CoV2-S, carrying an EGFP reporter, in transfected 293T-ACE2+TMPRSS2 (F8-2) cells as described in Extended Data Fig. S1. Single GFP+ cells detected at 2-3 days post-transfection (dpt) form a foci of syncytia by 4 dpt. Images are taken by Celigo imaging cytometer (Nexcelom) and are computational composites from the identical number of fields in each well. White bar is equal to 1 millimeter. **(B)** Entry efficiency of rcVSV-CoV2-S in parental 293T cells, 293T stably expressing ACE2 alone (293T-ACE2) or with TMPRSS2 (293T-ACE2+TMPRSS2). Serial dilutions of virus stocks amplified on Vero-TMPRSS2 cells were used to infect the indicated cell lines in 96-well plates in triplicates. GFP signal was detected and counted by a Celigo imaging cytometer (Nexcelom) 10 hours post-infection. Symbols are individual data points from triplicate infections at the indicated dilutions. Bars represent the average of 3 replicates with error bars indicating standard deviation. A two-way ANOVA was used to compare the differences between cell lines at any given dilution. Adjusted p values from Tukey’s multiple comparisons test are given (ns; not significant, * p < 0.05, ** p < 0.01, *** p < 0.001, **** p < 0.0001). **(C)** rcVSV-CoV-2-S containing the prevailing WT (D614G) and VOC (B.1.1.7 and B.1.351) spikes were inoculated into one 6-well each of F8-2 cells (MOI 0.1) and subsequently overlaid with methylcellulose-DMEM to monitor syncytia formation. Representative images of syncytial plaques at 48 hpi are shown. White bar equals 1 millimeter. **(D)** shows the growth of GFP positive area / infectious unit (IU) in the 6 well plate. GFP positive areas were imaged and measured by the Celigo imaging cytometer. IU was checked at 10 hpi in the same well. Bar shows the average of 3 independent experiments with error bar indicating standard deviation. No statistically significant differences were detected between WT and VOC spikes in the size of GFP+ syncytia at any given time point (two-way ANOVA as above, ‘ns’ not indicated in graph).

Next, we generated isogenic rcVSV-CoV2-S expressing the B.1.1.7, B.1.351 (Fig. 2A), or E484K S to evaluate the neutralizing activity of Sputnik V vaccine sera from Argentina. The relevant Spike substitutions that make up these variants are indicated in Fig. 2A. The characteristics of the vaccine recipient cohort (n=12) receiving the two-dose regimen of the Sputnik vaccine are given in Table 1. At one month post-completion of the two-dose regimen, the Sputnik V vaccine generated respectable virus neutralizing titers (VNT) against rcVSV-CoV2-S bearing the WT (D614G) and B.1.1.7 spike proteins (Fig. 2B). The geometric mean titer (GMT) and 95% CI for WT (1/IC_50_ GMT 49.4, 23.4 - 105) in our cohort of vaccine recipients was remarkably similar to that reported in the phase III Sputnik vaccine trial (GMT 44.5, 31.8 - 62.2)^48,49^. However, GMT against B.1.351 and E484K was reduced by a median 6.8- and 2.8-fold, respectively compared to WT (Fig. 2C).

**Table 1.** Cohort characteristics of Sputnik vaccine recipients from ANLIS MALBRÁN (Buenos Aires, República Argentina).

| Sera ID | 1<sup>st</sup> DOSE | 2<sup>nd</sup> DOSE | Vaccine Status | SEX | AGE |
| --- | --- | --- | --- | --- | --- |
| SP001 | Late Dec/2020 | Mid Jan/2021 | (+) | M | 45-50 |
| SP002 | Late Dec/2020 | Mid Jan/2021 | (+) | M | 40-45 |
| SP003 | Late Dec/2020 | Mid Jan/2021 | (+) | M | 55-60 |
| SP004 | Late Dec/2020 | Mid Jan/2021 | (+) | M | 50-55 |
| SP005 | Late Dec/2020 | Mid Jan/2021 | (+) | M | 35-40 |
| SP006 | Late Dec/2020 | Mid Jan/2021 | (+) | F | 35-40 |
| SP007 | Late Dec/2020 | Mid Jan/2021 | (+) | F | 20-25 |
| SP008 | Late Dec/2020 | Early Feb/2021 | (+) | M | 35-40 |
| SP009 | Late Dec/2020 | Early Feb/2021 | (+) | F | 30-35 |
| SP010 | Late Dec/2020 | Mid Jan/2021 | (+) | M | 30-35 |
| SP011 | Late Dec/2020 | Mid Jan/2021 | (+) | M | 40-45 |
| SP012 | Late Dec/2020 | Mid Jan/2021 | (+) | M | 25-30 |
|  |  |  |  | **Median Age** | **39.5** |
|  |  |  |  | **Range** | **25-56** |
| SP013 | N.A. | N.A. | (-) | F | 45-50 |
| SP014 | N.A. | N.A. | (-) | F | 50-55 |
| SP015 | N.A. | N.A. | (-) | M | 40-45 |

**Figure 2.**
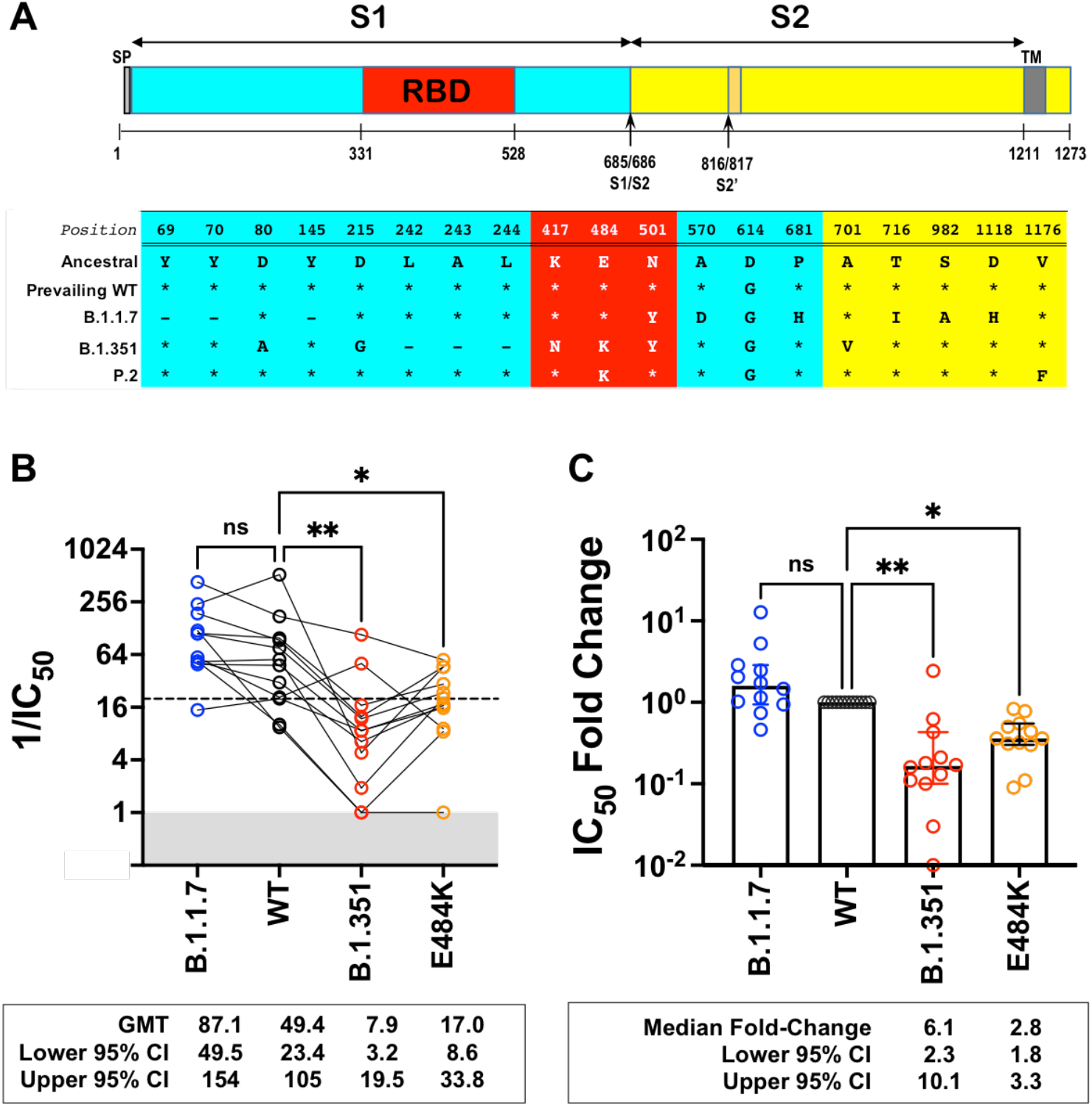
Neutralization activity of antibody responses elicited by the Sputnik V vaccine. **(A)** Schematic of the Spike substitutions that make up the variants being evaluated in this study. The amino acid positions and corresponding ‘Ancestral’ sequence of the Wuhan isolate is shown. The prevailing WT sequence now has a D614G substitution. All the variants and mutants have D614G. **(B)** Neutralization activity of individual serum samples against rcVSV-CoV2-S with the WT, variant (B.1.1.7 or B.1.351), or mutant E484K spike proteins. Neutralization is represented by the reciprocal 50% inhibitory dilution factor (1/IC_50_). Sera samples with no appreciable neutralization against a given virus were assigned a defined 1/IC_50_ value of 1.0, as values ≤1 are not physiological (Grey shaded area). Dashed line indicates the lowest serum dilution tested (1/IC50 = 20). Geometric mean titers (GMT and 95% CI) for the neutralizing activity of all vaccine sera are indicated below each of the viral spike proteins examined. NS; not significant, *; p<0.05, p < 0.01; ** are adjusted p values from non-parametric one-way ANOVA with Dunn’s multiple comparisons test. **(C)** For each serum sample, the fold-change in IC_50_ (reciprocal inhibitory dilution factor) against the indicated variant and mutant spike proteins relative to its IC_50_ against wild-type (WT) spike (set at 1) is plotted. Adjusted p values were calculated as in (B). Medians are represented by the bars and whiskers demarcate the 95% CI. Neutralization dose-response curves were performed in triplicates, and the mean values from each triplicate experiment are shown as the single data points for each sera sample.

Sputnik vaccine recipients appeared to generate qualitatively different neutralizing antibody responses against SARS-CoV-2 that could be segregated into three different groups (Fig. 3). Group (A) sera showed reasonable VNT against wild-type (WT) and B.1.1.7 (Fig. 3A and E). However, the Hill slope of their neutralization curves for B.1.351 were extremely shallow (h<0.40), resulting in a low IC50s and maximal neutralization of 50-60% even when extrapolated to full serum strength (Fig. 3E and Fig. 4). In contrast, Group (B) sera neutralized E484K and B.351 with similar potencies to WT and B.1.1.7, especially at high serum concentrations (Fig. 3B and E). This group of sera reveals that qualitatively different neutralizing responses can be generated that effectively neutralize B.1.351. Group (C) sera generally exhibited effective neutralization of WT, B.1.1.7, and even E484K at high serum concentrations, but not B.1.351 (Fig. 3C and E). The decreased potency and shallow Hill Slope result in <90% neutralization of B.1.351 even at full serum strength (see next section). One serum sample (SP012) exhibited little to no neutralizing activity against WT, E484K and B.1.351, yet it neutralized B.1.1.7 as well as Group A-C sera (Fig. 3D-E). That these three groups exhibit qualitatively distinct neutralization patterns is further highlighted by the Hill slopes of their neutralization curves (Fig. 3F). Group A sera not only have the lowest slopes against B.1.351, but as a group, they have slopes significantly <1 against the other viruses (median/IQR = 0.4965 / 0.2880 – 1.186). Group B sera mostly have slope values around 1 (median/IQR = 0.8855 / 0.7865 – 1.065) while Group C sera have the highest overall slopes (median/IQR = 1.348 / 0.8395 – 1.820) that are significantly >1 (Fig. 3F).

**Figure 3.**
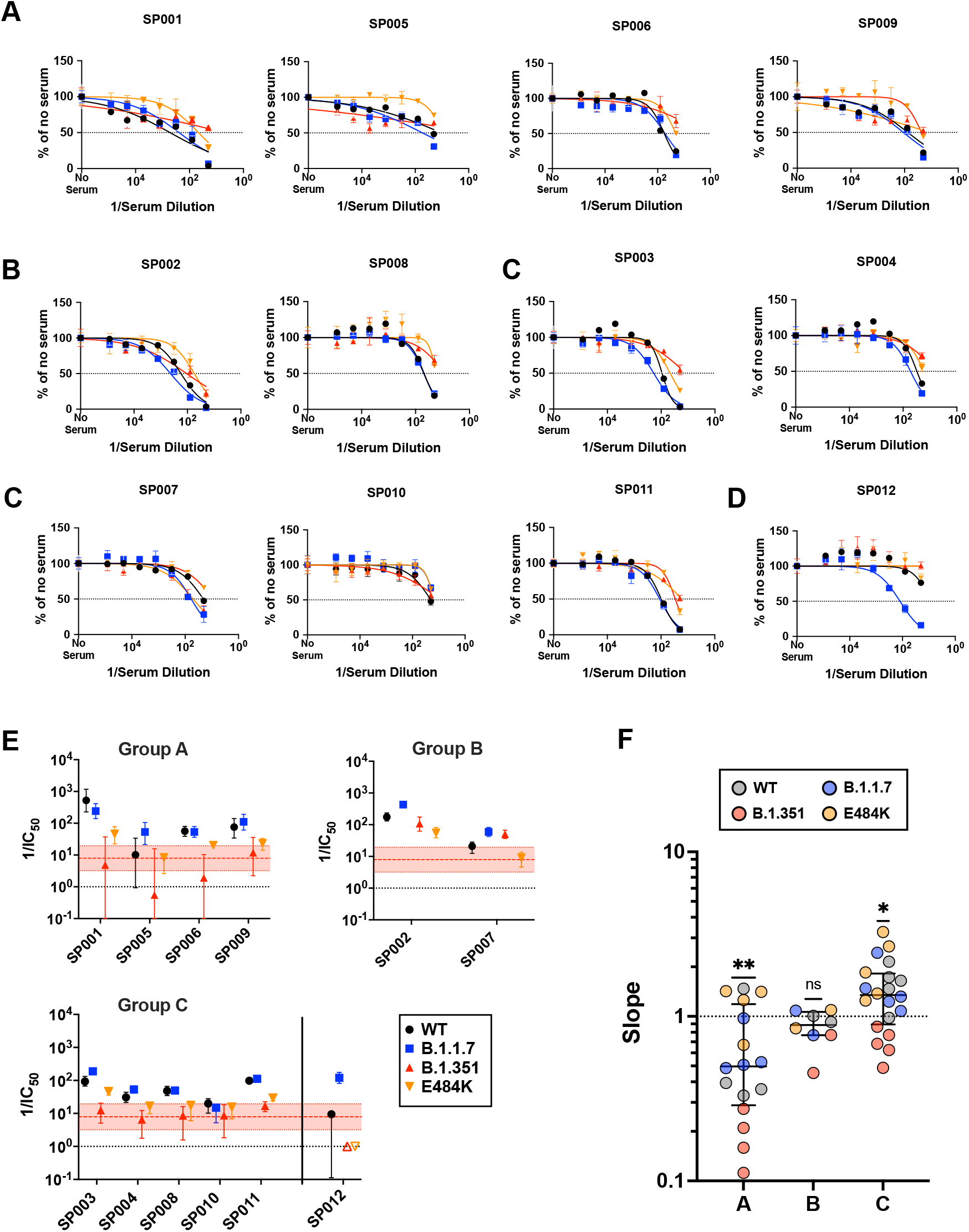
Sputnik vaccine recipients generate qualitatively different neutralizing antibody responses against SARS-CoV-2. **(A-C)** Group A (SP001, SP005, SP006, SP012), Group B (SP002, SP007), and Group C (SP003, SP004, SP008, SP010, SP011) represent potentially distinct classes of virus neutralizing activity present in the sera samples analyzed. Full neutralization curves for all sera tested against all viruses bearing the variant and mutant spike proteins are shown. **(D)** shows a singular example of a serum that only neutralized the B.1.1.7 spike. **(E)** graphs the serum neutralizing titers (SNT = 1/IC_50_) and 95% CI that can be extrapolated from the nonlinear regression curves shown for all the sera samples analyzed. Colored filled symbols represent the indicated viruses, open symbols in (E) represent assigned SNT values of 1.0 when no significant neutralization activity could be detected (SP012, B.1.351 and E484K). The dotted black line represents a reciprocal serum dilution of 1.0. The red dashed line and shaded boundaries represent the geometric mean titer and 95% CI, respectively, for B.1.351. **(F)** The Hill slope values for all the neutralization curves are aggregated according to their groups. The different colored symbols in each group represent the indicated virus tested. P values are from a non-parametric Wilcoxon signed rank test using a theoretical median of 1.0.

**Figure 4.**
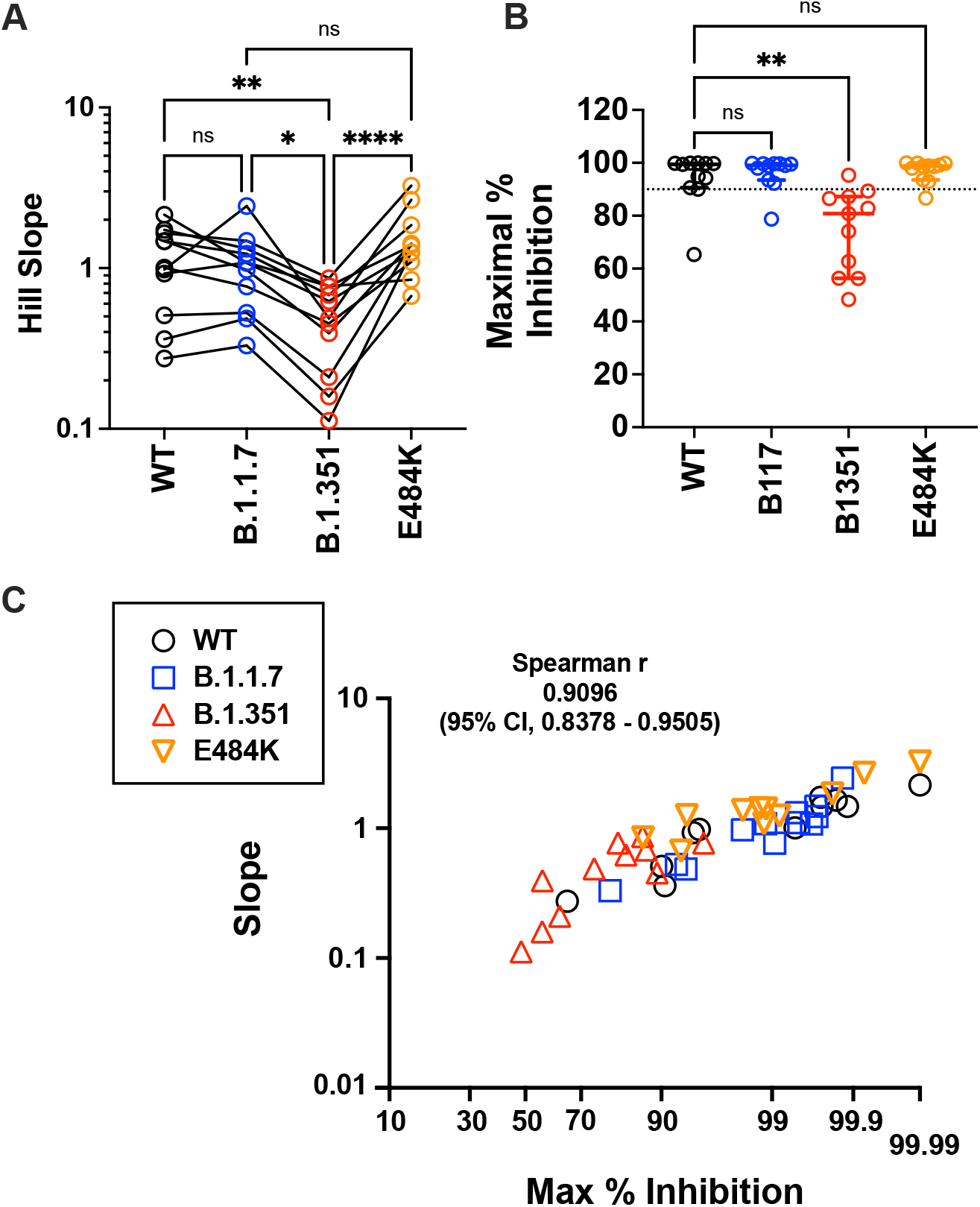
Maximal inhibition and slope help to define the distinct classes of neutralizing sera in Sputnik vaccine recipients. **(A)** Paired comparison of Hillslopes from the neutralization curves of all samples except for SP012 where no significant neutralization was observed for viruses other than B.1.1.7. NS; not significant, p<0.05, *; p<0.01; **, p < 0.0001; ****, are adjusted p values from non-parametric one-way ANOVA with Dunn’s multiple comparisons test, which assumes non-Gaussian distribution of values being analyzed. **(B)** Maximal percent inhibition (MPI) at full serum strength extrapolated from nonlinear regression of log(inhibitor) versus normalized response, variable slope curve. Model used is from PRISM v9.1 where Y= 100/(1+10^((LogIC50-X)*HillSlope))). Log IC50 and Hill slope values were obtained for each curve generated in Fig. 3. MPI = 100-Y, when X= 0 for reciprocal serum dilution of 1 (10^0 =1). Data points for one serum (SP012) against WT, B.1.351 and E484K could not be calculated because there was no best-fit value. The dotted line indicates 90% inhibition. Median (central bar) and interquartile values (whiskers) are indicated. Adjusted p values was calculated as in (A). **(C)** Correlation analysis of MPI versus the Hill Slope parameter for all sera samples tested against all spike proteins. SP012 was excluded for the abovementioned reasons. Non-parametric Spearman r values and 95% confidence interval are shown. X-axis is plotted as an asymptotic cumulative probability scale as x approaches 100% (PRISM v9.1.1) only to resolve the many MPI values >90%.

The Hill Slope of the neutralization curves against B.1.351 was significantly different from WT, B.1.1.7 and E484K (Fig. 4A). As a consequence, the maximal neutralization attainable when extrapolated to full serum strength was also significantly lower for B.1.351 compared to the rest (Fig. 4B). Conversely, the steep Hill slope for the E484K curves resulted in maximal neutralization potencies that were not significantly different from WT or B.1.1.7 despite significantly lower reciprocal IC_50_ values (compare Fig. 2B with Fig. 4B). Notably, the maximal percent inhibition was strongly correlated with the Hill Slope for WT and VOC/mutant spikes across all valid pairs of sample values (Fig. 4C, n=45 pairs), suggesting that antibody co-operativity likely plays a role at high serum concentrations (see Discussion) ^42^. While we acknowledge the limitations of extrapolating values from nonlinear regression curves, the striking correlation between slope and maximal percent inhibition attainable at full serum strength supports the robustness of our nonlinear regression model.

The heterogenous dose-response curves described in Fig. 3-4 is a property of Sputnik V vaccine elicited responses as soluble RBD-Fc inhibition of WT and VOC S-mediated entry produced classical dose response curves with Hill slopes close to -1.0 (Fig. 5). Both B.1.1.7 and B.1.351 were modestly but significantly more resistant to RBD-Fc inhibition (Fig. 5A-B). This is not surprising as both harbor the N501Y mutation known to enhance affinity of RBD for ACE2 ^43–45^. However, this 1.5 to 2-fold increase in RBD-Fc IC_50_ for B.1.1.7 and B.1.351, respectively, does not explain the neutralization-resistant versus sensitive phenotype of B.1.351 versus B.1.1.7 in our virus neutralization assays. Furthermore, the E484K mutant was more sensitive to RBD-Fc inhibition than B.1.1.7 (Fig. 5C-D), and yet also remained more neutralization-resistant relative to B.1.1.7. Experimental measurements of both RBD and trimeric spike binding to ACE2 have revealed that the E484K mutation alone does not confer increase binding affinity for ACE2 unlike N501Y ^44,46^. Our RBD-Fc inhibition studies in the context of virus infection confirm and extend these results. Our data reinforces the notion that the mechanism underlying the increased neutralization resistance of E484K containing variants and mutants do not involve ACE2 binding affinity per se, but rather affects a key immunodominant epitope targeted by a significant class of human neutralizing antibodies, variably termed as RBM class II, RBS-B, or Cluster 2 antibodies ^47–50^.

**Figure 5.**
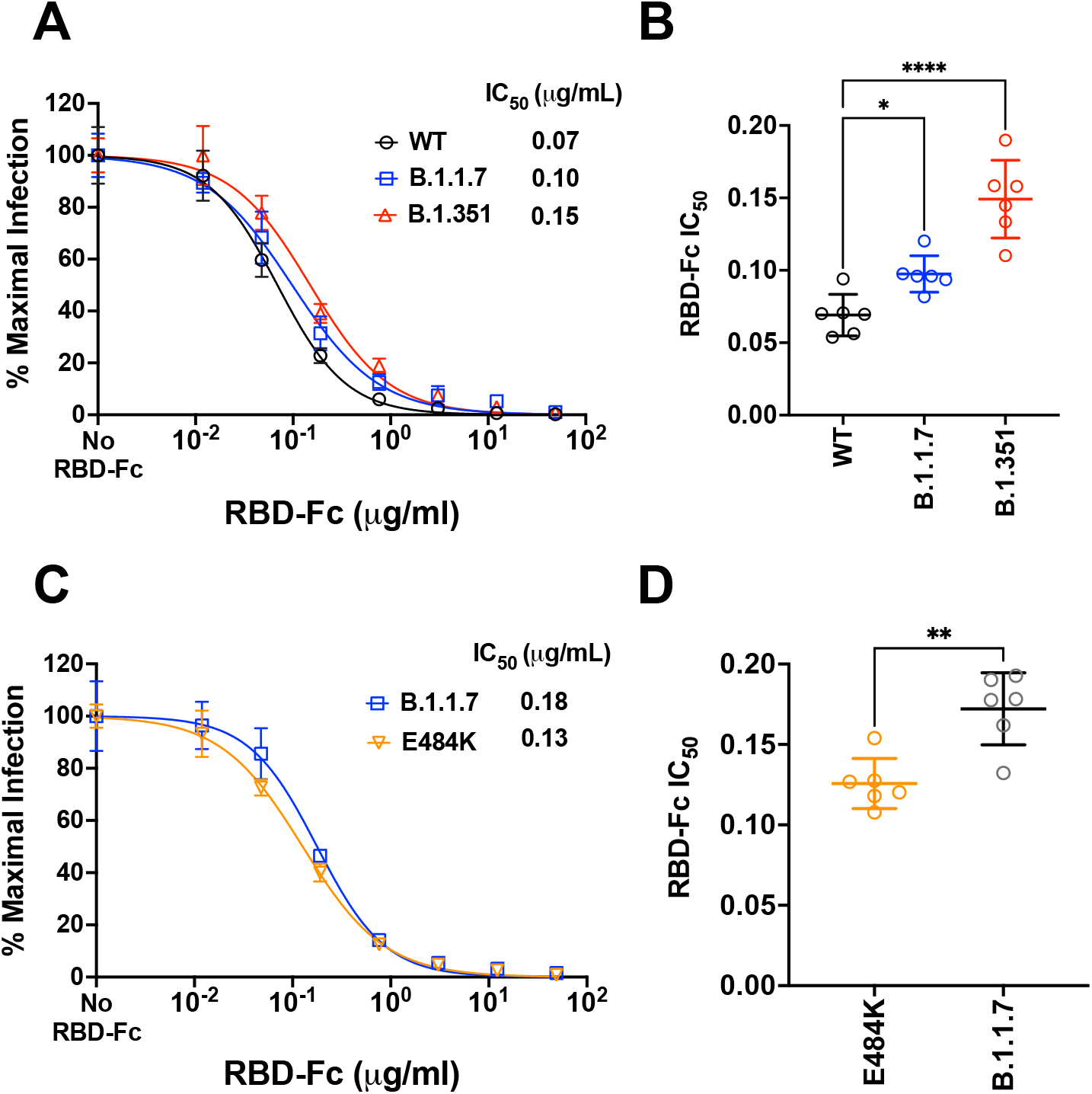
Competitive inhibition of rcVSV-CoV2-S entry by soluble RBD-Fc. **(A)** Recombinant RBD-Fc was serially titrated with the infection inoculum containing a fixed amount of rcVSV-CoV2-S bearing WT or the indicated VOC spike proteins. 10 hpi, GFP+ cells were quantified by the Celigo image cytometer. Data points are means of six independent replicates with error bars representing S.D. The number of GFP+ cells in the absence of any RBD-Fc was set to 100% and used to normalize the infection response in the presence of increasing amounts of RBD-Fc. Log[inhibitor] versus normalized response variable slope nonlinear regression curves were generated using GraphPad PRISM (v9.1.0). **(B)** The IC50 values from each replicate dose response curve generated for a given virus were grouped. The mean (central bar) and SD (whiskers) for each group are indicated. Adjusted p values (*; p<0.05, **; p<0.01, ****; p<0.0001) from ordinary one-way ANOVA with Dunnett’s multiple comparisons test are indicated. **(C)** is a repeat of the experiment done in A with the E484K mutant using a different preparation of recombinant RBD-Fc (see methods). B.1.1.7 serves as the common reference control. **(D)** The IC50 values were calculated and analyzed as in (B).

## DISCUSSION

A key public health concern related to emergent SARS-CoV-2 variants is that by incrementally accruing mutations that escape neutralizing antibodies, they will penetrate herd immunity and spread to reach unvaccinated individuals, some of whom will be susceptible to severe or fatal disease.

Three of the six COVID-19 vaccines currently in use worldwide, namely Moderna mRNA-1273, BioNTech BNT162b2, and Janssen Ad26.COV2.S, each express S harboring K986P and V987P substitutions (2P) within a loop abutting the central helix of the S2’ membrane fusion machinery ^51–53^. This modification locks the spike in a prefusion conformation and elicits higher titers of neutralizing antibodies ^54,55^. The Janssen vaccine has an additional deletion in the furin cleavage site, while the yet-to-be approved Novavax vaccine contains arrays of stabilized spikes conjugated onto a nanoparticle (Table 2). Of the three vaccines that do not appear to make use of 2P Spike mutants, Gamaleya’s Sputnik V and AstraZeneca’s AZD1222 are adenovirus-vectored vaccines encoding native S. The third is CoronaVac, a preparation of inactivated SARS-CoV-2 virions. Although all six vaccines are highly efficacious at preventing severe COVID-19 outcomes, they do not all uniformly prevent infection. Moreover, in all cases thus far examined, these first generation vaccines are less effective against variants with certain non-synonymous substitutions in Spike, such as E484K.

**Table 2.**
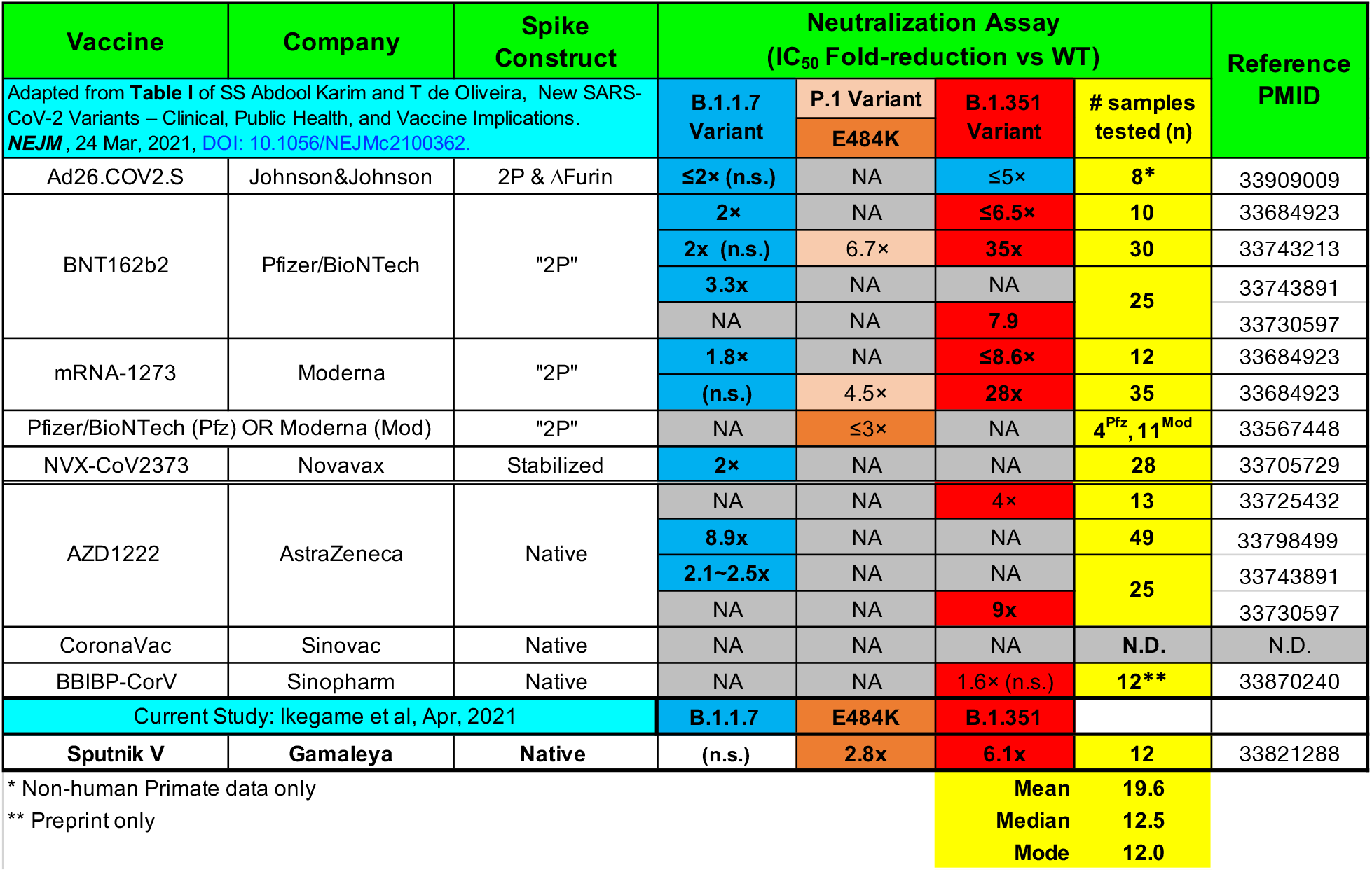
Summary of post-vaccine sera evaluated for neutralization potency against the indicated SARS-CoV-2 variants of concern (VOC). Table format adapted and updated from Abdool Karim and de Olivera^60^.

The most concerning variants are those with multiple mutations in the receptor binding domain (RBD) that confer both enhanced affinity for the hACE2 receptor and escape from neutralizing antibody responses ^17,24,27,33,56,57^. B.1.351 and P.1 have in common three RBD substitutions (K417N/T, E484K and N501Y) whereas B.1.351, P.1 and B.1.1.7 contain the N501Y substitution. Although B.1.1.7 shows enhanced transmissibility and more severe disease outcomes^52^, it does not appear to be consistently more resistant to serum neutralizing responses elicited by vaccines or natural infection ^58,59^. The same is not true, however, for the B.1.351 variant.

In live virus plaque reduction neutralization assays, sera from AstraZeneca vaccine recipients in South Africa exhibited 4.1 to 32.5-fold reduction in neutralizing activity against B.1.351 ^33^. The actual reduction is even more marked because 7 of 12 vaccine recipients who had neutralizing activity against the parental B.1.1 variant, had undetectable neutralization against the B.1.351 strain. Comparator sera from recipients of Moderna and BioNTech mRNA vaccines showed smaller, 6.5 to 8.6-fold reductions in neutralization ^60^.

As of this writing, there are no peer-reviewed data on the protective efficacy of Sputnik V and CoronaVac against SARS-CoV-2 S variants. Here, we showed that sera from Sputnik vaccine recipients in Argentina had a median 6.1-fold and 2.8-fold reduction in GMT against B.1.351 and the E484K mutant spike, respectively. Even more revealing is their dose-response curves. When extrapolated to full serum strength, half of the sera samples failed to achieve an IC_80_ and only 1 out 12 achieved an IC_90_ against B.1.351 (Fig. 4A). Table 2 summarizes peer-reviewed studies that have tested post-vaccination sera from the major vaccines against the VOC/mutant spikes used in this study. Our study shows a similar mean reduction in GMT (reciprocal IC50) against E484K and B.1.351 using 1-month post-Sputnik vaccine sera when compared to other vaccines. Our sample number is admittedly small but matches the median and modal number used in other studies to date. Nonetheless, we caution that comparing only the mean reduction in IC50 can be misleading as an aggregate measure of serum neutralizing activity. The neutralization curves for B.1.351 in our study are not classically sigmodal and have significantly shallower slopes than WT, B.1.17 and E484K, which result in ≤ 90% neutralization for all but one sample when extrapolated to full serum strength. The possible mechanisms for the varying slope values are discussed below.

E484K is present not only as part of an ensemble of RBD mutations present in B.1.351 and P.1, but in many of the 17 lineages detected from South America that carry it, such as P.2, E484K is the only RBD substitution (Supplementary Table 1). A more detailed report covering the genomic surveillance efforts in Argentina that detected the VOC which spurred our study is currently in preparation (Dr. Claudia Perandones, personal communication).

While the E484K substitution appears to be a common route of escape from many RBD-targeting monoclonal antibodies, it is somewhat surprising that a single mutation can confer a significant degree of neutralization resistance from polyclonal responses. Nonetheless, our data show that resistance conferred by E484K mutation be overcome by higher titer antibodies present in undiluted patient sera. But the neutralization resistance conferred by the suite of mutations present in B.1.351 appears qualitatively different. In the majority of cases, the slope of the dose response curve indicates a failure to neutralize even at full strength. We had previously shown that the dose-response curve slope is a major predictor of therapeutic potency for HIV broadly neutralizing antibodies at clinically relevant concentrations ^42^. Importantly, the slope parameter is independent of IC50 but is specifically related to an antibody’s epitope class. Here, we show that defining the neutralization phenotype of a given spike variant or mutant by both its relative IC50 and slope provides a fuller characterization of serum neutralizing activity against SARS-CoV-2 and emergent VOC.

The deletion of residue 242-244 in the NTD of the B.1.351 spike appear to cause large-scale resurfacing of the NTD antigenic surface resulting in greater conformational heterogeneity^44^. Variable neutralization responses across such a heterogenous virus population may result in the shallow slopes (<1) seen. Furthermore, three major classes of neutralizing antibodies (RBS-A, -B, and -C) identified from convalescent patients are sensitive to either the K417N (RBS-A) or E484K (RBS-B and -C) substitutions present in B.1.351. On the other hand, at high serum concentrations, co-operative effects from low-affinity spike binding antibodies or increased spike occupancy by different classes of antibodies can result in the steep Hill slope observed. The steep Hill slopes (>1.0) observed for E484K suggest such co-operative effects might be occurring.

The emergence of variants is fluid situation. B.1.427/1.429, B.1.526, and B.1.617 are other emergent VOI/VOC that could be tested. These strains have substitutions in the RBD (L452R and E484K/Q) and elsewhere in the spike that might confer some degree of neutralization resistance in our in vitro assays. However, all vaccines are effective against most variants, and we do not know what degree of resistance in *in vitro* assays translate to a decrease in the real world efficacy of any given vaccine.

Although we stress that the Gameyla Sputnik V vaccine is likely to retain strong efficacy at preventing severe COVID-19, even in the case of infection by VOC, our data reveal a concerning potential of B.1.351, and to a lesser extent, any variant carrying the E484K substitution (e.g. P.2), to escape the neutralizing antibody responses that this immunization elicits. Furthermore, we acknowledge that *in vivo* protective efficacy can be derived from Fc effector functions of antibodies that bind but do not neutralize. In addition, an adenoviral vectored vaccine should induce potent cell-mediated immunity against multiple epitopes, which were not measured in our study. Nevertheless, given the crucial roles neutralizing antibodies play in preventing infection, our results suggest that updated SARS-CoV-2 vaccines will be necessary to eliminate the virus.

## Materials and Methods

### Cell lines

Vero-CCL81 TMPRSS2, HEK 293T-hACE2 (clone 5-7), and 293T-hACE2-TMPRSS2 (clone F8-2) cells were described previously ^41^, and were maintained in DMEM + 10%FBS. The HEK 293T-hACE2-TMPRSS2 cells were plated on collagen coated plates or dishes. BSR-T7 cells ^61^, which stably express T7-polymerase were maintained in DMEM with 10% FBS.

### VSV-eGFP-CoV2 spike (Δ21aa) genomic clone and helper plasmids

We cloned VSV-eGFP sequence into pEMC vector (pEMC-VSV-eGFP), which includes an optimized T7 promoter and hammerhead ribozyme just before the 5’ end of the viral genome. The original VSV-eGFP sequence was from pVSV-eGFP, a generous gift of Dr. John Rose ^62^.

We generated pEMC-VSV-eGFP-CoV2-S (Genbank Accession: MW816496) as follows: the VSV-G open reading frame of pEMC-VSV-eGFP was replaced with the SARS-CoV-2 S, truncated to lack the final 21 amino acids ^63^. We introduced a Pac-I restriction enzyme site just after the open reading frame of S transcriptional unit, such that the S transcriptional unit is flanked by MluI / PacI sites. SARS-CoV-2 S is from pCAGGS-CoV-2-S ^64^, which codes the codon optimized S from the Wuhan Hu-1 isolate (NCBI ref. seq. NC_045512.2) with a point mutation of D614G, resulting in B.1 lineage. The B.1.1.7 Spike we used carries the mutations found in GISAID Accession Number EPI_ISL 668152: del 69-70, del145, N501Y, A570D, D614G, P681H, T716I, S982A, and D1118H. The B.1.351 Spike carries the mutations D80A, D215G, del242-244, K417N, E484K, N501Y, D614G, and A701V (from EPI_ISL_745109). The Spike sequences of WT, B.1.1.7, B.1.351, and E484K are available at Genbank (Accession Numbers: MW816497, MW816498, MW816499, and MW816500; please also see Supplemental Table 2).

Sequences encoding the VSV N, P, M, G, and L proteins were also cloned into pCI vector to make expression plasmids for virus rescue, resulting in plasmids: pCI-VSV-N, pCI-VSV-P, pCI-VSV-M, pCI-VSV-G, and pCI-VSV-L. These accessory plasmids were a kind gift from Dr. Benjamin tenOever.

### Generation of VSV-CoV2 spike from cDNA

4 × 10^5^ 293T-ACE2-TMPRSS2 cells per well were seeded onto collagen-I coated 6 well plates. The next day, 2000 ng of pEMC-VSV-EGFP-CoV2 spike, 2500 ng of pCAGGS-T7opt ^65^, 850 ng of pCI-VSV-N, 400 ng of pCI-VSV-P, 100 ng of pCI-VSV-M, 100 ng of pCI-VSV-G, 100 ng of pCI-VSV-L were mixed with 4 mL of Plus reagent and 6.6 mL of Lipofectamine LTX (Invitrogen). 30 min later, transfection mixture was applied to 293T-hACE2-TMPRSS2 cells in a dropwise fashion. Cells were maintained with medium replacement every day for 4 to 5 days until GFP positive syncytia appeared. Rescued viruses were amplified in Vero-CCL81 TMPRSS2 cells ^41^, then titered and used for the assay.

### Virus neutralization assay

5 × 10E4 293T-hACE2-TMPRSS2 cells per well were seeded onto collagen-coated 96 well cluster plates one day prior to use in viral neutralization assays. Virus stocks were mixed with serially diluted serum for 10 minutes at room temperature, then infected to cells. Note: all sera assayed in this study were previously heat inactivated by 56 degrees for 30 min before use in any viral neutralization studies. At 10 h post infection, GFP counts were counted by Celigo imaging cytometer (Nexcelom). Each assay was done in triplicate. For calculation of IC50, GFP counts from “no serum” conditions were set to 100%; GFP counts of each condition (serum treated) were normalized to no serum control well. Inhibition curves were generated using Prism 8.4.3 (GraphPad Software) with ‘log (inhibitor) vs normalized response - variable slope’ settings.

### Design of RBD-Fc producing Sendai virus

Sendai virus (SeV) Z strain cDNA sequence (AB855655.1) was generated and cloned into pRS vector with the addition of eGFP transcriptional unit at the head of SeV genome. The sequence of F transcriptional unit was from SeV fushimi strain (KY295909.1) due to the cloning reason. We refer to the pRS-based plasmid coding this sequence as pRS-SeVZ-GFP-F^fushimi^ in this paper. For the introduction of foreign gene into SeV, we generated additional transcriptional unit for RBD-Fc between P gene and M gene. RBD-Fc construct was generated as below; codon optimized DNA sequence of from SARS-CoV-2 spike (MN908947) in pCAGGS a gift of Dr. Florian Krammer ^64^. S amino acids 319 – 541 (corresponding to the RBD domain) sequence were C-terminally fused to the Fc region of human IgG_1_ (220 – 449 aa of P0DOX5.2)

### Generation of recombinant Sendai virus from cDNA

2×10E5 BSR-T7 cells per well were seeded onto 6-well cluster plates. The next day, 4 µg of pRS-SeVZ-GFP-F^fushimi^, 4 µg of pCAGGS-T7opt, 1.44 µg of SeV-N, 0.77 ug of SeV-P, 0.07 ug of SeV-L were mixed with 5.5 µl of Plus reagent and 8.9 µl of Lipofectamine LTX (Invitrogen). 30 min later, transfection mixtures were applied to Bsr-T7 cells in a dropwise fashion, as described previously ^65^. At one day post transfection, medium was replaced with DMEM + 0.2 µg/ml of TPCK-trypsin (Millipore Sigma, #T1426), with subsequent medium replacement each day until infection reached 100% cytopathic effect. Supernatants were stored at -80°C until use in experiments.

### Titration of viruses

For SeV titration, 2 x 10E4 Bsr-T7 cells per well were seeded onto 96-well plates. The next day, 100 µL of serially diluted virus stock (in DMEM + 10% FBS) were applied to each well. GFP positive foci were counted at 24 hours post infection using a Celigo imaging cytometer (Nexcelom, Inc.). Infectivity is presented in infectious units (IU) per mL.

For VSV-CoV2 titration, 5 x 10E4 293T-hACE2-TMPRSS2 cells per well were seeded onto a collagen-coated 96 well plate. Serially diluted virus stocks were then applied to the cells, and GFP positivity was scored at 10 h post infection using a Celigo imaging cytometer.

### Production of proteins and purification

5×10E6 Bsr-T7 cells are seeded in T175cm^2^-flask one day before infection. Cells were infected by SeV at MOI of 0.1 for one hour, followed by replacement of medium with DMEM supplemented with 0.2 mg/mL TPCK-trypsin. Medium was replaced with fresh 0.2 mg/ml TPCK-trypsin containing DMEM each day until infection reached 100% CPE, at which point medium was exchanged for DMEM lacking TPCK-trypsin. Cells were incubated for additional 24 h to allow protein production. Supernatants were centrifuged at 360 *g* for 5 min, then filtered with 0.1 µm filter (Corning^®^ 500 mL Vacuum Filter/Storage Bottle System, 0.1 µm Pore) to remove virions and debris. Supernatant including RBD-Fc were applied to Protein G Sepharose (Millipore Sigma, #GE17-0618-01) containing column (5ml Polypropylene Columns; ThermoFisher, #29922), followed by wash and elution.

### Human Subjects Research

Human subjects research was conducted following the Declaration of Helsinki and related institutional and local regulations. Studies and serum collection relating to the Sputnik vaccine at ANLIS Dr. Carlos G. Malbrán (NatIonal Administration Laboratories and Health Institutes - Carlos G. Malbrán, Argentina) were approved by the Research Ethics Committee of its Unidad Operativa Centro de Contención Biológica (UOCCB) on 9 Feb 2021.

## Data Availability

The datasets generated during and/or analysed during the current study are available from the corresponding author on reasonable request, except for GISAID sequences, which are available from GISAID.org under the GISAID Data Use Agreement.

## Authors contributions

S.I., C.P., B.H.L., J.P.K, conceived of and supervised the study. C.P., A.E.V. and A.E. supervised, collected, analyzed, and provided materials relevant to this study. S.I. generated VSV-CoV-2 S plasmid and rescued viruses. S.I., G.H., S.K., and M.N.A.S. were involved in the generation of S mutant viruses. S.I, L.B., M.N.A.S., K.Y.O. conducted neutralization assays. S.I. and C.T.H. developed the Sendai virus protein expressing system and purified RBD-Fc protein. S.I., B.H.L., and J.P.K. wrote the paper with input from C.P. and all co-authors.

## Acknowledgements

We gratefully acknowledge all submitting authors and collecting authors on whose work this research is based, and to all researchers, clinicians, and public health authorities who make SARS-CoV-2 sequence data available in a timely manner via the GISAID initiative^66,67^.

## Funding information

We acknowledge the following finding. K.Y.O. was supported by Viral-Host Pathogenesis Training Grant T32 AI07647 and additionally by a NRSA F31 AI154739. S.I. and C.-T.H. were supported by postdoctoral fellowships from CHOT-SG (Fukuoka University, Japan) and the Ministry of Science and Technology (MOST, Taiwan), respectively. B.L. acknowledges flexible funding support from NIH grants AI123449 and AI138921; a grant from the Department of Microbiology, Icahn School of Medicine at Mount Sinai; and the Ward-Coleman estate, which endowed the Ward-Coleman Chairs at the ISMMS. J.P.K. was supported by a COVID-19 Fast Grants award from Emergent Ventures, an initiative of the Mercatus Center at George Mason University, and by an intramural grant and other funding from the Office of the Vice Chancellor for Research at LSU Health Sciences Center Shreveport (J.P.K., M.N.A.S.). Processing costs recovered from multiple users of our standardized SARS-CoV-2 VSV pseudotyped particles provided additional support (BL). Work at ANLIS-MALBRAN (A.E.V., A.E., C.P.) was supported by the Ministry of Health (Ministerio de Salud), Argentina.

**SUPPLEMENTAL TABLE 1. Acknowledgement of S: E484K viruses from South America shared on GISAID**.

**SUPPLEMENTAL TABLE 2. Acknowledgement of B.1.1.7 and B.1.351 viruses used for selection of S variants evaluated in this study**.

**Extended Data Figure S1.**
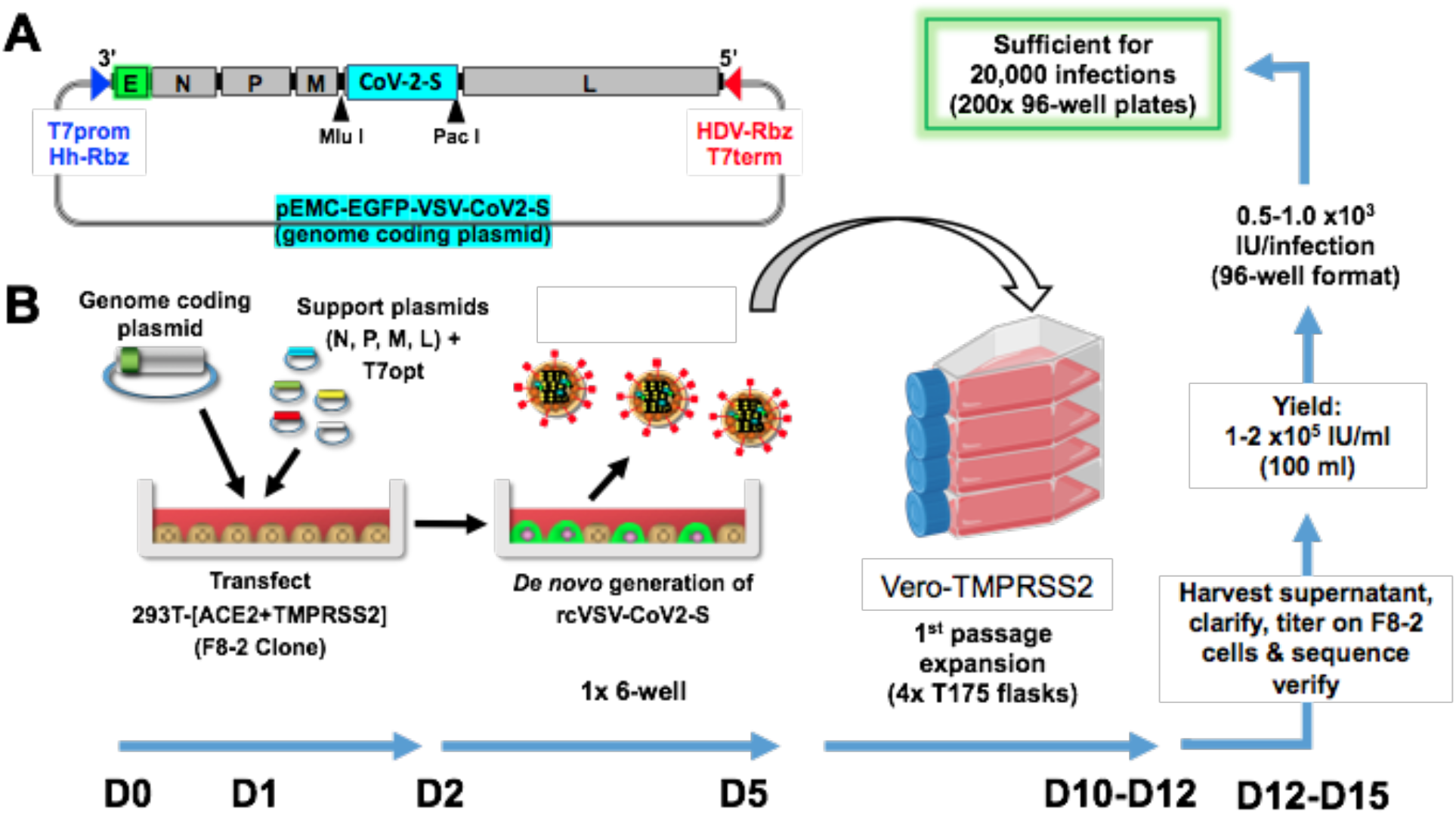
Robust and efficient generation of an EGFP-reporter replication-competent VSV bearing SARS-CoV-2 spike (rcVSV-CoV2-S). **(A)** Schematic of the rcVSV-CoV2-S genomic coding construct and the virus rescue procedure. The maximal T7 promoter (T7prom) followed by a hammer-head ribozyme (HhRbz) and the HDV ribozyme (HDVRbz) plus T7 terminator (T7term) are positioned at the 3’ and 5’ ends of the viral cDNA, respectively. An EGFP(E) transcriptional unit is placed at the 3’ terminus to allow for high level transcription. SARS-CoV-2-S is cloned in place of VSV-G using the indicated restriction sites designed to facilitate easy exchange of spike variant or mutants. (**B)** For virus rescue, highly permissive 293T cells stably expressing human ACE2 and TMPRSS2 (293T-[ACE2+TMPRSS2], F8-2 clone) cells were transfected with the genome coding plasmid, helper plasmids encoding CMV-driven N, P, M, and L genes, and pCAGS encoding codon-optimized T7-RNA polymerase(T7opt). 48-72 hpi, transfected cells turn EGFP+ and start forming syncytia. Supernatant containing rcVSV-CoV2-S are then amplified in Vero-TMPRSS2 cells at the scale shown. The blue arrows at the bottom indicate the timeline for production of each sequence verified stock.

