## Supplemental File 1 for "Neutralizing activity of Sputnik V vaccine sera against SARS-CoV-2 variants"

We gratefully acknowledge the following Authors from the Originating laboratories responsible for obtaining the specimens, as well as the Submitting laboratories where the genome data were generated and shared via GISAID, on which this research is based.

All Submitters of data may be contacted directly via www.gisaid.org

Authors are sorted alphabetically.

| Accession ID | Originating Laboratory | Submitting Laboratory | Authors |
|---|---|---|---|
| EPI_ISL_1000671, EPI_ISL_1000673, EPI_ISL_1000675, EPI_ISL_1000677 | Instituto de Biotecnologia - UNESP-Botucatu-SP | Instituto de Biotecnologia - UNESP-Botucatu-SP | Leila Sabrina Ullmann; Fábio Sossai Possebon, Camila Dantas Malossi, Paula Rahal, Paulo Inacio da Costa, João Pessoa Araújo Jr. |
| EPI_ISL_1001384, EPI_ISL_1001385, EPI_ISL_1001399, EPI_ISL_1001401, EPI_ISL_1001402, EPI_ISL_1001412, EPI_ISL_1001416, EPI_ISL_1001418, EPI_ISL_1001419, EPI_ISL_1001421, EPI_ISL_1001422, EPI_ISL_1001424, EPI_ISL_1001427, EPI_ISL_1001431, EPI_ISL_1001437, EPI_ISL_1001438, EPI_ISL_1001440, EPI_ISL_1001441, EPI_ISL_1001442, EPI_ISL_1001444, EPI_ISL_1001449, EPI_ISL_1001452 | | | |
| see above | Outre mer | National Reference Center for Viruses of Respiratory Infections, Institut Pasteur, Paris | Marion Barbet, Sylvie Behillil, Méline Bizard, Angela Brisebarre, Camille Capel, Etienne Simon-Lorière, Vincent Enouf, Maud Vanpeene, Sylvie van der Werf,Rousset (Guy) Dominique |
| EPI_ISL_1013438, EPI_ISL_1013441, EPI_ISL_1013442, EPI_ISL_1013447, EPI_ISL_1013448 | Laboratoire associé au CNR des Virus Influenzae | National Reference Center for Viruses of Respiratory Infections, Institut Pasteur, Paris | Marion Barbet, Sylvie Behillil, Méline Bizard, Angela Brisebarre, Camille Capel, Etienne Simon-Lorière, Vincent Enouf, Maud Vanpeene, Sylvie van der Werf,Rousset (Guy) Dominique |
| EPI_ISL_1014645, EPI_ISL_1014646 | Dutch COVID-19 response team | National Institute for Public Health and the Environment (RIVM) | Adam Meijer, Harry Vennema, Dirk Eggink, Jeroen Cremer, Sharon van den Brink, Bas van der Veer, AnneMarie van den Brandt, Florian Zwagemaker, Dennis Schmitz, Chantal Reusken, on behalf of the national COVID-19 response team |
| EPI_ISL_1017705 | Instituto Nacional de Salud- Dirección de Redes de Laboratorios de Salud Pública | Instituto Nacional de Salud- Dirección de Investigación en Salud Pública | Katherine Laiton-Donato, Diego A. Álvarez-Díaz, Carlos Franco-Muñoz, Hector Alejandro Ruiz-Moreno, Maria T. Herrera-Sepúlveda, Diego Andrés Prada, Jhonnatan Reales-González, Sheryll Corchuelo, Julian Naizaque, Gerardo Santamaría, Magdalena Wiesner, Martha Lucia Ospina Martinez, Marcela Mercado-Reyes |
| EPI_ISL_1034304, EPI_ISL_1034306 | Laboratorio de Ecologia de Doencas Transmissiveis na Amazonia, Instituto Leonidas e Maria Deane - Fiocruz Amazonia | Laboratorio de Ecologia de Doencas Transmissiveis na Amazonia, Instituto Leonidas e Maria Deane - Fiocruz Amazonia | Valdinete Nascimento, Victor Souza, André Corado, Fernanda Nascimento, George Silva, Ágatha Costa, Debora Duarte, Karina Pessoa, Matilde Mejía, Luciana Gonçalves, Maria Júlia Brandão, Michele Jesus, Felipe Naveca on behalf of the Fiocruz COVID-19 Genomic Surveillance Network |
| EPI_ISL_1035785 | Dutch COVID-19 response team | National Institute for Public Health and the Environment (RIVM) | Adam Meijer, Harry Vennema, Dirk Eggink, Jeroen Cremer, Sharon van den Brink, Bas van der Veer, AnneMarie van den Brandt, Florian Zwagemaker, Dennis Schmitz, Chantal Reusken, on behalf of the national COVID-19 response team |
| EPI_ISL_1036756 | Laboratorio de Salud Pública de Bogotá | Instituto Nacional de Salud- Direccion de Investigacion en Salud Publica | Katherine Laiton-Donato, Diego A. Álvarez-Díaz, Carlos Franco-Muñoz, Hector Alejandro Ruiz-Moreno, Maria T. Herrera-Sepúlveda, Diego Andrés Prada, Jhonnatan Reales-González, Sheryll Corchuelo, Julian Naizaque, Gerardo Santamaría, Magdalena Wiesner, Martha Lucia Ospina Martinez, Marcela Mercado-Reyes |
| EPI_ISL_1039691, EPI_ISL_1039692, EPI_ISL_1039693, EPI_ISL_1039694, EPI_ISL_1039695, EPI_ISL_1041509 | LACEN do Estado de Goias | Instituto Adolfo Lutz, Interdiciplinary Procedures Center, Strategic Laboratory | Claudio Tavares Sacchi, Claudia Regina Gonçalves, Erica Valessa Ramos Gomes, Karoline Rodrigues Campos |
| EPI_ISL_1060571, EPI_ISL_1060577, EPI_ISL_1060579, EPI_ISL_1060581, EPI_ISL_1060587, EPI_ISL_1060590, EPI_ISL_1060591, EPI_ISL_1060592 | Outre mer | National Reference Center for Viruses of Respiratory Infections, Institut Pasteur, Paris | Marion Barbet, Sylvie Behillil, Méline Bizard, Angela Brisebarre, Camille Capel, Etienne Simon-Lorière, Vincent Enouf, Maud Vanpeene, Sylvie van der Werf,Rousset Dominique |
| EPI_ISL_1060876 | DB Diagnosticos do Brasil | Instituto de Medicina Tropical de Sao Paulo | Brazil-UK Centre for Arbovirus Discovery Diagnosis Genomics and Epidemiology (CADDE) Genomic Network - Instituto de Medicina Tropical |
| EPI_ISL_1060881 | CDL Laboratorio Santos e Vidal LTDA. | Instituto de Medicina Tropical de Sao Paulo | Brazil-UK Centre for Arbovirus Discovery Diagnosis Genomics and Epidemiology (CADDE) Genomic Network - Instituto de Medicina Tropical |
| EPI_ISL_1060886 | DB Diagnosticos do Brasil | Instituto de Medicina Tropical de Sao Paulo | Brazil-UK Centre for Arbovirus Discovery Diagnosis Genomics and Epidemiology (CADDE) Genomic Network - Instituto de Medicina Tropical |
| EPI_ISL_1060888, EPI_ISL_1060889, EPI_ISL_1060890, EPI_ISL_1060895, EPI_ISL_1060898, EPI_ISL_1060899 | CDL Laboratorio Santos e Vidal LTDA. | Instituto de Medicina Tropical de Sao Paulo | Brazil-UK Centre for Arbovirus Discovery Diagnosis Genomics and Epidemiology (CADDE) Genomic Network - Instituto de Medicina Tropical |
| EPI_ISL_1060900, EPI_ISL_1060902, EPI_ISL_1060904 | DB Diagnosticos do Brasil | Instituto de Medicina Tropical de Sao Paulo | Brazil-UK Centre for Arbovirus Discovery Diagnosis Genomics and Epidemiology (CADDE) Genomic Network - Instituto de Medicina Tropical |
| EPI_ISL_1060905, EPI_ISL_1060906, EPI_ISL_1060911, EPI_ISL_1060912, EPI_ISL_1060913 | CDL Laboratorio Santos e Vidal LTDA. | Instituto de Medicina Tropical de Sao Paulo | Brazil-UK Centre for Arbovirus Discovery Diagnosis Genomics and Epidemiology (CADDE) Genomic Network - Instituto de Medicina Tropical |
| EPI_ISL_1060914 | DB Diagnosticos do Brasil | Instituto de Medicina Tropical de Sao Paulo | Brazil-UK Centre for Arbovirus Discovery Diagnosis Genomics and Epidemiology (CADDE) Genomic Network - Instituto de Medicina Tropical |
| EPI_ISL_1060915, EPI_ISL_1060917 | CDL Laboratorio Santos e Vidal LTDA. | Instituto de Medicina Tropical de Sao Paulo | Brazil-UK Centre for Arbovirus Discovery Diagnosis Genomics and Epidemiology (CADDE) Genomic Network - Instituto de Medicina Tropical |
| EPI_ISL_1060918 | DB Diagnosticos do Brasil | Instituto de Medicina Tropical de Sao Paulo | Brazil-UK Centre for Arbovirus Discovery Diagnosis Genomics and Epidemiology (CADDE) Genomic Network - Instituto de Medicina Tropical |
| EPI_ISL_1060919, EPI_ISL_1060921, EPI_ISL_1060922 | CDL Laboratorio Santos e Vidal LTDA. | Instituto de Medicina Tropical de Sao Paulo | Brazil-UK Centre for Arbovirus Discovery Diagnosis Genomics and Epidemiology (CADDE) Genomic Network - Instituto de Medicina Tropical |
| EPI_ISL_1060923, EPI_ISL_1060925, EPI_ISL_1060926 | DB Diagnosticos do Brasil | Instituto de Medicina Tropical de Sao Paulo | Brazil-UK Centre for Arbovirus Discovery Diagnosis Genomics and Epidemiology (CADDE) Genomic Network - Instituto de Medicina Tropical |
| EPI_ISL_1060931, EPI_ISL_1060935, EPI_ISL_1061032, EPI_ISL_1064737 | CDL Laboratorio Santos e Vidal LTDA. | Instituto de Medicina Tropical de Sao Paulo | Brazil-UK Centre for Arbovirus Discovery Diagnosis Genomics and Epidemiology (CADDE) Genomic Network - Instituto de Medicina Tropical |
| EPI_ISL_1067728 | Center for Biotechnology and Cell Therapy, São Rafael Hospital, Salvador, Brazil | Central Public Health Laboratory - LACEN -Bahia, Salvador, Brazil | Stephane Tosta, Luciana Oliveira, Vanessa Nardy,Patrícia Cajado,Marcela Gómez, Breno Dominguez, Jaqueline Gomes, Vagner Fonseca,Marta Giovanetti,Luiz Alcantara, Felicidade Pereira, Arabela Leal |
| EPI_ISL_1067729, EPI_ISL_1067730, EPI_ISL_1067731 | Central Public Health Laboratory - LACEN -Bahia, Salvador, Brazil | Central Public Health Laboratory - LACEN -Bahia, Salvador, Brazil | Stephane Tosta, Luciana Oliveira, Vanessa Nardy,Patrícia Cajado,Marcela Gómez, Breno Dominguez, Jaqueline Gomes, Vagner Fonseca,Marta Giovanetti,Luiz Alcantara, Felicidade Pereira, Arabela Leal |
| EPI_ISL_1067732 | Center for Biotechnology and Cell Therapy, São Rafael Hospital, Salvador, Brazil | Central Public Health Laboratory - LACEN -Bahia, Salvador, Brazil | Stephane Tosta, Luciana Oliveira, Vanessa Nardy,Patrícia Cajado,Marcela Gómez, Breno Dominguez, Jaqueline Gomes, Vagner Fonseca,Marta Giovanetti,Luiz Alcantara, Felicidade Pereira, Arabela Leal |
| EPI_ISL_1067733, EPI_ISL_1067734, EPI_ISL_1067735 | Central Public Health Laboratory - LACEN -Bahia, Salvador, Brazil | Central Public Health Laboratory - LACEN -Bahia, Salvador, Brazil | Stephane Tosta, Luciana Oliveira, Vanessa Nardy,Patrícia Cajado,Marcela Gómez, Breno Dominguez, Jaqueline Gomes, Vagner Fonseca,Marta Giovanetti,Luiz Alcantara, Felicidade Pereira, Arabela Leal |
| EPI_ISL_1067736 | Center for Biotechnology and Cell Therapy, São Rafael Hospital, Salvador, Brazil | Central Public Health Laboratory - LACEN -Bahia, Salvador, Brazil | Stephane Tosta, Luciana Oliveira, Vanessa Nardy,Patrícia Cajado,Marcela Gómez, Breno Dominguez, Jaqueline Gomes, Vagner Fonseca,Marta Giovanetti,Luiz Alcantara, Felicidade Pereira, Arabela Leal |
| EPI_ISL_1067737, EPI_ISL_1067738 | Central Public Health Laboratory - LACEN -Bahia, Salvador, Brazil | Central Public Health Laboratory - LACEN -Bahia, Salvador, Brazil | Stephane Tosta, Luciana Oliveira, Vanessa Nardy,Patrícia Cajado,Marcela Gómez, Breno Dominguez, Jaqueline Gomes, Vagner Fonseca,Marta Giovanetti,Luiz Alcantara, Felicidade Pereira, Arabela Leal |

| | | | |
|---|---|---|---|
| EPI_ISL_1068108, EPI_ISL_1068110, EPI_ISL_1068111, EPI_ISL_1068112, EPI_ISL_1068114, EPI_ISL_1068115, EPI_ISL_1068140, EPI_ISL_1068142, EPI_ISL_1068145, EPI_ISL_1068149, EPI_ISL_1068150, EPI_ISL_1068151, EPI_ISL_1068153, EPI_ISL_1068154, EPI_ISL_1068156, EPI_ISL_1068157, EPI_ISL_1068158, EPI_ISL_1068159, EPI_ISL_1068160, EPI_ISL_1068169, EPI_ISL_1068198, EPI_ISL_1068221, EPI_ISL_1068222, EPI_ISL_1068223, EPI_ISL_1068225, EPI_ISL_1068226, EPI_ISL_1068243, EPI_ISL_1068248, EPI_ISL_1068249, EPI_ISL_1068255, EPI_ISL_1068256, EPI_ISL_1068258, EPI_ISL_1068259, EPI_ISL_1068260, EPI_ISL_1068261, EPI_ISL_1068262, EPI_ISL_1068263, EPI_ISL_1068264, EPI_ISL_1068266, EPI_ISL_1068267, EPI_ISL_1068268, EPI_ISL_1068269, EPI_ISL_1068270, EPI_ISL_1068271, EPI_ISL_1068272, EPI_ISL_1068273, EPI_ISL_1068274, EPI_ISL_1068275, EPI_ISL_1068276, EPI_ISL_1068277, EPI_ISL_1068278, EPI_ISL_1068279, EPI_ISL_1068280, EPI_ISL_1068281, EPI_ISL_1068282, EPI_ISL_1068283, EPI_ISL_1068284, EPI_ISL_1068285, EPI_ISL_1068286, EPI_ISL_1068287, EPI_ISL_1068288, EPI_ISL_1068289, EPI_ISL_1068290, EPI_ISL_1068291, EPI_ISL_1068292 | | | |
| see above | Laboratorio de Ecologia de Doencas Transmissiveis na Amazonia, Instituto Leonidas e Maria Deane - Fiocruz Amazonia | Laboratorio de Ecologia de Doencas Transmissiveis na Amazonia, Instituto Leonidas e Maria Deane - Fiocruz Amazonia | Valdinete Nascimento, Victor Souza, André Corado, Fernanda Nascimento, George Silva, Ágatha Costa, Debora Duarte, Karina Pessoa, Matilde Mejía, Luciana Gonçalves, Maria Júlia Brandão, Michele Jesus, Felipe Naveca on behalf of the Fiocruz COVID-19 Genomic Surveillance Network |
| EPI_ISL_1068351, EPI_ISL_1068361, EPI_ISL_1068368 | Central Public Health Laboratory - LACEN -Bahia, Salvador, Brazil | Central Public Health Laboratory - LACEN -Bahia, Salvador, Brazil | Stephane Tosta, Luciana Oliveira, Vanessa Nardy,Patrícia Cajado,Marcela Gómez, Breno Dominguez, Jaqueline Gomes, Vagner Fonseca,Marta Giovanetti,Luiz Alcantara, Felicidade Pereira, Arabela Leal |
| EPI_ISL_1078986, EPI_ISL_1078987, EPI_ISL_1078988, EPI_ISL_1078989, EPI_ISL_1078992, EPI_ISL_1078993, EPI_ISL_1078995, EPI_ISL_1078997, EPI_ISL_1078999, EPI_ISL_1079000, EPI_ISL_1079002, EPI_ISL_1079004, EPI_ISL_1079007, EPI_ISL_1079008, EPI_ISL_1079159, EPI_ISL_1079162, EPI_ISL_1079165 | | | |
| see above | IAL Regional de Bauru | Instituto Adolfo Lutz, Interdiciplinary Procedures Center, Strategic Laboratory | Claudio Tavares Sacchi, Claudia Regina Gonçalves, Erica Valessa Ramos Gomes, Karoline Rodrigues Campos |
| EPI_ISL_1086034 | Diagnosticos da America - DASA | Instituto Adolfo Lutz, Interdiciplinary Procedures Center, Strategic Laboratory | Claudio Tavares Sacchi, Claudia Regina Gonçalves, Erica Valessa Ramos Gomes, Karoline Rodrigues Campos, Caio Vinicius Dias Lopes |
| EPI_ISL_1086035, EPI_ISL_1086036 | IAL Regional de Bauru | Instituto Adolfo Lutz, Interdiciplinary Procedures Center, Strategic Laboratory | Claudio Tavares Sacchi, Claudia Regina Gonçalves, Erica Valessa Ramos Gomes, Karoline Rodrigues Campos, Caio Vinicius Dias Lopes |
| EPI_ISL_1086037, EPI_ISL_1086038, EPI_ISL_1086039, EPI_ISL_1086040, EPI_ISL_1086041, EPI_ISL_1086042, EPI_ISL_1086043 | Diagnosticos da America - DASA | Instituto Adolfo Lutz, Interdiciplinary Procedures Center, Strategic Laboratory | Claudio Tavares Sacchi, Claudia Regina Gonçalves, Erica Valessa Ramos Gomes, Karoline Rodrigues Campos, Caio Vinicius Dias Lopes |
| EPI_ISL_1086044, EPI_ISL_1086045, EPI_ISL_1086046, EPI_ISL_1086047, EPI_ISL_1086048, EPI_ISL_1086049, EPI_ISL_1086050, EPI_ISL_1086052, EPI_ISL_1086053, EPI_ISL_1086054, EPI_ISL_1086055, EPI_ISL_1086057 | | | |
| see above | IAL Regional de Bauru | Instituto Adolfo Lutz, Interdiciplinary Procedures Center, Strategic Laboratory | Claudio Tavares Sacchi, Claudia Regina Gonçalves, Erica Valessa Ramos Gomes, Karoline Rodrigues Campos, Caio Vinicius Dias Lopes |
| EPI_ISL_1086374 | LACEN - Laboratório Central de Saúde Pública do Maranhao | Evandro Chagas Institute | Santos, M.C.; Silva, A.M.; Junior, W.D.C.; Barbagelata, L.S.; Ferreira, J.A.; Sousa, E.M.A.; da Silva, P.S.; Pinheiro, K.C.; L.C.; Sousa Junior, E.C. |
| EPI_ISL_1086375 | LACEN - Laboratório Central de Saúde Pública do Rio Grande do Norte | Evandro Chagas Institute | Santos, M.C.; Silva, A.M.; Junior, W.D.C.; Barbagelata, L.S.; Ferreira, J.A.; Sousa, E.M.A.; da Silva, P.S.; Pinheiro, K.C.; L.C.; Sousa Junior, E.C. |
| EPI_ISL_1091781 | ICMT | Universidad Nacional de Colombia - Laboratorio Genómico One Health | Andres F. Cardona-Rios, Daniel O. Maldonado-Perez, Laura Silvana Perez, Karl A Ciuoderis, Idabely Betancur Ortiz, Sandra Ines Cano, Diego A. Álvarez-Díaz, Carlos Franco-Muñoz, Marcela Mercado-Reyes, Jorge E. Osorio, Juan P. Hernandez-Ortiz |
| EPI_ISL_1092005 | Laboratorio de Salud Publica de Cesar | Instituto Nacional de Salud- Dirección de Investigación en Salud Pública | Katherine Laiton-Donato, Diego A. Álvarez-Díaz, Carlos Franco-Muñoz, Hector Alejandro Ruiz-Moreno, Maria T. Herrera-Sepúlveda, Diego Andrés Prada, Jhonnatan Reales-González, Sheryll Corchuelo, Julian Naizaque, Gerardo Santamaría, Magdalena Wiesner, Martha Lucia Ospina Martinez, Marcela Mercado-Reyes |
| EPI_ISL_1092006 | Laboratorio IMAT | Instituto Nacional de Salud- Dirección de Investigación en Salud Pública | Katherine Laiton-Donato, Diego A. Álvarez-Díaz, Carlos Franco-Muñoz, Hector Alejandro Ruiz-Moreno, Maria T. Herrera-Sepúlveda, Diego Andrés Prada, Jhonnatan Reales-González, Sheryll Corchuelo, Julian Naizaque, Gerardo Santamaría, Magdalena Wiesner, Martha Lucia Ospina Martinez, Marcela Mercado-Reyes |
| EPI_ISL_1092007 | Laboratorio Continental | Instituto Nacional de Salud- Dirección de Investigación en Salud Pública | Katherine Laiton-Donato, Diego A. Álvarez-Díaz, Carlos Franco-Muñoz, Hector Alejandro Ruiz-Moreno, Maria T. Herrera-Sepúlveda, Diego Andrés Prada, Jhonnatan Reales-González, Sheryll Corchuelo, Julian Naizaque, Gerardo Santamaría, Magdalena Wiesner, Martha Lucia Ospina Martinez, Marcela Mercado-Reyes |
| EPI_ISL_1092008 | Laboratorio de Salud Publica de Bogota | Instituto Nacional de Salud- Dirección de Investigación en Salud Pública | Katherine Laiton-Donato, Diego A. Álvarez-Díaz, Carlos Franco-Muñoz, Hector Alejandro Ruiz-Moreno, Maria T. Herrera-Sepúlveda, Diego Andrés Prada, Jhonnatan Reales-González, Sheryll Corchuelo, Julian Naizaque, Gerardo Santamaría, Magdalena Wiesner, Martha Lucia Ospina Martinez, Marcela Mercado-Reyes |
| EPI_ISL_1092360, EPI_ISL_1095913 | IAL Regional de Bauru | Instituto Adolfo Lutz, Interdiciplinary Procedures Center, Strategic Laboratory | Claudio Tavares Sacchi, Claudia Regina Gonçalves, Erica Valessa Ramos Gomes, Karoline Rodrigues Campos |
| EPI_ISL_1096120 | IAL Regional de Bauru | Instituto Adolfo Lutz, Interdiciplinary Procedures Center, Strategic Laboratory | Claudio Tavares Sacchi, Claudia Regina Gonçalves, Erica Valessa Ramos Gomes, Karoline Rodrigues Campos, Caio Vinicius Dias Lopes |
| EPI_ISL_1096121 | Diagnosticos da America - DASA | Instituto Adolfo Lutz, Interdiciplinary Procedures Center, Strategic Laboratory | Claudio Tavares Sacchi, Claudia Regina Gonçalves, Erica Valessa Ramos Gomes, Karoline Rodrigues Campos, Caio Vinicius Dias Lopes |
| EPI_ISL_1096122, EPI_ISL_1096123, EPI_ISL_1096124, EPI_ISL_1096125, EPI_ISL_1096126, EPI_ISL_1096127, EPI_ISL_1096128, EPI_ISL_1096129, EPI_ISL_1096130, EPI_ISL_1096131, EPI_ISL_1096132, EPI_ISL_1096133, EPI_ISL_1096134 | | | |
| see above | IAL Regional de Bauru | Instituto Adolfo Lutz, Interdiciplinary Procedures Center, Strategic Laboratory | Claudio Tavares Sacchi, Claudia Regina Gonçalves, Erica Valessa Ramos Gomes, Karoline Rodrigues Campos, Caio Vinicius Dias Lopes |
| EPI_ISL_1096135 | Diagnosticos da America - DASA | Instituto Adolfo Lutz, Interdiciplinary Procedures Center, Strategic Laboratory | Claudio Tavares Sacchi, Claudia Regina Gonçalves, Erica Valessa Ramos Gomes, Karoline Rodrigues Campos, Caio Vinicius Dias Lopes |
| EPI_ISL_1096136 | IAL Regional de Bauru | Instituto Adolfo Lutz, Interdiciplinary Procedures Center, Strategic Laboratory | Claudio Tavares Sacchi, Claudia Regina Gonçalves, Erica Valessa Ramos Gomes, Karoline Rodrigues Campos, Caio Vinicius Dias Lopes |
| EPI_ISL_1096235, EPI_ISL_1096262 | Outre mer | National Reference Center for Viruses of Respiratory Infections, Institut Pasteur, Paris | Marion Barbet, Sylvie Behillil, Méline Bizard, Angela Brisebarre, Camille Capel, Etienne Simon-Lorière, Vincent Enouf, Maud Vanpeene, Sylvie van der Werf,Rousset Dominique |
| EPI_ISL_1111137, EPI_ISL_1111143, EPI_ISL_1111151, EPI_ISL_1111152, EPI_ISL_1111160, EPI_ISL_1111281, EPI_ISL_1111450, EPI_ISL_1111451, EPI_ISL_1111456, EPI_ISL_1111457, EPI_ISL_1111461, EPI_ISL_1111465, EPI_ISL_1111467, EPI_ISL_1111469, EPI_ISL_1111472, EPI_ISL_1111474, EPI_ISL_1111475, EPI_ISL_1111483, EPI_ISL_1111484, EPI_ISL_1111486, EPI_ISL_1111490, EPI_ISL_1111496 | | | |
| see above | Laboratorio de Referencia Nacional de Virus Respiratorio. Instituto Nacional de Salud Perú | Laboratorio de Referencia Nacional de Enteropatógenos. Instituto Nacional de Salud del Perú | Ronnie Gavilan Chavez, Junior Caro Castro, Willi Quino Sifuentes, Veronica Hurtado Vela, Iris Silva Molina, Fiorella Orellana Peralta |
| EPI_ISL_1121305 | Policlinica Maria Dirce | Instituto Adolfo Lutz, Interdiciplinary Procedures Center, Strategic Laboratory | Claudio Tavares Sacchi, Claudia Regina Gonçalves, Erica Valessa Ramos Gomes, Karoline Rodrigues Campos, Caio Vinicius Dias Lopes |
| EPI_ISL_1121307 | Hospital E Antonio Policarpo de Oliveira | Instituto Adolfo Lutz, Interdiciplinary Procedures Center, Strategic Laboratory | Claudio Tavares Sacchi, Claudia Regina Gonçalves, Erica Valessa Ramos Gomes, Karoline Rodrigues Campos, Caio Vinicius Dias Lopes |
| EPI_ISL_1121308 | Hospital e Pronto Socorro Portinari | Instituto Adolfo Lutz, Interdiciplinary Procedures Center, Strategic Laboratory | Claudio Tavares Sacchi, Claudia Regina Gonçalves, Erica Valessa Ramos Gomes, Karoline Rodrigues Campos, Caio Vinicius Dias Lopes |
| EPI_ISL_1121309 | UBS Jose Francisco Rezende | Instituto Adolfo Lutz, Interdiciplinary Procedures Center, Strategic Laboratory | Claudio Tavares Sacchi, Claudia Regina Gonçalves, Erica Valessa Ramos Gomes, Karoline Rodrigues Campos, Caio Vinicius Dias Lopes |
| EPI_ISL_1121310 | IAL Regional de Bauru | Instituto Adolfo Lutz, Interdiciplinary Procedures Center, Strategic Laboratory | Claudio Tavares Sacchi, Claudia Regina Gonçalves, Erica Valessa Ramos Gomes, Karoline Rodrigues Campos, Caio Vinicius Dias Lopes |
| EPI_ISL_1121311 | Centro de Saude II Dr. Jose Paione Mococa | Instituto Adolfo Lutz, Interdiciplinary Procedures Center, Strategic Laboratory | Claudio Tavares Sacchi, Claudia Regina Gonçalves, Erica Valessa Ramos Gomes, Karoline Rodrigues Campos, Caio Vinicius Dias Lopes |

| | | | |
|---|---|---|---|
| EPI_ISL_1121312, EPI_ISL_1121313, EPI_ISL_1121314, EPI_ISL_1121315 | IAL Regional de Bauru | Instituto Adolfo Lutz, Interdiciplinary Procedures Center, Strategic Laboratory | Claudio Tavares Sacchi, Claudia Regina Gonçalves, Erica Valessa Ramos Gomes, Karoline Rodrigues Campos, Caio Vinicius Dias Lopes |
| EPI_ISL_1121316 | LACEN do Rio Grande do Sul | Instituto Adolfo Lutz, Interdiciplinary Procedures Center, Strategic Laboratory | Claudio Tavares Sacchi, Claudia Regina Gonçalves, Erica Valessa Ramos Gomes, Karoline Rodrigues Campos, Caio Vinicius Dias Lopes |
| EPI_ISL_1121318, EPI_ISL_1121319 | Hospital de Campanha COVID 19 Caieiras | Instituto Adolfo Lutz, Interdiciplinary Procedures Center, Strategic Laboratory | Claudio Tavares Sacchi, Claudia Regina Gonçalves, Erica Valessa Ramos Gomes, Karoline Rodrigues Campos, Caio Vinicius Dias Lopes |
| EPI_ISL_1121320, EPI_ISL_1121321 | IAL Regional de Bauru | Instituto Adolfo Lutz, Interdiciplinary Procedures Center, Strategic Laboratory | Claudio Tavares Sacchi, Claudia Regina Gonçalves, Erica Valessa Ramos Gomes, Karoline Rodrigues Campos, Caio Vinicius Dias Lopes |
| EPI_ISL_1121324 | Complexo Hospitalar Padre Bentode Guarulhos | Instituto Adolfo Lutz, Interdiciplinary Procedures Center, Strategic Laboratory | Claudio Tavares Sacchi, Claudia Regina Gonçalves, Erica Valessa Ramos Gomes, Karoline Rodrigues Campos, Caio Vinicius Dias Lopes |
| EPI_ISL_1121325 | IAL Regional de Santos | Instituto Adolfo Lutz, Interdiciplinary Procedures Center, Strategic Laboratory | Claudio Tavares Sacchi, Claudia Regina Gonçalves, Erica Valessa Ramos Gomes, Karoline Rodrigues Campos, Caio Vinicius Dias Lopes |
| EPI_ISL_1123373 | Grupo Tecnico de Vigilancia Sanitaria e Epidemiologica | Instituto Adolfo Lutz, Interdiciplinary Procedures Center, Strategic Laboratory | Claudio Tavares Sacchi, Claudia Regina Gonçalves, Erica Valessa Ramos Gomes, Karoline Rodrigues Campos, Caio Vinicius Dias Lopes |
| EPI_ISL_1123375 | IAL Regional de Santos | Instituto Adolfo Lutz, Interdiciplinary Procedures Center, Strategic Laboratory | Claudio Tavares Sacchi, Claudia Regina Gonçalves, Erica Valessa Ramos Gomes, Karoline Rodrigues Campos, Caio Vinicius Dias Lopes |
| EPI_ISL_1133120, EPI_ISL_1133121, EPI_ISL_1133122, EPI_ISL_1133123, EPI_ISL_1133124, EPI_ISL_1133125, EPI_ISL_1133126, EPI_ISL_1133127, EPI_ISL_1133128, EPI_ISL_1133129, EPI_ISL_1133130, EPI_ISL_1133131, EPI_ISL_1133132, EPI_ISL_1133133, EPI_ISL_1133134, EPI_ISL_1133135, EPI_ISL_1133136, EPI_ISL_1133137, EPI_ISL_1133138, EPI_ISL_1133139, EPI_ISL_1133140, EPI_ISL_1133141, EPI_ISL_1133142, EPI_ISL_1133143, EPI_ISL_1133144 | | | |
| see above | LABCOVID_HCPA | LABRESIS_HCPA | Martins AF, Wink PL, Volpato F, Rosset C, de Paris F, Monteiro F, Barth AL |
| EPI_ISL_1137478 | Laboratorio de Salud Pública - Secretaría Distrital de Salud | Instituto Nacional de Salud- Dirección de Investigación en Salud Pública | Katherine Laiton-Donato, Carlos Franco-Muñoz, Diego A. Álvarez-Díaz, Hector Alejandro Ruiz-Moreno, Jhonnatan Reales-González, Diego Andrés Prada, Sheryll Corchuelo, Maria T. Herrera-Sepúlveda, Julian Naizaque, Gerardo Santamaría, Magdalena Wiesner, Martha Lucia Ospina Martínez, Marcela Mercado-Reyes. |
| EPI_ISL_1137615 | Laboratorio de Salud Publica de Cauca | Instituto Nacional de Salud- Dirección de Investigación en Salud Pública | Katherine Laiton-Donato, Carlos Franco-Muñoz, Diego A. Álvarez-Díaz, Hector Alejandro Ruiz-Moreno, Jhonnatan Reales-González, Diego Andrés Prada, Sheryll Corchuelo, Maria T. Herrera-Sepúlveda, Julian Naizaque, Gerardo Santamaría, Magdalena Wiesner, Martha Lucia Ospina Martínez, Marcela Mercado-Reyes. |
| EPI_ISL_1137616, EPI_ISL_1137617 | Fundación Cardio Infantil | Instituto Nacional de Salud- Dirección de Investigación en Salud Pública | Katherine Laiton-Donato, Carlos Franco-Muñoz, Diego A. Álvarez-Díaz, Hector Alejandro Ruiz-Moreno, Jhonnatan Reales-González, Diego Andrés Prada, Sheryll Corchuelo, Maria T. Herrera-Sepúlveda, Julian Naizaque, Gerardo Santamaría, Magdalena Wiesner, Martha Lucia Ospina Martínez, Marcela Mercado-Reyes. |
| EPI_ISL_1137619 | Laboratorio de Salud Publica de Cauca | Instituto Nacional de Salud- Dirección de Investigación en Salud Pública | Katherine Laiton-Donato, Carlos Franco-Muñoz, Diego A. Álvarez-Díaz, Hector Alejandro Ruiz-Moreno, Jhonnatan Reales-González, Diego Andrés Prada, Sheryll Corchuelo, Maria T. Herrera-Sepúlveda, Julian Naizaque, Gerardo Santamaría, Magdalena Wiesner, Martha Lucia Ospina Martínez, Marcela Mercado-Reyes. |
| EPI_ISL_1138414 | Laboratorio de Referencia Nacional de Virus Respiratorio. Instituto Nacional de Salud Perú | Laboratorio de Referencia Nacional de Biotecnología y Biología Molecular. Instituto Nacional de Salud Perú | Carlos Padilla Rojas, Karolyn Vega Chozo, Luis Barcena, Priscila Lope Pari, Omar Caceres Rey, Marco Galarza Perez, Maribel Huaringa Nuñez, Johanna Balbuena Torrez, Henri Bailon Calderon, Nancy Rojas Serrano |
| EPI_ISL_1139069 | SMS Aruja | Instituto Adolfo Lutz, Interdiciplinary Procedures Center, Strategic Laboratory | Claudio Tavares Sacchi, Claudia Regina Gonçalves, Erica Valessa Ramos Gomes, Karoline Rodrigues Campos, Caio Vinicius Dias Lopes |
| EPI_ISL_1139070 | IAL Regional de Ribeirao Preto | Instituto Adolfo Lutz, Interdiciplinary Procedures Center, Strategic Laboratory | Claudio Tavares Sacchi, Claudia Regina Gonçalves, Erica Valessa Ramos Gomes, Karoline Rodrigues Campos, Caio Vinicius Dias Lopes |
| EPI_ISL_1139071, EPI_ISL_1139072, EPI_ISL_1139073, EPI_ISL_1139074 | Instituto Adolfo Lutz Central | Instituto Adolfo Lutz, Interdiciplinary Procedures Center, Strategic Laboratory | Claudio Tavares Sacchi, Claudia Regina Gonçalves, Erica Valessa Ramos Gomes, Karoline Rodrigues Campos, Caio Vinicius Dias Lopes |
| EPI_ISL_1139075, EPI_ISL_1139077 | Lab Loc - Itapecerica da Serra | Instituto Adolfo Lutz, Interdiciplinary Procedures Center, Strategic Laboratory | Claudio Tavares Sacchi, Claudia Regina Gonçalves, Erica Valessa Ramos Gomes, Karoline Rodrigues Campos, Caio Vinicius Dias Lopes |
| EPI_ISL_1161402, EPI_ISL_1161403, EPI_ISL_1161404, EPI_ISL_1161406, EPI_ISL_1161407, EPI_ISL_1161408, EPI_ISL_1161409, EPI_ISL_1161411, EPI_ISL_1163532, EPI_ISL_1163714 | LABCOVID_HCPA | LABRESIS_HCPA | Martins AF, Wink PL, Volpato F, Rosset C, de Paris F, Monteiro F, Zavascki AP, Barth AL |
| EPI_ISL_1164970, EPI_ISL_1164971 | LACEN - Laboratório Central de Saúde Pública do Ceará | Evandro Chagas Institute | Santos, M.C.; Silva, A.M.; Junior, W.D.C.; Barbagelata, L.S.; Ferreira, J.A.; Sousa, E.M.A.; da Silva, P.S.; Pinheiro, K.C.; L.C.; Sousa Junior, E.C. |
| EPI_ISL_1164972 | LACEN - Laboratório Central de Saúde Pública do Pará | Evandro Chagas Institute | Santos, M.C.; Silva, A.M.; Junior, W.D.C.; Barbagelata, L.S.; Ferreira, J.A.; Sousa, E.M.A.; da Silva, P.S.; Pinheiro, K.C.; L.C.; Sousa Junior, E.C. |
| EPI_ISL_1164973 | LACEN - Laboratório Central de Saúde Pública do Ceará | Evandro Chagas Institute | Santos, M.C.; Silva, A.M.; Junior, W.D.C.; Barbagelata, L.S.; Ferreira, J.A.; Sousa, E.M.A.; da Silva, P.S.; Pinheiro, K.C.; L.C.; Sousa Junior, E.C. |
| EPI_ISL_1164974, EPI_ISL_1164975 | LACEN - Laboratório Central de Saúde Pública do Pará | Evandro Chagas Institute | Santos, M.C.; Silva, A.M.; Junior, W.D.C.; Barbagelata, L.S.; Ferreira, J.A.; Sousa, E.M.A.; da Silva, P.S.; Pinheiro, K.C.; L.C.; Sousa Junior, E.C. |
| EPI_ISL_1164976 | LACEN - Laboratório Central de Saúde Pública do Amapá | Evandro Chagas Institute | Santos, M.C.; Silva, A.M.; Junior, W.D.C.; Barbagelata, L.S.; Ferreira, J.A.; Sousa, E.M.A.; da Silva, P.S.; Pinheiro, K.C.; L.C.; Sousa Junior, E.C. |
| EPI_ISL_1164978 | LACEN - Laboratório Central de Saúde Pública do Pará | Evandro Chagas Institute | Santos, M.C.; Silva, A.M.; Junior, W.D.C.; Barbagelata, L.S.; Ferreira, J.A.; Sousa, E.M.A.; da Silva, P.S.; Pinheiro, K.C.; L.C.; Sousa Junior, E.C. |
| EPI_ISL_1164979 | LACEN - Laboratório Central de Saúde Pública do Maranhao | Evandro Chagas Institute | Santos, M.C.; Silva, A.M.; Junior, W.D.C.; Barbagelata, L.S.; Ferreira, J.A.; Sousa, E.M.A.; da Silva, P.S.; Pinheiro, K.C.; L.C.; Sousa Junior, E.C. |
| EPI_ISL_1164980 | LACEN - Laboratório Central de Saúde Pública do Ceará | Evandro Chagas Institute | Santos, M.C.; Silva, A.M.; Junior, W.D.C.; Barbagelata, L.S.; Ferreira, J.A.; Sousa, E.M.A.; da Silva, P.S.; Pinheiro, K.C.; L.C.; Sousa Junior, E.C. |
| EPI_ISL_1164981, EPI_ISL_1164982 | LACEN - Laboratório Central de Saúde Pública do Amapá | Evandro Chagas Institute | Santos, M.C.; Silva, A.M.; Junior, W.D.C.; Barbagelata, L.S.; Ferreira, J.A.; Sousa, E.M.A.; da Silva, P.S.; Pinheiro, K.C.; L.C.; Sousa Junior, E.C. |
| EPI_ISL_1164983 | LACEN - Laboratório Central de Saúde Pública do Pará | Evandro Chagas Institute | Santos, M.C.; Silva, A.M.; Junior, W.D.C.; Barbagelata, L.S.; Ferreira, J.A.; Sousa, E.M.A.; da Silva, P.S.; Pinheiro, K.C.; L.C.; Sousa Junior, E.C. |
| EPI_ISL_1164984, EPI_ISL_1164985 | LACEN - Laboratório Central de Saúde Pública do Amapá | Evandro Chagas Institute | Santos, M.C.; Silva, A.M.; Junior, W.D.C.; Barbagelata, L.S.; Ferreira, J.A.; Sousa, E.M.A.; da Silva, P.S.; Pinheiro, K.C.; L.C.; Sousa Junior, E.C. |
| EPI_ISL_1164986 | LACEN - Laboratório Central de Saúde Pública do Ceará | Evandro Chagas Institute | Santos, M.C.; Silva, A.M.; Junior, W.D.C.; Barbagelata, L.S.; Ferreira, J.A.; Sousa, E.M.A.; da Silva, P.S.; Pinheiro, K.C.; L.C.; Sousa Junior, E.C. |
| EPI_ISL_1164987, EPI_ISL_1164988 | LACEN - Laboratório Central de Saúde Pública do Rio Grande do Norte | Evandro Chagas Institute | Santos, M.C.; Silva, A.M.; Junior, W.D.C.; Barbagelata, L.S.; Ferreira, J.A.; Sousa, E.M.A.; da Silva, P.S.; Pinheiro, K.C.; L.C.; Sousa Junior, E.C. |
| EPI_ISL_1164989, EPI_ISL_1164991, EPI_ISL_1164992 | LACEN - Laboratório Central de Saúde Pública do Paraiba | Evandro Chagas Institute | Santos, M.C.; Silva, A.M.; Junior, W.D.C.; Barbagelata, L.S.; Ferreira, J.A.; Sousa, E.M.A.; da Silva, P.S.; Pinheiro, K.C.; L.C.; Sousa Junior, E.C. |
| EPI_ISL_1164993 | LACEN - Laboratório Central de Saúde Pública do Ceará | Evandro Chagas Institute | Santos, M.C.; Silva, A.M.; Junior, W.D.C.; Barbagelata, L.S.; Ferreira, J.A.; Sousa, E.M.A.; da Silva, P.S.; Pinheiro, K.C.; L.C.; Sousa Junior, E.C. |
| EPI_ISL_1164996, EPI_ISL_1164998 | LACEN - Laboratório Central de Saúde Pública do Maranhao | Evandro Chagas Institute | Santos, M.C.; Silva, A.M.; Junior, W.D.C.; Barbagelata, L.S.; Ferreira, J.A.; Sousa, E.M.A.; da Silva, P.S.; Pinheiro, K.C.; L.C.; Sousa Junior, E.C. |
| EPI_ISL_1166615 | LACEN - Laboratório Central de Saúde Pública do Rio Grande do Norte | Evandro Chagas Institute Virology | Santos, M.C.; Silva, A.M.; Junior, W.D.C.; Barbagelata, L.S.; Ferreira, J.A.; Sousa, E.M.A.; da Silva, P.S.; Pinheiro, K.C.; L.C.; Sousa Junior, E.C. |
| EPI_ISL_1167743, EPI_ISL_1167859, EPI_ISL_1167887, EPI_ISL_1167899, EPI_ISL_1167912, EPI_ISL_1167917, | Genetica Molecular and Subdepartamento de Virologia ISP Chile | Instituto de Salud Publica de Chile | Javier Tognarelli, Karen Orostica, Barbara Parra, Loredana Arata, Jaime Lagos, Gisselle Barra, Patricia Bustos, Rodrigo Fasce, Andres Castillo, Jorge Fernandez |

| | | | |
|---|---|---|---|
| EPI_ISL_1167918, EPI_ISL_1167929 | | | |
| EPI_ISL_1171619 | IAL Regional de Presidente Prudente | Instituto Adolfo Lutz, Interdiciplinary Procedures Center, Strategic Laboratory | Claudio Tavares Sacchi, Claudia Regina Gonçalves, Erica Valessa Ramos Gomes, Karoline Rodrigues Campos |
| EPI_ISL_1171622, EPI_ISL_1171626, EPI_ISL_1171628, EPI_ISL_1171629, EPI_ISL_1171630, EPI_ISL_1171634, EPI_ISL_1171636, EPI_ISL_1171637, EPI_ISL_1171638, EPI_ISL_1171639, EPI_ISL_1171640 | | | |
| see above | IAL Regional de Santos | Instituto Adolfo Lutz, Interdiciplinary Procedures Center, Strategic Laboratory | Claudio Tavares Sacchi, Claudia Regina Gonçalves, Erica Valessa Ramos Gomes, Karoline Rodrigues Campos, Caio Vinicius Dias Lopes |
| EPI_ISL_1171641, EPI_ISL_1171642, EPI_ISL_1171643, EPI_ISL_1171644, EPI_ISL_1171645 | IAL Regional de Marilia | Instituto Adolfo Lutz, Interdiciplinary Procedures Center, Strategic Laboratory | Claudio Tavares Sacchi, Claudia Regina Gonçalves, Erica Valessa Ramos Gomes, Karoline Rodrigues Campos, Caio Vinicius Dias Lopes |
| EPI_ISL_1171648, EPI_ISL_1171649, EPI_ISL_1171650 | Centro de Saude Dr. Jose Paione em Mococa | Instituto Adolfo Lutz, Interdiciplinary Procedures Center, Strategic Laboratory | Claudio Tavares Sacchi, Claudia Regina Gonçalves, Erica Valessa Ramos Gomes, Karoline Rodrigues Campos, Caio Vinicius Dias Lopes |
| EPI_ISL_1171651, EPI_ISL_1171652, EPI_ISL_1171653, EPI_ISL_1171654, EPI_ISL_1171655, EPI_ISL_1171656, EPI_ISL_1171657, EPI_ISL_1171658, EPI_ISL_1171659, EPI_ISL_1171660, EPI_ISL_1171661, EPI_ISL_1171662, EPI_ISL_1171663, EPI_ISL_1171665, EPI_ISL_1171666, EPI_ISL_1171667, EPI_ISL_1171670, EPI_ISL_1171672, EPI_ISL_1171673, EPI_ISL_1171674 | | | |
| see above | IAL Regional de Presidente Prudente | Instituto Adolfo Lutz, Interdiciplinary Procedures Center, Strategic Laboratory | Claudio Tavares Sacchi, Claudia Regina Gonçalves, Erica Valessa Ramos Gomes, Karoline Rodrigues Campos, Caio Vinicius Dias Lopes |
| EPI_ISL_1181362 | Instituto Aggeu Magalhães - Oswaldo Cruz Foundation, FIOCRUZ (FIOCRUZ-PE) | Laboratory of Respiratory Viruses and Measles, Oswaldo Cruz Institute, FIOCRUZ | Paola Resende, Luciana Appolinario, Fernando Motta, Anna Carolina Paixao, Ana Carolina Mendonca, Alice Sampaio Rocha, Renata Serrano Lopes, Gabriel Wallau, Marilda Siqueira on behalf of the Fiocruz COVID-19 Genomic Surveillance Network |
| EPI_ISL_1181370, EPI_ISL_1181371 | Laboratorio Central de Saude Publica do Estado Maranhao (LACEN-MA) | Laboratory of Respiratory Viruses and Measles, Oswaldo Cruz Institute, FIOCRUZ | Paola Resende, Luciana Appolinario, Fernando Motta, Anna Carolina Paixao, Ana Carolina Mendonca, Alice Sampaio Rocha, Renata Serrano Lopes, Lidio Gonçalves Lima Neto, Marilda Siqueira on behalf of the Fiocruz COVID-19 Genomic Surveillance Network |
| EPI_ISL_1181380 | Gonçalo Moniz Institute, FIOCRUZ, Bahia | Laboratory of Respiratory Viruses and Measles, Oswaldo Cruz Institute, FIOCRUZ | Paola Resende, Luciana Appolinario, Fernando Motta, Anna Carolina Paixao, Ana Carolina Mendonca, Alice Sampaio Rocha, Renata Serrano Lopes, Tiago Graf, Ricardo Khouri, Marilda Siqueira on behalf of the Fiocruz COVID-19 Genomic Surveillance Network |
| EPI_ISL_1181400, EPI_ISL_1181401 | Laboratorio Central de Saude Publica do Estado Maranhao (LACEN-MA) | Laboratory of Respiratory Viruses and Measles, Oswaldo Cruz Institute, FIOCRUZ | Paola Resende, Luciana Appolinario, Fernando Motta, Anna Carolina Paixao, Ana Carolina Mendonca, Alice Sampaio Rocha, Renata Serrano Lopes, Lidio Gonçalves Lima Neto, Marilda Siqueira on behalf of the Fiocruz COVID-19 Genomic Surveillance Network |
| EPI_ISL_1181402 | Laboratory of Respiratory Viruses and Measles, Oswaldo Cruz Institute, FIOCRUZ | Laboratory of Respiratory Viruses and Measles, Oswaldo Cruz Institute, FIOCRUZ | Paola Resende, Luciana Appolinario, Fernando Motta, Anna Carolina Paixao, Ana Carolina Mendonca, Alice Sampaio Rocha, Renata Serrano Lopes, Marilda Siqueira on behalf of the Fiocruz COVID-19 Genomic Surveillance Network |
| EPI_ISL_1181403 | Laboratorio Central de Saude Publica do Estado de Santa Catarina (LACEN-SC) | Laboratory of Respiratory Viruses and Measles, Oswaldo Cruz Institute, FIOCRUZ | Paola Resende, Luciana Appolinario, Fernando Motta, Anna Carolina Paixao, Ana Carolina Mendonca, Alice Sampaio Rocha, Renata Serrano Lopes, Darcita Buerger Rovaris, Sandra Bianchini Fernandes, Marilda Siqueira on behalf of the Fiocruz COVID-19 Genomic Surveillance Network |
| EPI_ISL_1181407 | Laboratorio Central de Saude Publica do Estado de Sergipe (LACEN-SE) | Laboratory of Respiratory Viruses and Measles, Oswaldo Cruz Institute, FIOCRUZ | Paola Resende, Luciana Appolinario, Fernando Motta, Anna Carolina Paixao, Ana Carolina Mendonca, Alice Sampaio Rocha, Renata Serrano Lopes, Cliomar Alves dos Santos, Marilda Siqueira on behalf of the Fiocruz COVID-19 Genomic Surveillance Network |
| EPI_ISL_1181408 | Gonçalo Moniz Institute, FIOCRUZ, Bahia | Laboratory of Respiratory Viruses and Measles, Oswaldo Cruz Institute, FIOCRUZ | Paola Resende, Luciana Appolinario, Fernando Motta, Anna Carolina Paixao, Ana Carolina Mendonca, Alice Sampaio Rocha, Renata Serrano Lopes, Tiago Graf, Ricardo Khouri, Marilda Siqueira on behalf of the Fiocruz COVID-19 Genomic Surveillance Network |
| EPI_ISL_1181409 | Laboratorio Central de Saude Publica do Estado de Sergipe (LACEN-SE) | Laboratory of Respiratory Viruses and Measles, Oswaldo Cruz Institute, FIOCRUZ | Paola Resende, Luciana Appolinario, Fernando Motta, Anna Carolina Paixao, Ana Carolina Mendonca, Alice Sampaio Rocha, Renata Serrano Lopes, Cliomar Alves dos Santos, Marilda Siqueira on behalf of the Fiocruz COVID-19 Genomic Surveillance Network |
| EPI_ISL_1181413, EPI_ISL_1181414 | Laboratorio Central de Saude Publica do Estado Maranhao (LACEN-MA) | Laboratory of Respiratory Viruses and Measles, Oswaldo Cruz Institute, FIOCRUZ | Paola Resende, Luciana Appolinario, Fernando Motta, Anna Carolina Paixao, Ana Carolina Mendonca, Alice Sampaio Rocha, Renata Serrano Lopes, Lidio Gonçalves Lima Neto, Marilda Siqueira on behalf of the Fiocruz COVID-19 Genomic Surveillance Network |
| EPI_ISL_1181416 | Laboratorio Central de Saude Publica do Estado da Paraiba (LACEN-PB) | Laboratory of Respiratory Viruses and Measles, Oswaldo Cruz Institute, FIOCRUZ | Paola Resende, Luciana Appolinario, Fernando Motta, Anna Carolina Paixao, Ana Carolina Mendonca, Alice Sampaio Rocha, Renata Serrano Lopes, Joao Felipe Bezerra, Dalane Loudal Florentino Teixeira, Marilda Siqueira on behalf of the Fiocruz COVID-19 Genomic Surveillance Network |
| EPI_ISL_1181419 | Gonçalo Moniz Institute, FIOCRUZ, Bahia | Laboratory of Respiratory Viruses and Measles, Oswaldo Cruz Institute, FIOCRUZ | Paola Resende, Luciana Appolinario, Fernando Motta, Anna Carolina Paixao, Ana Carolina Mendonca, Alice Sampaio Rocha, Renata Serrano Lopes, Tiago Graf, Ricardo Khouri, Marilda Siqueira on behalf of the Fiocruz COVID-19 Genomic Surveillance Network |
| EPI_ISL_1182096, EPI_ISL_1182098, EPI_ISL_1182099, EPI_ISL_1182100, EPI_ISL_1182101, EPI_ISL_1182102 | Adolfo Lutz Institute Santos Regional Center Brazil | Retrovirus Laboratory Adolfo Lutz Institute | Gabriela Bastos Cabral, Giselle I S Lopez-Lopes, Cintia Ahagon, Audrey Cilli, Paula Morena Guimaraes, Igor Mohamed Hussein, Luis Brigido |
| EPI_ISL_1182541, EPI_ISL_1182543, EPI_ISL_1182544, EPI_ISL_1182545 | Fundação Ezequiel Dias (FUNED) | Coordenação Geral de Laboratórios de Saúde Pública (CGLAB/DAEVS/SVS/MS) | Vagner Fonseca, et al. |
| EPI_ISL_1182546 | Laboratório Central do Estado do Paraná | Coordenação Geral de Laboratórios de Saúde Pública (CGLAB/DAEVS/SVS/MS) | Vagner Fonseca, et al. |
| EPI_ISL_1182551 | Fundação Ezequiel Dias (FUNED) | Coordenação Geral de Laboratórios de Saúde Pública (CGLAB/DAEVS/SVS/MS) | Vagner Fonseca, et al. |
| EPI_ISL_1182552, EPI_ISL_1182553 | Laboratório Central do Estado do Rio de Janeiro | Coordenação Geral de Laboratórios de Saúde Pública (CGLAB/DAEVS/SVS/MS) | Vagner Fonseca, et al. |
| EPI_ISL_1182555 | Fundação Ezequiel Dias (FUNED) | Coordenação Geral de Laboratórios de Saúde Pública (CGLAB/DAEVS/SVS/MS) | Vagner Fonseca, et al. |
| EPI_ISL_1182556, EPI_ISL_1182557, EPI_ISL_1182558 | Laboratório Central do Estado do Rio de Janeiro | Coordenação Geral de Laboratórios de Saúde Pública (CGLAB/DAEVS/SVS/MS) | Vagner Fonseca, et al. |
| EPI_ISL_1182559, EPI_ISL_1182560, EPI_ISL_1182561 | Fundação Ezequiel Dias (FUNED) | Coordenação Geral de Laboratórios de Saúde Pública (CGLAB/DAEVS/SVS/MS) | Vagner Fonseca, et al. |
| EPI_ISL_1182563, EPI_ISL_1182565 | Laboratório Central do Estado do Paraná | Coordenação Geral de Laboratórios de Saúde Pública (CGLAB/DAEVS/SVS/MS) | Vagner Fonseca, et al. |
| EPI_ISL_1182566 | Fundação Ezequiel Dias (FUNED) | Coordenação Geral de Laboratórios de Saúde Pública (CGLAB/DAEVS/SVS/MS) | Vagner Fonseca, et al. |
| EPI_ISL_1182568 | Laboratório Central do Estado do Paraná | Coordenação Geral de Laboratórios de Saúde Pública (CGLAB/DAEVS/SVS/MS) | Vagner Fonseca, et al. |
| EPI_ISL_1182569, EPI_ISL_1182570 | Fundação Ezequiel Dias (FUNED) | Coordenação Geral de Laboratórios de Saúde Pública (CGLAB/DAEVS/SVS/MS) | Vagner Fonseca, et al. |
| EPI_ISL_1182571 | Laboratório Central do Estado do Paraná | Coordenação Geral de Laboratórios de Saúde Pública (CGLAB/DAEVS/SVS/MS) | Vagner Fonseca, et al. |
| EPI_ISL_1182573, EPI_ISL_1182574 | Fundação Ezequiel Dias (FUNED) | Coordenação Geral de Laboratórios de Saúde Pública (CGLAB/DAEVS/SVS/MS) | Vagner Fonseca, et al. |
| EPI_ISL_1182575 | Laboratório Central do Estado do Paraná | Coordenação Geral de Laboratórios de Saúde Pública (CGLAB/DAEVS/SVS/MS) | Vagner Fonseca, et al. |

| | | | |
|---|---|---|---|
| EPI_ISL_1182577, EPI_ISL_1182578, EPI_ISL_1182579 | Fundação Ezequiel Dias (FUNED) | Coordenação Geral de Laboratórios de Saúde Pública (CGLAB/DAEVS/SVS/MS) | Vagner Fonseca, et al. |
| EPI_ISL_1182580, EPI_ISL_1182581, EPI_ISL_1182582, EPI_ISL_1182583 | Laboratório Central do Estado do Paraná | Coordenação Geral de Laboratórios de Saúde Pública (CGLAB/DAEVS/SVS/MS) | Vagner Fonseca, et al. |
| EPI_ISL_1182585, EPI_ISL_1182586 | Fundação Ezequiel Dias (FUNED) | Coordenação Geral de Laboratórios de Saúde Pública (CGLAB/DAEVS/SVS/MS) | Vagner Fonseca, et al. |
| EPI_ISL_1182588, EPI_ISL_1182589 | Laboratório Central do Estado do Paraná | Coordenação Geral de Laboratórios de Saúde Pública (CGLAB/DAEVS/SVS/MS) | Vagner Fonseca, et al. |
| EPI_ISL_1182590, EPI_ISL_1182591 | Fundação Ezequiel Dias (FUNED) | Coordenação Geral de Laboratórios de Saúde Pública (CGLAB/DAEVS/SVS/MS) | Vagner Fonseca, et al. |
| EPI_ISL_1182592 | Laboratório Central do Estado do Paraná | Coordenação Geral de Laboratórios de Saúde Pública (CGLAB/DAEVS/SVS/MS) | Vagner Fonseca, et al. |
| EPI_ISL_1182593 | Fundação Ezequiel Dias (FUNED) | Coordenação Geral de Laboratórios de Saúde Pública (CGLAB/DAEVS/SVS/MS) | Vagner Fonseca, et al. |
| EPI_ISL_1182594, EPI_ISL_1182596 | Laboratório Central do Estado do Paraná | Coordenação Geral de Laboratórios de Saúde Pública (CGLAB/DAEVS/SVS/MS) | Vagner Fonseca, et al. |
| EPI_ISL_1182597 | Laboratório Central de Saúde Pública do Rio Grande do Sul | Coordenação Geral de Laboratórios de Saúde Pública (CGLAB/DAEVS/SVS/MS) | Vagner Fonseca, et al. |
| EPI_ISL_1182598, EPI_ISL_1182600, EPI_ISL_1182602 | Fundação Ezequiel Dias (FUNED) | Coordenação Geral de Laboratórios de Saúde Pública (CGLAB/DAEVS/SVS/MS) | Vagner Fonseca, et al. |
| EPI_ISL_1182604 | Laboratório Central do Estado do Paraná | Coordenação Geral de Laboratórios de Saúde Pública (CGLAB/DAEVS/SVS/MS) | Vagner Fonseca, et al. |
| EPI_ISL_1182605, EPI_ISL_1182606 | Laboratório Central de Saúde Pública do Rio Grande do Sul | Coordenação Geral de Laboratórios de Saúde Pública (CGLAB/DAEVS/SVS/MS) | Vagner Fonseca, et al. |
| EPI_ISL_1182611, EPI_ISL_1182615 | Fundação Ezequiel Dias (FUNED) | Coordenação Geral de Laboratórios de Saúde Pública (CGLAB/DAEVS/SVS/MS) | Vagner Fonseca, et al. |
| EPI_ISL_1182616 | Laboratório Central de Saúde Pública do Rio Grande do Sul | Coordenação Geral de Laboratórios de Saúde Pública (CGLAB/DAEVS/SVS/MS) | Vagner Fonseca, et al. |
| EPI_ISL_1182617 | Laboratório Central do Estado do Paraná | Coordenação Geral de Laboratórios de Saúde Pública (CGLAB/DAEVS/SVS/MS) | Vagner Fonseca, et al. |
| EPI_ISL_1182618 | Fundação Ezequiel Dias (FUNED) | Coordenação Geral de Laboratórios de Saúde Pública (CGLAB/DAEVS/SVS/MS) | Vagner Fonseca, et al. |
| EPI_ISL_1182619, EPI_ISL_1182620 | Laboratório Central de Saúde Pública do Rio Grande do Sul | Coordenação Geral de Laboratórios de Saúde Pública (CGLAB/DAEVS/SVS/MS) | Vagner Fonseca, et al. |
| EPI_ISL_1182622 | Laboratório Central do Estado do Paraná | Coordenação Geral de Laboratórios de Saúde Pública (CGLAB/DAEVS/SVS/MS) | Vagner Fonseca, et al. |
| EPI_ISL_1182625 | Fundação Ezequiel Dias (FUNED) | Coordenação Geral de Laboratórios de Saúde Pública (CGLAB/DAEVS/SVS/MS) | Vagner Fonseca, et al. |
| EPI_ISL_1196289, EPI_ISL_1196290, EPI_ISL_1196292, EPI_ISL_1196294 | LACEN do Distrito Federal | Instituto Adolfo Lutz, Interdiciplinary Procedures Center, Strategic Laboratory | Claudio Tavares Sacchi, Claudia Regina Gonçalves, Erica Valessa Ramos Gomes, Karoline Rodrigues Campos, Caio Vinicius Dias Lopes |
| EPI_ISL_1196295 | UBS Otacilio Firmino Lopes | Instituto Adolfo Lutz, Interdiciplinary Procedures Center, Strategic Laboratory | Claudio Tavares Sacchi, Claudia Regina Gonçalves, Erica Valessa Ramos Gomes, Karoline Rodrigues Campos, Caio Vinicius Dias Lopes |
| EPI_ISL_1196296 | Centro de Saude II Dr. Jose Paione Mococa | Instituto Adolfo Lutz, Interdiciplinary Procedures Center, Strategic Laboratory | Claudio Tavares Sacchi, Claudia Regina Gonçalves, Erica Valessa Ramos Gomes, Karoline Rodrigues Campos, Caio Vinicius Dias Lopes |
| EPI_ISL_1196299, EPI_ISL_1196300, EPI_ISL_1196302 | IAL Regional de Marilia | Instituto Adolfo Lutz, Interdiciplinary Procedures Center, Strategic Laboratory | Claudio Tavares Sacchi, Claudia Regina Gonçalves, Erica Valessa Ramos Gomes, Karoline Rodrigues Campos, Caio Vinicius Dias Lopes |
| EPI_ISL_1201526, EPI_ISL_1201527, EPI_ISL_1201528, EPI_ISL_1201530, EPI_ISL_1201532 | Outre Mer | National Reference Center for Viruses of Respiratory Infections, Institut Pasteur, Paris | Marion Barbet, Sylvie Behillil, Méline Bizard, Angela Brisebarre, Camille Capel, Etienne Simon-Lorière, Vincent Enouf, Maud Vanpeene, Sylvie van der Werf,Rousset Dominique |
| EPI_ISL_1201884, EPI_ISL_1201885, EPI_ISL_1201887 | Aeroporto Internacional de Guarulhos | Instituto Adolfo Lutz, Interdiciplinary Procedures Center, Strategic Laboratory | Claudio Tavares Sacchi, Claudia Regina Gonçalves, Erica Valessa Ramos Gomes, Karoline Rodrigues Campos, Caio Vinicius Dias Lopes |
| EPI_ISL_1201890, EPI_ISL_1201891, EPI_ISL_1201892 | IAL Regional de Marilia | Instituto Adolfo Lutz, Interdiciplinary Procedures Center, Strategic Laboratory | Claudio Tavares Sacchi, Claudia Regina Gonçalves, Erica Valessa Ramos Gomes, Karoline Rodrigues Campos, Caio Vinicius Dias Lopes |
| EPI_ISL_1201893 | IAL Regional de Sorocaba | Instituto Adolfo Lutz, Interdiciplinary Procedures Center, Strategic Laboratory | Claudio Tavares Sacchi, Claudia Regina Gonçalves, Erica Valessa Ramos Gomes, Karoline Rodrigues Campos, Caio Vinicius Dias Lopes |
| EPI_ISL_1201897 | Fundación Cardio Infantil | Instituto Nacional de Salud- Dirección de Investigación en Salud Pública | Katherine Laiton-Donato, Carlos Franco-Muñoz, Diego A. Álvarez-Díaz, Hector Alejandro Ruiz-Moreno, Jhonnatan Reales-González, Diego Andrés Prada, Sheryll Corchuelo, Maria T. Herrera-Sepúlveda, Julian Naizaque, Gerardo Santamaría, Magdalena Wiesner, Martha Lucia Ospina Martínez, Marcela Mercado-Reyes. |
| EPI_ISL_1213148, EPI_ISL_1213149, EPI_ISL_1213151 | Laboratório HLA/UERJ | Bioinformatics Laboratory / LNCC | Alessandra P Lamarca, Luiz G P de Almeida, Ronaldo da Silva Francisco Jr, Lucymara Fassarella Agnez Lima, Kátia Castanho Scortecci, Vinícius Pietta Perez, Otavio J. Brustolini, Eduardo Sérgio Soares Sousa, Danielle Angst Secco, Angela Maria Guimarães Santos, George Rego Albuquerque, Ana Paula Melo Mariano, Bianca Mendes Maciel, Alexandra L Gerber, Ana Paula de C Guimarães, Paulo Ricardo Nascimento, Francisco Paulo Freire Neto, Sandra Rocha Gadelha, Luís Cristóvão Porto, Eloiza Helena Campana, Selma Maria Bezerra Jeronimo, Ana Tereza R Vasconcelos |
| EPI_ISL_1213153 | LBM/UFPB | Bioinformatics Laboratory / LNCC | Alessandra P Lamarca, Luiz G P de Almeida, Ronaldo da Silva Francisco Jr, Lucymara Fassarella Agnez Lima, Kátia Castanho Scortecci, Vinícius Pietta Perez, Otavio J. Brustolini, Eduardo Sérgio Soares Sousa, Danielle Angst Secco, Angela Maria Guimarães Santos, George Rego Albuquerque, Ana Paula Melo Mariano, Bianca Mendes Maciel, Alexandra L Gerber, Ana Paula de C Guimarães, Paulo Ricardo Nascimento, Francisco Paulo Freire Neto, Sandra Rocha Gadelha, Luís Cristóvão Porto, Eloiza Helena Campana, Selma Maria Bezerra Jeronimo, Ana Tereza R Vasconcelos |
| EPI_ISL_1213155, EPI_ISL_1213158 | Laboratório HLA/UERJ | Bioinformatics Laboratory / LNCC | Alessandra P Lamarca, Luiz G P de Almeida, Ronaldo da Silva Francisco Jr, Lucymara Fassarella Agnez Lima, Kátia Castanho Scortecci, Vinícius Pietta Perez, Otavio J. Brustolini, Eduardo Sérgio Soares Sousa, Danielle Angst Secco, Angela Maria Guimarães Santos, George Rego Albuquerque, Ana Paula Melo Mariano, Bianca Mendes Maciel, Alexandra L Gerber, Ana Paula de C Guimarães, Paulo Ricardo Nascimento, Francisco Paulo Freire Neto, Sandra Rocha Gadelha, Luís Cristóvão Porto, Eloiza Helena Campana, Selma Maria Bezerra Jeronimo, Ana Tereza R Vasconcelos |
| EPI_ISL_1213161 | LBM/UFPB | Bioinformatics Laboratory / LNCC | Alessandra P Lamarca, Luiz G P de Almeida, Ronaldo da Silva Francisco Jr, Lucymara Fassarella Agnez Lima, Kátia Castanho Scortecci, Vinícius Pietta Perez, Otavio J. Brustolini, Eduardo Sérgio Soares Sousa, Danielle Angst Secco, Angela Maria Guimarães Santos, George Rego Albuquerque, Ana Paula |

| | | | |
|---|---|---|---|
| | | | Melo Mariano, Bianca Mendes Maciel, Alexandra L Gerber, Ana Paula de C Guimarães, Paulo Ricardo Nascimento, Francisco Paulo Freire Neto, Sandra Rocha Gadelha, Luís Cristóvão Porto, Eloiza Helena Campana, Selma Maria Bezerra Jeronimo, Ana Tereza R Vasconcelos |
| EPI_ISL_1213163 | Laboratório HLA/UERJ | Bioinformatics Laboratory / LNCC | Alessandra P Lamarca, Luiz G P de Almeida, Ronaldo da Silva Francisco Jr, Lucymara Fassarella Agnez Lima, Kátia Castanho Scortecci, Vinícius Pietta Perez, Otavio J. Brustolini, Eduardo Sérgio Soares Sousa, Danielle Angst Secco, Angela Maria Guimarães Santos, George Rego Albuquerque, Ana Paula Melo Mariano, Bianca Mendes Maciel, Alexandra L Gerber, Ana Paula de C Guimarães, Paulo Ricardo Nascimento, Francisco Paulo Freire Neto, Sandra Rocha Gadelha, Luís Cristóvão Porto, Eloiza Helena Campana, Selma Maria Bezerra Jeronimo, Ana Tereza R Vasconcelos |
| EPI_ISL_1213168, EPI_ISL_1213170 | LAFEM/UESC | Bioinformatics Laboratory / LNCC | Alessandra P Lamarca, Luiz G P de Almeida, Ronaldo da Silva Francisco Jr, Lucymara Fassarella Agnez Lima, Kátia Castanho Scortecci, Vinícius Pietta Perez, Otavio J. Brustolini, Eduardo Sérgio Soares Sousa, Danielle Angst Secco, Angela Maria Guimarães Santos, George Rego Albuquerque, Ana Paula Melo Mariano, Bianca Mendes Maciel, Alexandra L Gerber, Ana Paula de C Guimarães, Paulo Ricardo Nascimento, Francisco Paulo Freire Neto, Sandra Rocha Gadelha, Luís Cristóvão Porto, Eloiza Helena Campana, Selma Maria Bezerra Jeronimo, Ana Tereza R Vasconcelos |
| EPI_ISL_1213171 | Laboratório HLA/UERJ | Bioinformatics Laboratory / LNCC | Alessandra P Lamarca, Luiz G P de Almeida, Ronaldo da Silva Francisco Jr, Lucymara Fassarella Agnez Lima, Kátia Castanho Scortecci, Vinícius Pietta Perez, Otavio J. Brustolini, Eduardo Sérgio Soares Sousa, Danielle Angst Secco, Angela Maria Guimarães Santos, George Rego Albuquerque, Ana Paula Melo Mariano, Bianca Mendes Maciel, Alexandra L Gerber, Ana Paula de C Guimarães, Paulo Ricardo Nascimento, Francisco Paulo Freire Neto, Sandra Rocha Gadelha, Luís Cristóvão Porto, Eloiza Helena Campana, Selma Maria Bezerra Jeronimo, Ana Tereza R Vasconcelos |
| EPI_ISL_1213173, EPI_ISL_1213175, EPI_ISL_1213177, EPI_ISL_1213178, EPI_ISL_1213180, EPI_ISL_1213182, EPI_ISL_1213183, EPI_ISL_1213185, EPI_ISL_1213187 | IMT-UFRN/RN | Bioinformatics Laboratory / LNCC | Alessandra P Lamarca, Luiz G P de Almeida, Ronaldo da Silva Francisco Jr, Lucymara Fassarella Agnez Lima, Kátia Castanho Scortecci, Vinícius Pietta Perez, Otavio J. Brustolini, Eduardo Sérgio Soares Sousa, Danielle Angst Secco, Angela Maria Guimarães Santos, George Rego Albuquerque, Ana Paula Melo Mariano, Bianca Mendes Maciel, Alexandra L Gerber, Ana Paula de C Guimarães, Paulo Ricardo Nascimento, Francisco Paulo Freire Neto, Sandra Rocha Gadelha, Luís Cristóvão Porto, Eloiza Helena Campana, Selma Maria Bezerra Jeronimo, Ana Tereza R Vasconcelos |
| EPI_ISL_1213189, EPI_ISL_1213190 | LBM/UFPB | Bioinformatics Laboratory / LNCC | Alessandra P Lamarca, Luiz G P de Almeida, Ronaldo da Silva Francisco Jr, Lucymara Fassarella Agnez Lima, Kátia Castanho Scortecci, Vinícius Pietta Perez, Otavio J. Brustolini, Eduardo Sérgio Soares Sousa, Danielle Angst Secco, Angela Maria Guimarães Santos, George Rego Albuquerque, Ana Paula Melo Mariano, Bianca Mendes Maciel, Alexandra L Gerber, Ana Paula de C Guimarães, Paulo Ricardo Nascimento, Francisco Paulo Freire Neto, Sandra Rocha Gadelha, Luís Cristóvão Porto, Eloiza Helena Campana, Selma Maria Bezerra Jeronimo, Ana Tereza R Vasconcelos |
| EPI_ISL_1213192, EPI_ISL_1213194, EPI_ISL_1213196, EPI_ISL_1213197, EPI_ISL_1213199 | IMT-UFRN/RN | Bioinformatics Laboratory / LNCC | Alessandra P Lamarca, Luiz G P de Almeida, Ronaldo da Silva Francisco Jr, Lucymara Fassarella Agnez Lima, Kátia Castanho Scortecci, Vinícius Pietta Perez, Otavio J. Brustolini, Eduardo Sérgio Soares Sousa, Danielle Angst Secco, Angela Maria Guimarães Santos, George Rego Albuquerque, Ana Paula Melo Mariano, Bianca Mendes Maciel, Alexandra L Gerber, Ana Paula de C Guimarães, Paulo Ricardo Nascimento, Francisco Paulo Freire Neto, Sandra Rocha Gadelha, Luís Cristóvão Porto, Eloiza Helena Campana, Selma Maria Bezerra Jeronimo, Ana Tereza R Vasconcelos |
| EPI_ISL_1213201, EPI_ISL_1213202, EPI_ISL_1213204, EPI_ISL_1213206, EPI_ISL_1213207 | LBM/UFPB | Bioinformatics Laboratory / LNCC | Alessandra P Lamarca, Luiz G P de Almeida, Ronaldo da Silva Francisco Jr, Lucymara Fassarella Agnez Lima, Kátia Castanho Scortecci, Vinícius Pietta Perez, Otavio J. Brustolini, Eduardo Sérgio Soares Sousa, Danielle Angst Secco, Angela Maria Guimarães Santos, George Rego Albuquerque, Ana Paula Melo Mariano, Bianca Mendes Maciel, Alexandra L Gerber, Ana Paula de C Guimarães, Paulo Ricardo Nascimento, Francisco Paulo Freire Neto, Sandra Rocha Gadelha, Luís Cristóvão Porto, Eloiza Helena Campana, Selma Maria Bezerra Jeronimo, Ana Tereza R Vasconcelos |
| EPI_ISL_1213209 | IMT-UFRN/RN | Bioinformatics Laboratory / LNCC | Alessandra P Lamarca, Luiz G P de Almeida, Ronaldo da Silva Francisco Jr, Lucymara Fassarella Agnez Lima, Kátia Castanho Scortecci, Vinícius Pietta Perez, Otavio J. Brustolini, Eduardo Sérgio Soares Sousa, Danielle Angst Secco, Angela Maria Guimarães Santos, George Rego Albuquerque, Ana Paula Melo Mariano, Bianca Mendes Maciel, Alexandra L Gerber, Ana Paula de C Guimarães, Paulo Ricardo Nascimento, Francisco Paulo Freire Neto, Sandra Rocha Gadelha, Luís Cristóvão Porto, Eloiza Helena Campana, Selma Maria Bezerra Jeronimo, Ana Tereza R Vasconcelos |
| EPI_ISL_1213211 | LAFEM/UESC | Bioinformatics Laboratory / LNCC | Alessandra P Lamarca, Luiz G P de Almeida, Ronaldo da Silva Francisco Jr, Lucymara Fassarella Agnez Lima, Kátia Castanho Scortecci, Vinícius Pietta Perez, Otavio J. Brustolini, Eduardo Sérgio Soares Sousa, Danielle Angst Secco, Angela Maria Guimarães Santos, George Rego Albuquerque, Ana Paula Melo Mariano, Bianca Mendes Maciel, Alexandra L Gerber, Ana Paula de C Guimarães, Paulo Ricardo Nascimento, Francisco Paulo Freire Neto, Sandra Rocha Gadelha, Luís Cristóvão Porto, Eloiza Helena Campana, Selma Maria Bezerra Jeronimo, Ana Tereza R Vasconcelos |
| EPI_ISL_1213213, EPI_ISL_1213215 | LBM/UFPB | Bioinformatics Laboratory / LNCC | Alessandra P Lamarca, Luiz G P de Almeida, Ronaldo da Silva Francisco Jr, Lucymara Fassarella Agnez Lima, Kátia Castanho Scortecci, Vinícius Pietta Perez, Otavio J. Brustolini, Eduardo Sérgio Soares Sousa, Danielle Angst Secco, Angela Maria Guimarães Santos, George Rego Albuquerque, Ana Paula Melo Mariano, Bianca Mendes Maciel, Alexandra L Gerber, Ana Paula de C Guimarães, Paulo Ricardo Nascimento, Francisco Paulo Freire Neto, Sandra Rocha Gadelha, Luís Cristóvão Porto, Eloiza Helena Campana, Selma Maria Bezerra Jeronimo, Ana Tereza R Vasconcelos |
| EPI_ISL_1213216, EPI_ISL_1213218, EPI_ISL_1213222, EPI_ISL_1213224, EPI_ISL_1213227, EPI_ISL_1213229, EPI_ISL_1213230, EPI_ISL_1213232, EPI_ISL_1213234, EPI_ISL_1213236, EPI_ISL_1213239, EPI_ISL_1213241, EPI_ISL_1213242, EPI_ISL_1213244, EPI_ISL_1213245 | | | |
| see above | LAFEM/UESC | Bioinformatics Laboratory / LNCC | Alessandra P Lamarca, Luiz G P de Almeida, Ronaldo da Silva Francisco Jr, Lucymara Fassarella Agnez Lima, Kátia Castanho Scortecci, Vinícius Pietta Perez, Otavio J. Brustolini, Eduardo Sérgio Soares Sousa, Danielle Angst Secco, Angela Maria Guimarães Santos, George Rego Albuquerque, Ana Paula Melo Mariano, Bianca Mendes Maciel, Alexandra L Gerber, Ana Paula de C Guimarães, Paulo Ricardo Nascimento, Francisco Paulo Freire Neto, Sandra Rocha Gadelha, Luís Cristóvão Porto, Eloiza Helena Campana, Selma Maria Bezerra Jeronimo, Ana Tereza R Vasconcelos |
| EPI_ISL_1213248, EPI_ISL_1213249 | Laboratório HLA/UERJ | Bioinformatics Laboratory / LNCC | Alessandra P Lamarca, Luiz G P de Almeida, Ronaldo da Silva Francisco Jr, Lucymara Fassarella Agnez Lima, Kátia Castanho Scortecci, Vinícius Pietta Perez, Otavio J. Brustolini, Eduardo Sérgio Soares Sousa, Danielle Angst Secco, Angela Maria Guimarães Santos, George Rego Albuquerque, Ana Paula Melo Mariano, Bianca Mendes Maciel, Alexandra L Gerber, Ana Paula de C Guimarães, Paulo Ricardo Nascimento, Francisco Paulo Freire Neto, Sandra Rocha Gadelha, Luís Cristóvão Porto, Eloiza Helena Campana, Selma Maria Bezerra Jeronimo, Ana Tereza R Vasconcelos |
| EPI_ISL_1213251, EPI_ISL_1213252, EPI_ISL_1213254, EPI_ISL_1213256, EPI_ISL_1213258, EPI_ISL_1213261, EPI_ISL_1213263, EPI_ISL_1213265, EPI_ISL_1213267 | LAFEM/UESC | Bioinformatics Laboratory / LNCC | Alessandra P Lamarca, Luiz G P de Almeida, Ronaldo da Silva Francisco Jr, Lucymara Fassarella Agnez Lima, Kátia Castanho Scortecci, Vinícius Pietta Perez, Otavio J. Brustolini, Eduardo Sérgio Soares Sousa, Danielle Angst Secco, Angela Maria Guimarães Santos, George Rego Albuquerque, Ana Paula Melo Mariano, Bianca Mendes Maciel, Alexandra L Gerber, Ana Paula de C Guimarães, Paulo Ricardo Nascimento, Francisco Paulo Freire Neto, Sandra Rocha Gadelha, Luís Cristóvão Porto, Eloiza Helena Campana, Selma Maria Bezerra Jeronimo, Ana Tereza R Vasconcelos |
| EPI_ISL_1213272 | Laboratório HLA/UERJ | Bioinformatics Laboratory / LNCC | Alessandra P Lamarca, Luiz G P de Almeida, Ronaldo da Silva Francisco Jr, Lucymara Fassarella Agnez Lima, Kátia Castanho Scortecci, Vinícius Pietta Perez, Otavio J. Brustolini, Eduardo Sérgio Soares Sousa, Danielle Angst Secco, Angela Maria Guimarães Santos, George Rego Albuquerque, Ana Paula Melo Mariano, Bianca Mendes Maciel, Alexandra L Gerber, Ana Paula de C Guimarães, Paulo Ricardo Nascimento, Francisco Paulo Freire Neto, Sandra Rocha Gadelha, Luís Cristóvão Porto, Eloiza Helena Campana, Selma Maria Bezerra Jeronimo, Ana Tereza R Vasconcelos |
| EPI_ISL_1213274 | LAFEM/UESC | Bioinformatics Laboratory / LNCC | Alessandra P Lamarca, Luiz G P de Almeida, Ronaldo da Silva Francisco Jr, Lucymara Fassarella Agnez Lima, Kátia Castanho Scortecci, Vinícius Pietta Perez, Otavio J. Brustolini, Eduardo Sérgio Soares Sousa, Danielle Angst Secco, Angela Maria Guimarães Santos, George Rego Albuquerque, Ana Paula Melo Mariano, Bianca Mendes Maciel, Alexandra L Gerber, Ana Paula de C Guimarães, Paulo Ricardo Nascimento, Francisco Paulo Freire Neto, Sandra Rocha Gadelha, Luís Cristóvão Porto, Eloiza Helena Campana, Selma Maria Bezerra Jeronimo, Ana Tereza R Vasconcelos |
| EPI_ISL_1213275, EPI_ISL_1213277, EPI_ISL_1213279, EPI_ISL_1213282, EPI_ISL_1213284, EPI_ISL_1213288, EPI_ISL_1213289, EPI_ISL_1213291, EPI_ISL_1213293 | IMT-UFRN/RN | Bioinformatics Laboratory / LNCC | Alessandra P Lamarca, Luiz G P de Almeida, Ronaldo da Silva Francisco Jr, Lucymara Fassarella Agnez Lima, Kátia Castanho Scortecci, Vinícius Pietta Perez, Otavio J. Brustolini, Eduardo Sérgio Soares Sousa, Danielle Angst Secco, Angela Maria Guimarães Santos, George Rego Albuquerque, Ana Paula Melo Mariano, Bianca Mendes Maciel, Alexandra L Gerber, Ana Paula de C Guimarães, Paulo Ricardo Nascimento, Francisco Paulo Freire Neto, Sandra Rocha Gadelha, Luís Cristóvão Porto, Eloiza Helena Campana, Selma Maria Bezerra Jeronimo, Ana Tereza R Vasconcelos |
| EPI_ISL_1213297, EPI_ISL_1213298, EPI_ISL_1213300, EPI_ISL_1213302, EPI_ISL_1213305 | LAFEM/UESC | Bioinformatics Laboratory / LNCC | Alessandra P Lamarca, Luiz G P de Almeida, Ronaldo da Silva Francisco Jr, Lucymara Fassarella Agnez Lima, Kátia Castanho Scortecci, Vinícius Pietta Perez, Otavio J. Brustolini, Eduardo Sérgio Soares Sousa, Danielle Angst Secco, Angela Maria Guimarães Santos, George Rego Albuquerque, Ana Paula Melo Mariano, Bianca Mendes Maciel, Alexandra L Gerber, Ana Paula de C Guimarães, Paulo Ricardo Nascimento, Francisco Paulo Freire Neto, Sandra Rocha Gadelha, Luís Cristóvão Porto, Eloiza Helena Campana, Selma Maria Bezerra Jeronimo, Ana Tereza R Vasconcelos |
| EPI_ISL_1213307, EPI_ISL_1213310, EPI_ISL_1213312, EPI_ISL_1213314 | Laboratório HLA/UERJ | Bioinformatics Laboratory / LNCC | Alessandra P Lamarca, Luiz G P de Almeida, Ronaldo da Silva Francisco Jr, Lucymara Fassarella Agnez Lima, Kátia Castanho Scortecci, Vinícius Pietta Perez, Otavio J. Brustolini, Eduardo Sérgio Soares Sousa, Danielle Angst Secco, Angela Maria Guimarães Santos, George Rego Albuquerque, Ana Paula Melo Mariano, Bianca Mendes Maciel, Alexandra L Gerber, Ana Paula de C Guimarães, Paulo Ricardo Nascimento, Francisco Paulo Freire Neto, Sandra Rocha Gadelha, Luís Cristóvão Porto, Eloiza Helena Campana, Selma Maria Bezerra Jeronimo, Ana Tereza R Vasconcelos |

| | | | |
|---|---|---|---|
| EPI_ISL_1213315, EPI_ISL_1213319, EPI_ISL_1213320, EPI_ISL_1213322 | IMT-UFRN/RN | Bioinformatics Laboratory / LNCC | Alessandra P Lamarca, Luiz G P de Almeida, Ronaldo da Silva Francisco Jr, Lucymara Fassarella Agnez Lima, Kátia Castanho Scortecci, Vinícius Pietta Perez, Otavio J. Brustolini, Eduardo Sérgio Soares Sousa, Danielle Angst Secco, Angela Maria Guimarães Santos, George Rego Albuquerque, Ana Paula Melo Mariano, Bianca Mendes Maciel, Alexandra L Gerber, Ana Paula de C Guimarães, Paulo Ricardo Nascimento, Francisco Paulo Freire Neto, Sandra Rocha Gadelha, Luís Cristóvão Porto, Eloiza Helena Campana, Selma Maria Bezerra Jeronimo, Ana Tereza R Vasconcelos |
| EPI_ISL_1213324, EPI_ISL_1213326 | LAFEM/UESC | Bioinformatics Laboratory / LNCC | Alessandra P Lamarca, Luiz G P de Almeida, Ronaldo da Silva Francisco Jr, Lucymara Fassarella Agnez Lima, Kátia Castanho Scortecci, Vinícius Pietta Perez, Otavio J. Brustolini, Eduardo Sérgio Soares Sousa, Danielle Angst Secco, Angela Maria Guimarães Santos, George Rego Albuquerque, Ana Paula Melo Mariano, Bianca Mendes Maciel, Alexandra L Gerber, Ana Paula de C Guimarães, Paulo Ricardo Nascimento, Francisco Paulo Freire Neto, Sandra Rocha Gadelha, Luís Cristóvão Porto, Eloiza Helena Campana, Selma Maria Bezerra Jeronimo, Ana Tereza R Vasconcelos |
| EPI_ISL_1213329 | IMT-UFRN/RN | Bioinformatics Laboratory / LNCC | Alessandra P Lamarca, Luiz G P de Almeida, Ronaldo da Silva Francisco Jr, Lucymara Fassarella Agnez Lima, Kátia Castanho Scortecci, Vinícius Pietta Perez, Otavio J. Brustolini, Eduardo Sérgio Soares Sousa, Danielle Angst Secco, Angela Maria Guimarães Santos, George Rego Albuquerque, Ana Paula Melo Mariano, Bianca Mendes Maciel, Alexandra L Gerber, Ana Paula de C Guimarães, Paulo Ricardo Nascimento, Francisco Paulo Freire Neto, Sandra Rocha Gadelha, Luís Cristóvão Porto, Eloiza Helena Campana, Selma Maria Bezerra Jeronimo, Ana Tereza R Vasconcelos |
| EPI_ISL_1213331, EPI_ISL_1213333, EPI_ISL_1213335 | LAFEM/UESC | Bioinformatics Laboratory / LNCC | Alessandra P Lamarca, Luiz G P de Almeida, Ronaldo da Silva Francisco Jr, Lucymara Fassarella Agnez Lima, Kátia Castanho Scortecci, Vinícius Pietta Perez, Otavio J. Brustolini, Eduardo Sérgio Soares Sousa, Danielle Angst Secco, Angela Maria Guimarães Santos, George Rego Albuquerque, Ana Paula Melo Mariano, Bianca Mendes Maciel, Alexandra L Gerber, Ana Paula de C Guimarães, Paulo Ricardo Nascimento, Francisco Paulo Freire Neto, Sandra Rocha Gadelha, Luís Cristóvão Porto, Eloiza Helena Campana, Selma Maria Bezerra Jeronimo, Ana Tereza R Vasconcelos |
| EPI_ISL_1213336 | LBM/UFPB | Bioinformatics Laboratory / LNCC | Alessandra P Lamarca, Luiz G P de Almeida, Ronaldo da Silva Francisco Jr, Lucymara Fassarella Agnez Lima, Kátia Castanho Scortecci, Vinícius Pietta Perez, Otavio J. Brustolini, Eduardo Sérgio Soares Sousa, Danielle Angst Secco, Angela Maria Guimarães Santos, George Rego Albuquerque, Ana Paula Melo Mariano, Bianca Mendes Maciel, Alexandra L Gerber, Ana Paula de C Guimarães, Paulo Ricardo Nascimento, Francisco Paulo Freire Neto, Sandra Rocha Gadelha, Luís Cristóvão Porto, Eloiza Helena Campana, Selma Maria Bezerra Jeronimo, Ana Tereza R Vasconcelos |
| EPI_ISL_1213339, EPI_ISL_1213340 | Laboratório HLA/UERJ | Bioinformatics Laboratory / LNCC | Alessandra P Lamarca, Luiz G P de Almeida, Ronaldo da Silva Francisco Jr, Lucymara Fassarella Agnez Lima, Kátia Castanho Scortecci, Vinícius Pietta Perez, Otavio J. Brustolini, Eduardo Sérgio Soares Sousa, Danielle Angst Secco, Angela Maria Guimarães Santos, George Rego Albuquerque, Ana Paula Melo Mariano, Bianca Mendes Maciel, Alexandra L Gerber, Ana Paula de C Guimarães, Paulo Ricardo Nascimento, Francisco Paulo Freire Neto, Sandra Rocha Gadelha, Luís Cristóvão Porto, Eloiza Helena Campana, Selma Maria Bezerra Jeronimo, Ana Tereza R Vasconcelos |
| EPI_ISL_1213341, EPI_ISL_1213342 | LAFEM/UESC | Bioinformatics Laboratory / LNCC | Alessandra P Lamarca, Luiz G P de Almeida, Ronaldo da Silva Francisco Jr, Lucymara Fassarella Agnez Lima, Kátia Castanho Scortecci, Vinícius Pietta Perez, Otavio J. Brustolini, Eduardo Sérgio Soares Sousa, Danielle Angst Secco, Angela Maria Guimarães Santos, George Rego Albuquerque, Ana Paula Melo Mariano, Bianca Mendes Maciel, Alexandra L Gerber, Ana Paula de C Guimarães, Paulo Ricardo Nascimento, Francisco Paulo Freire Neto, Sandra Rocha Gadelha, Luís Cristóvão Porto, Eloiza Helena Campana, Selma Maria Bezerra Jeronimo, Ana Tereza R Vasconcelos |
| EPI_ISL_1213344 | LBM/UFPB | Bioinformatics Laboratory / LNCC | Alessandra P Lamarca, Luiz G P de Almeida, Ronaldo da Silva Francisco Jr, Lucymara Fassarella Agnez Lima, Kátia Castanho Scortecci, Vinícius Pietta Perez, Otavio J. Brustolini, Eduardo Sérgio Soares Sousa, Danielle Angst Secco, Angela Maria Guimarães Santos, George Rego Albuquerque, Ana Paula Melo Mariano, Bianca Mendes Maciel, Alexandra L Gerber, Ana Paula de C Guimarães, Paulo Ricardo Nascimento, Francisco Paulo Freire Neto, Sandra Rocha Gadelha, Luís Cristóvão Porto, Eloiza Helena Campana, Selma Maria Bezerra Jeronimo, Ana Tereza R Vasconcelos |
| EPI_ISL_1213345, EPI_ISL_1213346, EPI_ISL_1213348, EPI_ISL_1213350, EPI_ISL_1213352, EPI_ISL_1213353, EPI_ISL_1213355, EPI_ISL_1213357, EPI_ISL_1213358, EPI_ISL_1213360, EPI_ISL_1213362, EPI_ISL_1213364 | | | |
| see above | IMT-UFRN/RN | Bioinformatics Laboratory / LNCC | Alessandra P Lamarca, Luiz G P de Almeida, Ronaldo da Silva Francisco Jr, Lucymara Fassarella Agnez Lima, Kátia Castanho Scortecci, Vinícius Pietta Perez, Otavio J. Brustolini, Eduardo Sérgio Soares Sousa, Danielle Angst Secco, Angela Maria Guimarães Santos, George Rego Albuquerque, Ana Paula Melo Mariano, Bianca Mendes Maciel, Alexandra L Gerber, Ana Paula de C Guimarães, Paulo Ricardo Nascimento, Francisco Paulo Freire Neto, Sandra Rocha Gadelha, Luís Cristóvão Porto, Eloiza Helena Campana, Selma Maria Bezerra Jeronimo, Ana Tereza R Vasconcelos |
| EPI_ISL_1213365 | LAFEM/UESC | Bioinformatics Laboratory / LNCC | Alessandra P Lamarca, Luiz G P de Almeida, Ronaldo da Silva Francisco Jr, Lucymara Fassarella Agnez Lima, Kátia Castanho Scortecci, Vinícius Pietta Perez, Otavio J. Brustolini, Eduardo Sérgio Soares Sousa, Danielle Angst Secco, Angela Maria Guimarães Santos, George Rego Albuquerque, Ana Paula Melo Mariano, Bianca Mendes Maciel, Alexandra L Gerber, Ana Paula de C Guimarães, Paulo Ricardo Nascimento, Francisco Paulo Freire Neto, Sandra Rocha Gadelha, Luís Cristóvão Porto, Eloiza Helena Campana, Selma Maria Bezerra Jeronimo, Ana Tereza R Vasconcelos |
| EPI_ISL_1213367, EPI_ISL_1213369, EPI_ISL_1213370 | LBM/UFPB | Bioinformatics Laboratory / LNCC | Alessandra P Lamarca, Luiz G P de Almeida, Ronaldo da Silva Francisco Jr, Lucymara Fassarella Agnez Lima, Kátia Castanho Scortecci, Vinícius Pietta Perez, Otavio J. Brustolini, Eduardo Sérgio Soares Sousa, Danielle Angst Secco, Angela Maria Guimarães Santos, George Rego Albuquerque, Ana Paula Melo Mariano, Bianca Mendes Maciel, Alexandra L Gerber, Ana Paula de C Guimarães, Paulo Ricardo Nascimento, Francisco Paulo Freire Neto, Sandra Rocha Gadelha, Luís Cristóvão Porto, Eloiza Helena Campana, Selma Maria Bezerra Jeronimo, Ana Tereza R Vasconcelos |
| EPI_ISL_1213372, EPI_ISL_1213374, EPI_ISL_1213376, EPI_ISL_1213378, EPI_ISL_1213379, EPI_ISL_1213382 | Laboratório HLA/UERJ | Bioinformatics Laboratory / LNCC | Alessandra P Lamarca, Luiz G P de Almeida, Ronaldo da Silva Francisco Jr, Lucymara Fassarella Agnez Lima, Kátia Castanho Scortecci, Vinícius Pietta Perez, Otavio J. Brustolini, Eduardo Sérgio Soares Sousa, Danielle Angst Secco, Angela Maria Guimarães Santos, George Rego Albuquerque, Ana Paula Melo Mariano, Bianca Mendes Maciel, Alexandra L Gerber, Ana Paula de C Guimarães, Paulo Ricardo Nascimento, Francisco Paulo Freire Neto, Sandra Rocha Gadelha, Luís Cristóvão Porto, Eloiza Helena Campana, Selma Maria Bezerra Jeronimo, Ana Tereza R Vasconcelos |
| EPI_ISL_1213384, EPI_ISL_1213386 | LAFEM/UESC | Bioinformatics Laboratory / LNCC | Alessandra P Lamarca, Luiz G P de Almeida, Ronaldo da Silva Francisco Jr, Lucymara Fassarella Agnez Lima, Kátia Castanho Scortecci, Vinícius Pietta Perez, Otavio J. Brustolini, Eduardo Sérgio Soares Sousa, Danielle Angst Secco, Angela Maria Guimarães Santos, George Rego Albuquerque, Ana Paula Melo Mariano, Bianca Mendes Maciel, Alexandra L Gerber, Ana Paula de C Guimarães, Paulo Ricardo Nascimento, Francisco Paulo Freire Neto, Sandra Rocha Gadelha, Luís Cristóvão Porto, Eloiza Helena Campana, Selma Maria Bezerra Jeronimo, Ana Tereza R Vasconcelos |
| EPI_ISL_1213388 | LBM/UFPB | Bioinformatics Laboratory / LNCC | Alessandra P Lamarca, Luiz G P de Almeida, Ronaldo da Silva Francisco Jr, Lucymara Fassarella Agnez Lima, Kátia Castanho Scortecci, Vinícius Pietta Perez, Otavio J. Brustolini, Eduardo Sérgio Soares Sousa, Danielle Angst Secco, Angela Maria Guimarães Santos, George Rego Albuquerque, Ana Paula Melo Mariano, Bianca Mendes Maciel, Alexandra L Gerber, Ana Paula de C Guimarães, Paulo Ricardo Nascimento, Francisco Paulo Freire Neto, Sandra Rocha Gadelha, Luís Cristóvão Porto, Eloiza Helena Campana, Selma Maria Bezerra Jeronimo, Ana Tereza R Vasconcelos |
| EPI_ISL_1213390 | LAFEM/UESC | Bioinformatics Laboratory / LNCC | Alessandra P Lamarca, Luiz G P de Almeida, Ronaldo da Silva Francisco Jr, Lucymara Fassarella Agnez Lima, Kátia Castanho Scortecci, Vinícius Pietta Perez, Otavio J. Brustolini, Eduardo Sérgio Soares Sousa, Danielle Angst Secco, Angela Maria Guimarães Santos, George Rego Albuquerque, Ana Paula Melo Mariano, Bianca Mendes Maciel, Alexandra L Gerber, Ana Paula de C Guimarães, Paulo Ricardo Nascimento, Francisco Paulo Freire Neto, Sandra Rocha Gadelha, Luís Cristóvão Porto, Eloiza Helena Campana, Selma Maria Bezerra Jeronimo, Ana Tereza R Vasconcelos |
| EPI_ISL_1213392 | LBM/UFPB | Bioinformatics Laboratory / LNCC | Alessandra P Lamarca, Luiz G P de Almeida, Ronaldo da Silva Francisco Jr, Lucymara Fassarella Agnez Lima, Kátia Castanho Scortecci, Vinícius Pietta Perez, Otavio J. Brustolini, Eduardo Sérgio Soares Sousa, Danielle Angst Secco, Angela Maria Guimarães Santos, George Rego Albuquerque, Ana Paula Melo Mariano, Bianca Mendes Maciel, Alexandra L Gerber, Ana Paula de C Guimarães, Paulo Ricardo Nascimento, Francisco Paulo Freire Neto, Sandra Rocha Gadelha, Luís Cristóvão Porto, Eloiza Helena Campana, Selma Maria Bezerra Jeronimo, Ana Tereza R Vasconcelos |
| EPI_ISL_1213393, EPI_ISL_1213395 | Laboratório HLA/UERJ | Bioinformatics Laboratory / LNCC | Alessandra P Lamarca, Luiz G P de Almeida, Ronaldo da Silva Francisco Jr, Lucymara Fassarella Agnez Lima, Kátia Castanho Scortecci, Vinícius Pietta Perez, Otavio J. Brustolini, Eduardo Sérgio Soares Sousa, Danielle Angst Secco, Angela Maria Guimarães Santos, George Rego Albuquerque, Ana Paula Melo Mariano, Bianca Mendes Maciel, Alexandra L Gerber, Ana Paula de C Guimarães, Paulo Ricardo Nascimento, Francisco Paulo Freire Neto, Sandra Rocha Gadelha, Luís Cristóvão Porto, Eloiza Helena Campana, Selma Maria Bezerra Jeronimo, Ana Tereza R Vasconcelos |
| EPI_ISL_1213399, EPI_ISL_1213402 | LAFEM/UESC | Bioinformatics Laboratory / LNCC | Alessandra P Lamarca, Luiz G P de Almeida, Ronaldo da Silva Francisco Jr, Lucymara Fassarella Agnez Lima, Kátia Castanho Scortecci, Vinícius Pietta Perez, Otavio J. Brustolini, Eduardo Sérgio Soares Sousa, Danielle Angst Secco, Angela Maria Guimarães Santos, George Rego Albuquerque, Ana Paula Melo Mariano, Bianca Mendes Maciel, Alexandra L Gerber, Ana Paula de C Guimarães, Paulo Ricardo Nascimento, Francisco Paulo Freire Neto, Sandra Rocha Gadelha, Luís Cristóvão Porto, Eloiza Helena Campana, Selma Maria Bezerra Jeronimo, Ana Tereza R Vasconcelos |
| EPI_ISL_1213404, EPI_ISL_1213406 | LBM/UFPB | Bioinformatics Laboratory / LNCC | Alessandra P Lamarca, Luiz G P de Almeida, Ronaldo da Silva Francisco Jr, Lucymara Fassarella Agnez Lima, Kátia Castanho Scortecci, Vinícius Pietta Perez, Otavio J. Brustolini, Eduardo Sérgio Soares Sousa, Danielle Angst Secco, Angela Maria Guimarães Santos, George Rego Albuquerque, Ana Paula Melo Mariano, Bianca Mendes Maciel, Alexandra L Gerber, Ana Paula de C Guimarães, Paulo Ricardo Nascimento, Francisco Paulo Freire Neto, Sandra Rocha Gadelha, Luís Cristóvão Porto, Eloiza Helena Campana, Selma Maria Bezerra Jeronimo, Ana Tereza R Vasconcelos |

| | | | |
|---|---|---|---|
| EPI_ISL_1213408, EPI_ISL_1213410 | Laboratório HLA/UERJ | Bioinformatics Laboratory / LNCC | Alessandra P Lamarca, Luiz G P de Almeida, Ronaldo da Silva Francisco Jr, Lucymara Fassarella Agnez Lima, Kátia Castanho Scortecci, Vinícius Pietta Perez, Otavio J. Brustolini, Eduardo Sérgio Soares Sousa, Danielle Angst Secco, Angela Maria Guimarães Santos, George Rego Albuquerque, Ana Paula Melo Mariano, Bianca Mendes Maciel, Alexandra L Gerber, Ana Paula de C Guimarães, Paulo Ricardo Nascimento, Francisco Paulo Freire Neto, Sandra Rocha Gadelha, Luís Cristóvão Porto, Eloiza Helena Campana, Selma Maria Bezerra Jeronimo, Ana Tereza R Vasconcelos |
| EPI_ISL_1213411, EPI_ISL_1213413, EPI_ISL_1213415 | LAFEM/UESC | Bioinformatics Laboratory / LNCC | Alessandra P Lamarca, Luiz G P de Almeida, Ronaldo da Silva Francisco Jr, Lucymara Fassarella Agnez Lima, Kátia Castanho Scortecci, Vinícius Pietta Perez, Otavio J. Brustolini, Eduardo Sérgio Soares Sousa, Danielle Angst Secco, Angela Maria Guimarães Santos, George Rego Albuquerque, Ana Paula Melo Mariano, Bianca Mendes Maciel, Alexandra L Gerber, Ana Paula de C Guimarães, Paulo Ricardo Nascimento, Francisco Paulo Freire Neto, Sandra Rocha Gadelha, Luís Cristóvão Porto, Eloiza Helena Campana, Selma Maria Bezerra Jeronimo, Ana Tereza R Vasconcelos |
| EPI_ISL_1213417, EPI_ISL_1213418, EPI_ISL_1213420, EPI_ISL_1213422, EPI_ISL_1213424, EPI_ISL_1213425 | Laboratório HLA/UERJ | Bioinformatics Laboratory / LNCC | Alessandra P Lamarca, Luiz G P de Almeida, Ronaldo da Silva Francisco Jr, Lucymara Fassarella Agnez Lima, Kátia Castanho Scortecci, Vinícius Pietta Perez, Otavio J. Brustolini, Eduardo Sérgio Soares Sousa, Danielle Angst Secco, Angela Maria Guimarães Santos, George Rego Albuquerque, Ana Paula Melo Mariano, Bianca Mendes Maciel, Alexandra L Gerber, Ana Paula de C Guimarães, Paulo Ricardo Nascimento, Francisco Paulo Freire Neto, Sandra Rocha Gadelha, Luís Cristóvão Porto, Eloiza Helena Campana, Selma Maria Bezerra Jeronimo, Ana Tereza R Vasconcelos |
| EPI_ISL_1213427, EPI_ISL_1213431, EPI_ISL_1213436, EPI_ISL_1213438, EPI_ISL_1213439, EPI_ISL_1213441, EPI_ISL_1213446, EPI_ISL_1213449, EPI_ISL_1213451, EPI_ISL_1213456 | LBM/UFPB | Bioinformatics Laboratory / LNCC | Alessandra P Lamarca, Luiz G P de Almeida, Ronaldo da Silva Francisco Jr, Lucymara Fassarella Agnez Lima, Kátia Castanho Scortecci, Vinícius Pietta Perez, Otavio J. Brustolini, Eduardo Sérgio Soares Sousa, Danielle Angst Secco, Angela Maria Guimarães Santos, George Rego Albuquerque, Ana Paula Melo Mariano, Bianca Mendes Maciel, Alexandra L Gerber, Ana Paula de C Guimarães, Paulo Ricardo Nascimento, Francisco Paulo Freire Neto, Sandra Rocha Gadelha, Luís Cristóvão Porto, Eloiza Helena Campana, Selma Maria Bezerra Jeronimo, Ana Tereza R Vasconcelos |
| EPI_ISL_1219021 | Hospital Aviccena | Instituto Adolfo Lutz, Interdiciplinary Procedures Center, Strategic Laboratory | Claudio Tavares Sacchi, Claudia Regina Gonçalves, Erica Valessa Ramos Gomes, Karoline Rodrigues Campos, Caio Vinicius Dias Lopes |
| EPI_ISL_1219023, EPI_ISL_1219024, EPI_ISL_1219025, EPI_ISL_1219026 | IAL Regional de Sorocaba | Instituto Adolfo Lutz, Interdiciplinary Procedures Center, Strategic Laboratory | Claudio Tavares Sacchi, Claudia Regina Gonçalves, Erica Valessa Ramos Gomes, Karoline Rodrigues Campos, Caio Vinicius Dias Lopes |
| EPI_ISL_1219027 | IAL Regional de Presidente Prudente | Instituto Adolfo Lutz, Interdiciplinary Procedures Center, Strategic Laboratory | Claudio Tavares Sacchi, Claudia Regina Gonçalves, Erica Valessa Ramos Gomes, Karoline Rodrigues Campos, Caio Vinicius Dias Lopes |
| EPI_ISL_1219029, EPI_ISL_1219030, EPI_ISL_1219031, EPI_ISL_1219033, EPI_ISL_1219034, EPI_ISL_1219035, EPI_ISL_1219036 | Aeroporto Internacional de Guarulhos | Instituto Adolfo Lutz, Interdiciplinary Procedures Center, Strategic Laboratory | Claudio Tavares Sacchi, Claudia Regina Gonçalves, Erica Valessa Ramos Gomes, Karoline Rodrigues Campos, Caio Vinicius Dias Lopes |
| EPI_ISL_1219037 | IAL Regional de Presidente Prudente | Instituto Adolfo Lutz, Interdiciplinary Procedures Center, Strategic Laboratory | Claudio Tavares Sacchi, Claudia Regina Gonçalves, Erica Valessa Ramos Gomes, Karoline Rodrigues Campos, Caio Vinicius Dias Lopes |
| EPI_ISL_1219132, EPI_ISL_1219133 | Laboratorio Central de Saude Publica do Estado do Parana (LACEN-PR) | Laboratory of Respiratory Viruses and Measles, Oswaldo Cruz Institute, FIOCRUZ | Paola Resende, Luciana Appolinario, Fernando Motta, Anna Carolina Paixao, Ana Carolina Mendonca, Alice Sampaio Rocha, Renata Serrano Lopes, Maria do Carmo Debur, Irina Nastassja Riediger, Marilda Siqueira on behalf of the Fiocruz COVID-19 Genomic Surveillance Network |
| EPI_ISL_1219134, EPI_ISL_1219135 | Laboratorio Central de Saude Publica do Estado do Alagoas (LACEN-AL) | Laboratory of Respiratory Viruses and Measles, Oswaldo Cruz Institute, FIOCRUZ | Paola Resende, Luciana Appolinario, Fernando Motta, Anna Carolina Paixao, Ana Carolina Mendonca, Alice Sampaio Rocha, Renata Serrano Lopes, Anderson Brandao Leite, Marilda Siqueira on behalf of the Fiocruz COVID-19 Genomic Surveillance Network |
| EPI_ISL_1219136 | Gonçalo Moniz Institute, FIOCRUZ, Bahia | Laboratory of Respiratory Viruses and Measles, Oswaldo Cruz Institute, FIOCRUZ | Paola Resende, Luciana Appolinario, Fernando Motta, Anna Carolina Paixao, Ana Carolina Mendonca, Alice Sampaio Rocha, Renata Serrano Lopes, Tiago Graf, Ricardo Khouri, Marilda Siqueira on behalf of the Fiocruz COVID-19 Genomic Surveillance Network |
| EPI_ISL_1219137 | Laboratorio Central de Saude Publica do Estado de Minas Gerais (LACEN-MG) | Laboratory of Respiratory Viruses and Measles, Oswaldo Cruz Institute, FIOCRUZ | Paola Resende, Luciana Appolinario, Fernando Motta, Anna Carolina Paixao, Ana Carolina Mendonca, Alice Sampaio Rocha, Renata Serrano Lopes, Felipe Iani, Marilda Siqueira on behalf of the Fiocruz COVID-19 Genomic Surveillance Network |
| EPI_ISL_1220045 | Universidad de Magdalena | Instituto Nacional de Salud- Dirección de Investigación en Salud Pública | Katherine Laiton-Donato, Carlos Franco-Muñoz, Diego A. Álvarez-Díaz, Hector Alejandro Ruiz-Moreno, Jhonnatan Reales-González, Diego Andrés Prada, Sheryll Corchuelo, Maria T. Herrera-Sepúlveda, Julian Naizaque, Gerardo Santamaría, Magdalena Wiesner, Martha Lucia Ospina Martínez, Marcela Mercado-Reyes. |
| EPI_ISL_1220050 | Laboratorio Clinico Clinica General del Norte | Instituto Nacional de Salud- Dirección de Investigación en Salud Pública | Katherine Laiton-Donato, Carlos Franco-Muñoz, Diego A. Álvarez-Díaz, Hector Alejandro Ruiz-Moreno, Jhonnatan Reales-González, Diego Andrés Prada, Sheryll Corchuelo, Maria T. Herrera-Sepúlveda, Julian Naizaque, Gerardo Santamaría, Magdalena Wiesner, Martha Lucia Ospina Martínez, Marcela Mercado-Reyes. |
| EPI_ISL_1220056 | LABORATORIO BIOLOGIA MOLECULAR IMAT SAS | Instituto Nacional de Salud- Dirección de Investigación en Salud Pública | Katherine Laiton-Donato, Carlos Franco-Muñoz, Diego A. Álvarez-Díaz, Hector Alejandro Ruiz-Moreno, Jhonnatan Reales-González, Diego Andrés Prada, Sheryll Corchuelo, Maria T. Herrera-Sepúlveda, Julian Naizaque, Gerardo Santamaría, Magdalena Wiesner, Martha Lucia Ospina Martínez, Marcela Mercado-Reyes. |
| EPI_ISL_1220061, EPI_ISL_1220062, EPI_ISL_1220065 | Investigaciones Biomédicas - Universidad de Sucre | Instituto Nacional de Salud- Dirección de Investigación en Salud Pública | Katherine Laiton-Donato, Carlos Franco-Muñoz, Diego A. Álvarez-Díaz, Hector Alejandro Ruiz-Moreno, Jhonnatan Reales-González, Diego Andrés Prada, Sheryll Corchuelo, Maria T. Herrera-Sepúlveda, Julian Naizaque, Gerardo Santamaría, Magdalena Wiesner, Martha Lucia Ospina Martínez, Marcela Mercado-Reyes. |
| EPI_ISL_1220095 | Hospital Municipal Gov. Mario Covas Jr. | Instituto Butantan (genome assembly and bioinformatics) and Mendelics (sequencing) | Maria Carolina Quartim Barbosa Elias Sabbaga, Jose Patane, Simone Haddad, Rafael dos Santos Bezerra, Sandra Coccuzzo Sampaio Vessoni, Antonio Jorge Martins, Dimas Tadeu Covas |
| EPI_ISL_1232714, EPI_ISL_1232738 | Dutch COVID-19 response team | National Institute for Public Health and the Environment (RIVM) | Adam Meijer, Harry Vennema, Dirk Eggink, Jeroen Cremer, Sharon van den Brink, Bas van der Veer, AnneMarie van den Brandt, Florian Zwagemaker, Dennis Schmitz, Chantal Reusken, on behalf of the national COVID-19 response team |
| EPI_ISL_1239012, EPI_ISL_1239013, EPI_ISL_1239014, EPI_ISL_1239015, EPI_ISL_1239016 | Laboratório Central de Saúde Pública do Estado de Pernambuco (LACEN-PE) | WallauLab, Aggeu Magalhaes Institute | Marcelo Henrique dos Santos Paiva, Duschinka Ribeiro Duarte Guedes, Cássia Docena, Matheus Filgueira Bezerra, Filipe Zimmer Dezordi, Laís Ceschini Machado, Larissa Krokovsky, Elisama Helvecio, Alexandre Freitas da Silva, Antonio Mauro Rezende, Sinval Pinto Brandão Filho, Constância Flávia Junqueira Ayres, Gabriel Luz Wallau on behalf of the Fiocruz COVID-19 Genomic Surveillance Network |
| EPI_ISL_1239111, EPI_ISL_1239112, EPI_ISL_1239113 | Laboratório Central de Saúde Pública Noel Nutels | Coordenação Geral de Laboratórios de Saúde Pública (CGLAB) | Vagner Fonseca et al, |
| EPI_ISL_1239114, EPI_ISL_1239115, EPI_ISL_1239119, EPI_ISL_1239120, EPI_ISL_1239122, EPI_ISL_1239123 | Laboratório Central de Saúde Pública do Espírito Santo | Coordenação Geral de Laboratórios de Saúde Pública (CGLAB) | Vagner Fonseca et al, |
| EPI_ISL_1239124 | Fundação Ezequiel Dias | Coordenação Geral de Laboratórios de Saúde Pública (CGLAB) | Vagner Fonseca et al, |
| EPI_ISL_1239125, EPI_ISL_1239126, EPI_ISL_1239128, EPI_ISL_1239129, EPI_ISL_1239130, EPI_ISL_1239131, EPI_ISL_1239132, EPI_ISL_1239133, EPI_ISL_1239135, EPI_ISL_1239136 | Laboratório Central de Saúde Pública do Espírito Santo | Coordenação Geral de Laboratórios de Saúde Pública (CGLAB) | Vagner Fonseca et al, |
| EPI_ISL_1239137, EPI_ISL_1239138, EPI_ISL_1239139 | Fundação Ezequiel Dias | Coordenação Geral de Laboratórios de Saúde Pública (CGLAB) | Vagner Fonseca et al, |
| EPI_ISL_1240639, EPI_ISL_1240640, EPI_ISL_1240641 | Laboratório Central de Saúde Pública do Espírito Santo | Coordenação Geral de Laboratórios de Saúde Pública (CGLAB) | Vagner Fonseca et al. |
| EPI_ISL_1240642 | Fundação Ezequiel Dias | Coordenação Geral de Laboratórios de Saúde Pública (CGLAB) | Vagner Fonseca et al. |

| | | | |
|---|---|---|---|
| EPI_ISL_1259308 | Outre Mer | National Reference Center for Viruses of Respiratory Infections, Institut Pasteur, Paris | Marion Barbet, Sylvie Behillil, Méline Bizard, Angela Brisebarre, Camille Capel, Etienne Simon-Lorière, Vincent Enouf, Maud Vanpeene, Sylvie van der Werf,Rousset Dominique |
| EPI_ISL_1261122, EPI_ISL_1261123 | Laboratorio de Ecologia de Doencas Transmissiveis na Amazonia, Instituto Leonidas e Maria Deane - Fiocruz Amazonia | Laboratorio de Ecologia de Doencas Transmissiveis na Amazonia, Instituto Leonidas e Maria Deane - Fiocruz Amazonia | Valdinete Nascimento, Victor Souza, André Corado, Fernanda Nascimento, George Silva, Ágatha Costa, Debora Duarte, Karina Pessoa, Matilde Mejía, Luciana Gonçalves, Maria Júlia Brandão, Michele Jesus, Felipe Naveca |
| EPI_ISL_1261683 | LACEN - Laboratório Central de Saúde Pública do Amazonas | Evandro Chagas Institute | Santos, M.C.; Silva, A.M.; Junior, W.D.C.; Barbagelata, L.S.; Ferreira, J.A.; Sousa, E.M.A.; da Silva, P.S.; Pinheiro, K.C.; L.C.; Sousa Junior, E.C. |
| EPI_ISL_1261684 | LACEN - Laboratório Central de Saúde Pública do Ceará | Evandro Chagas Institute | Santos, M.C.; Silva, A.M.; Junior, W.D.C.; Barbagelata, L.S.; Ferreira, J.A.; Sousa, E.M.A.; da Silva, P.S.; Pinheiro, K.C.; L.C.; Sousa Junior, E.C. |
| EPI_ISL_1261685 | LACEN - Laboratório Central de Saúde Pública do Amazonas | Evandro Chagas Institute | Santos, M.C.; Silva, A.M.; Junior, W.D.C.; Barbagelata, L.S.; Ferreira, J.A.; Sousa, E.M.A.; da Silva, P.S.; Pinheiro, K.C.; L.C.; Sousa Junior, E.C. |
| EPI_ISL_1261686 | LACEN - Laboratório Central de Saúde Pública do Amapá | Evandro Chagas Institute | Santos, M.C.; Silva, A.M.; Junior, W.D.C.; Barbagelata, L.S.; Ferreira, J.A.; Sousa, E.M.A.; da Silva, P.S.; Pinheiro, K.C.; L.C.; Sousa Junior, E.C. |
| EPI_ISL_1261687 | LACEN - Laboratório Central de Saúde Pública de Roraima | Evandro Chagas Institute | Santos, M.C.; Silva, A.M.; Junior, W.D.C.; Barbagelata, L.S.; Ferreira, J.A.; Sousa, E.M.A.; da Silva, P.S.; Pinheiro, K.C.; L.C.; Sousa Junior, E.C. |
| EPI_ISL_1261688, EPI_ISL_1261689 | LACEN - Laboratório Central de Saúde Pública do Amapá | Evandro Chagas Institute | Santos, M.C.; Silva, A.M.; Junior, W.D.C.; Barbagelata, L.S.; Ferreira, J.A.; Sousa, E.M.A.; da Silva, P.S.; Pinheiro, K.C.; L.C.; Sousa Junior, E.C. |
| EPI_ISL_1261690 | LACEN - Laboratório Central de Saúde Pública do Amazonas | Evandro Chagas Institute | Santos, M.C.; Silva, A.M.; Junior, W.D.C.; Barbagelata, L.S.; Ferreira, J.A.; Sousa, E.M.A.; da Silva, P.S.; Pinheiro, K.C.; L.C.; Sousa Junior, E.C. |
| EPI_ISL_1261691 | Laboratório Paulo C. Azevedo | Evandro Chagas Institute | Santos, M.C.; Silva, A.M.; Junior, W.D.C.; Barbagelata, L.S.; Ferreira, J.A.; Sousa, E.M.A.; da Silva, P.S.; Pinheiro, K.C.; L.C.; Sousa Junior, E.C. |
| EPI_ISL_1261692 | LACEN - Laboratório Central de Saúde Pública do Amapá | Evandro Chagas Institute | Santos, M.C.; Silva, A.M.; Junior, W.D.C.; Barbagelata, L.S.; Ferreira, J.A.; Sousa, E.M.A.; da Silva, P.S.; Pinheiro, K.C.; L.C.; Sousa Junior, E.C. |
| EPI_ISL_1261693 | LACEN - Laboratório Central de Saúde Pública do Ceará | Evandro Chagas Institute | Santos, M.C.; Silva, A.M.; Junior, W.D.C.; Barbagelata, L.S.; Ferreira, J.A.; Sousa, E.M.A.; da Silva, P.S.; Pinheiro, K.C.; L.C.; Sousa Junior, E.C. |
| EPI_ISL_1261694 | LACEN - Laboratório Central de Saúde Pública do Amazonas | Evandro Chagas Institute | Santos, M.C.; Silva, A.M.; Junior, W.D.C.; Barbagelata, L.S.; Ferreira, J.A.; Sousa, E.M.A.; da Silva, P.S.; Pinheiro, K.C.; L.C.; Sousa Junior, E.C. |
| EPI_ISL_1261695, EPI_ISL_1261696 | LACEN - Laboratório Central de Saúde Pública do Amapá | Evandro Chagas Institute | Santos, M.C.; Silva, A.M.; Junior, W.D.C.; Barbagelata, L.S.; Ferreira, J.A.; Sousa, E.M.A.; da Silva, P.S.; Pinheiro, K.C.; L.C.; Sousa Junior, E.C. |
| EPI_ISL_1261697 | LACEN - Laboratório Central de Saúde Pública do Ceará | Evandro Chagas Institute | Santos, M.C.; Silva, A.M.; Junior, W.D.C.; Barbagelata, L.S.; Ferreira, J.A.; Sousa, E.M.A.; da Silva, P.S.; Pinheiro, K.C.; L.C.; Sousa Junior, E.C. |
| EPI_ISL_1261699 | LACEN - Laboratório Central de Saúde Pública de Pernambuco | Evandro Chagas Institute | Santos, M.C.; Silva, A.M.; Junior, W.D.C.; Barbagelata, L.S.; Ferreira, J.A.; Sousa, E.M.A.; da Silva, P.S.; Pinheiro, K.C.; L.C.; Sousa Junior, E.C. |
| EPI_ISL_1261700 | LACEN - Laboratório Central de Saúde Pública do Maranhao | Evandro Chagas Institute | Santos, M.C.; Silva, A.M.; Junior, W.D.C.; Barbagelata, L.S.; Ferreira, J.A.; Sousa, E.M.A.; da Silva, P.S.; Pinheiro, K.C.; L.C.; Sousa Junior, E.C. |
| EPI_ISL_1262909, EPI_ISL_1262910, EPI_ISL_1262911, EPI_ISL_1262912, EPI_ISL_1262913 | Outre Mer | National Reference Center for Viruses of Respiratory Infections, Institut Pasteur, Paris | Marion Barbet, Sylvie Behillil, Méline Bizard, Angela Brisebarre, Camille Capel, Etienne Simon-Lorière, Vincent Enouf, Maud Vanpeene, Sylvie van der Werf,Rousset Dominique |
| EPI_ISL_1272236 | Universidade Federal do Norte do Tocantins (UFNT) | Laboratório de Bioinformática e Biotecnologia (Labinftec/UFT) | Ueric José Borges de Souza, Fabrício Souza Campos, Raíssa Nunes dos Santos, José Carlos Ribeiro Júnior, Rogério Fernandes Carvalho, Monike da Silva Oliveira, Bergmann Morais Ribeiro, Fernando Lucas Melo |
| EPI_ISL_1278274 | Laboratory of Virology, Ribeirão Preto General Hospital, Ribeirão Preto Medical School, University of São Paulo | Laboratory of Oncology, Blood Center of Ribeirão Preto, Ribeirão Preto School of Medicine, University of São Paulo | CAMPOS, MR; SANTOS, A.L.P.; YAMAMOTO, A.Y.; COLLI, L.M.; FONSECA, B.A.L.; BELLISSIMO-RODRIGUES, F. |
| EPI_ISL_1289226, EPI_ISL_1289228, EPI_ISL_1289229 | Dutch COVID-19 response team | National Institute for Public Health and the Environment (RIVM) | Adam Meijer, Harry Vennema, Dirk Eggink, Jeroen Cremer, Sharon van den Brink, Bas van der Veer, AnneMarie van den Brandt, Florian Zwagemaker, Dennis Schmitz, Chantal Reusken, on behalf of the national COVID-19 response team |
| EPI_ISL_1289956, EPI_ISL_1289957, EPI_ISL_1289958, EPI_ISL_1289959, EPI_ISL_1289960 | Laboratory of Virology, Ribeirão Preto General Hospital, Ribeirão Preto Medical School, University of São Paulo | Laboratory of Oncology, Blood Center of Ribeirão Preto, Ribeirão Preto School of Medicine, University of São Paulo | CAMPOS, MR; SANTOS, A.L.P.; YAMAMOTO, A.Y.; COLLI, L.M.; FONSECA, B.A.L.; BELLISSIMO-RODRIGUES, F. |
| EPI_ISL_1290802 | Genomic and molecular Biology Group, A.C.Camargo Cancer Center | Laboratory of Bioinformatics and Computational Biology, A.C.Camargo Cancer Center | Giovana Torrezan, Dirce Carraro, Israel Tojal, Alexandre Defelicibus |
| EPI_ISL_1293051 | LACEN do Distrito Federal | Instituto Adolfo Lutz, Interdiciplinary Procedures Center, Strategic Laboratory | Claudio Tavares Sacchi, Claudia Regina Gonçalves, Erica Valessa Ramos Gomes, Karoline Rodrigues Campos, Caio Vinicius Dias Lopes |
| EPI_ISL_1293053, EPI_ISL_1293054, EPI_ISL_1293055 | LACEN de Rondonia | Instituto Adolfo Lutz, Interdiciplinary Procedures Center, Strategic Laboratory | Claudio Tavares Sacchi, Claudia Regina Gonçalves, Erica Valessa Ramos Gomes, Karoline Rodrigues Campos, Caio Vinicius Dias Lopes |
| EPI_ISL_1293058, EPI_ISL_1293059, EPI_ISL_1293060, EPI_ISL_1293061, EPI_ISL_1293063, EPI_ISL_1293065, EPI_ISL_1293066, EPI_ISL_1293067, EPI_ISL_1293068, EPI_ISL_1293071, EPI_ISL_1293073, EPI_ISL_1293074, EPI_ISL_1293075, EPI_ISL_1293076, EPI_ISL_1293077, EPI_ISL_1293078, EPI_ISL_1293079, EPI_ISL_1293080 | | | |
| see above | IAL Regional de Sorocaba | Instituto Adolfo Lutz, Interdiciplinary Procedures Center, Strategic Laboratory | Claudio Tavares Sacchi, Claudia Regina Gonçalves, Erica Valessa Ramos Gomes, Karoline Rodrigues Campos, Caio Vinicius Dias Lopes |
| EPI_ISL_1300494 | Genetica Molecular and Subdepartamento de Virologia ISP Chile | Instituto de Salud Publica de Chile | Javier Tognarelli, Karen Orostica, Barbara Parra, Loredana Arata, Jaime Lagos, Gisselle Barra, Patricia Bustos, Rodrigo Fasce, Andres Castillo, Jorge Fernandez |
| EPI_ISL_1303366, EPI_ISL_1303367 | Instituto Nacional de Salud- Dirección de Redes de Laboratorios de Salud Pública | Instituto Nacional de Salud- Dirección de Investigación en Salud Pública | Katherine Laiton-Donato, Diego A. Álvarez-Díaz, Carlos Franco-Muñoz, Hector Alejandro Ruiz-Moreno, Maria T. Herrera-Sepúlveda, Diego Andrés Prada, Jhonnatan Reales-González, Sheryll Corchuelo, Julian Naizaque, Gerardo Santamaría, Sergio Gomez, Lisseth Pardo, Juan Camilo Martinez, Marta Lopez Blanco, Ángela Alarcon Cruz, Diana Malo, Carmen Osorio, Magdalena Wiesner, Martha Lucia Ospina Martinez, Marcela Mercado-Reyes |
| EPI_ISL_1303368 | Clinica de la Costa | Instituto Nacional de Salud- Dirección de Investigación en Salud Pública | Katherine Laiton-Donato, Diego A. Álvarez-Díaz, Carlos Franco-Muñoz, Hector Alejandro Ruiz-Moreno, Maria T. Herrera-Sepúlveda, Diego Andrés Prada, Jhonnatan Reales-González, Sheryll Corchuelo, Julian Naizaque, Gerardo Santamaría, Sergio Gomez, Lisseth Pardo, Juan Camilo Martinez, Marta Lopez Blanco, Ángela Alarcon Cruz, Diana Malo, Carmen Osorio, Magdalena Wiesner, Martha Lucia Ospina Martinez, Marcela Mercado-Reyes |
| EPI_ISL_1303371, EPI_ISL_1303372, EPI_ISL_1303373 | Dirección de Sanidad Policia Nacional | Instituto Nacional de Salud- Dirección de Investigación en Salud Pública | Katherine Laiton-Donato, Diego A. Álvarez-Díaz, Carlos Franco-Muñoz, Hector Alejandro Ruiz-Moreno, Maria T. Herrera-Sepúlveda, Diego Andrés Prada, Jhonnatan Reales-González, Sheryll Corchuelo, Julian Naizaque, Gerardo Santamaría, Sergio Gomez, Lisseth Pardo, Juan Camilo Martinez, Marta Lopez Blanco, Ángela Alarcon Cruz, Diana Malo, Carmen Osorio, Magdalena Wiesner, Martha Lucia Ospina Martinez, Marcela Mercado-Reyes |
| EPI_ISL_1303374 | CLINICA COLSANITAS CENTRAL DE REFERENCIA | Instituto Nacional de Salud- Dirección de Investigación en Salud Pública | Katherine Laiton-Donato, Diego A. Álvarez-Díaz, Carlos Franco-Muñoz, Hector Alejandro Ruiz-Moreno, Maria T. Herrera-Sepúlveda, Diego Andrés Prada, Jhonnatan Reales-González, Sheryll Corchuelo, Julian Naizaque, Gerardo Santamaría, Sergio Gomez, Lisseth Pardo, Juan Camilo Martinez, Marta Lopez Blanco, Ángela Alarcon Cruz, Diana Malo, Carmen Osorio, Magdalena Wiesner, Martha Lucia Ospina Martinez, Marcela Mercado-Reyes |
| EPI_ISL_1303377 | Instituto Nacional de Salud- Dirección de Redes de Laboratorios de Salud Pública | Instituto Nacional de Salud- Dirección de Investigación en Salud Pública | Katherine Laiton-Donato, Diego A. Álvarez-Díaz, Carlos Franco-Muñoz, Hector Alejandro Ruiz-Moreno, Maria T. Herrera-Sepúlveda, Diego Andrés Prada, Jhonnatan Reales-González, Sheryll Corchuelo, Julian Naizaque, Gerardo Santamaría, Sergio Gomez, Lisseth Pardo, Juan Camilo Martinez, Marta Lopez Blanco, Ángela Alarcon Cruz, Diana Malo, Carmen Osorio, Magdalena Wiesner, Martha Lucia Ospina Martinez, Marcela Mercado-Reyes |
| EPI_ISL_1303502, EPI_ISL_1303503, EPI_ISL_1303505 | LACEN de Rondonia | Instituto Adolfo Lutz, Interdiciplinary Procedures Center, Strategic Laboratory | Claudio Tavares Sacchi, Claudia Regina Gonçalves, Erica Valessa Ramos Gomes, Karoline Rodrigues Campos, Caio Vinicius Dias Lopes |
| EPI_ISL_1303506, EPI_ISL_1303507 | LACEN do Distrito Federal | Instituto Adolfo Lutz, Interdiciplinary Procedures Center, Strategic Laboratory | Claudio Tavares Sacchi, Claudia Regina Gonçalves, Erica Valessa Ramos Gomes, Karoline Rodrigues Campos, Caio Vinicius Dias Lopes |
| EPI_ISL_1303509 | LACEN do Estado de Tocantins | Instituto Adolfo Lutz, Interdiciplinary Procedures Center, Strategic Laboratory | Claudio Tavares Sacchi, Claudia Regina Gonçalves, Erica Valessa Ramos Gomes, Karoline Rodrigues Campos, Caio Vinicius Dias Lopes |
| EPI_ISL_1303510, EPI_ISL_1303511, EPI_ISL_1303512, EPI_ISL_1303513, EPI_ISL_1303514, EPI_ISL_1303515, EPI_ISL_1303516, EPI_ISL_1303517 | LACEN do Estado de Goias | Instituto Adolfo Lutz, Interdiciplinary Procedures Center, Strategic Laboratory | Claudio Tavares Sacchi, Claudia Regina Gonçalves, Erica Valessa Ramos Gomes, Karoline Rodrigues Campos, Caio Vinicius Dias Lopes |

| | | | |
|---|---|---|---|
| EPI_ISL_1303518, EPI_ISL_1303519, EPI_ISL_1303520, EPI_ISL_1303522, EPI_ISL_1303523, EPI_ISL_1303524, EPI_ISL_1303525, EPI_ISL_1303526, EPI_ISL_1303529, EPI_ISL_1303530, EPI_ISL_1303531, EPI_ISL_1303532, EPI_ISL_1303533, EPI_ISL_1303534 | | | |
| see above | IAL Regional de São Jose do Rio Preto | Instituto Adolfo Lutz, Interdiciplinary Procedures Center, Strategic Laboratory | Claudio Tavares Sacchi, Claudia Regina Gonçalves, Erica Valessa Ramos Gomes, Karoline Rodrigues Campos, Caio Vinicius Dias Lopes |
| EPI_ISL_1303535, EPI_ISL_1303536 | Hospital Heliopolis | Instituto Adolfo Lutz, Interdiciplinary Procedures Center, Strategic Laboratory | Claudio Tavares Sacchi, Claudia Regina Gonçalves, Erica Valessa Ramos Gomes, Karoline Rodrigues Campos, Caio Vinicius Dias Lopes |
| EPI_ISL_1303537 | Hospital Estadual de Vila Alpina | Instituto Adolfo Lutz, Interdiciplinary Procedures Center, Strategic Laboratory | Claudio Tavares Sacchi, Claudia Regina Gonçalves, Erica Valessa Ramos Gomes, Karoline Rodrigues Campos, Caio Vinicius Dias Lopes |
| EPI_ISL_1303538, EPI_ISL_1303539 | Hospital Presidente | Instituto Adolfo Lutz, Interdiciplinary Procedures Center, Strategic Laboratory | Claudio Tavares Sacchi, Claudia Regina Gonçalves, Erica Valessa Ramos Gomes, Karoline Rodrigues Campos, Caio Vinicius Dias Lopes |
| EPI_ISL_1303540, EPI_ISL_1303541 | Hospital Municipal Cidade Tiradentes Carmen Prudente | Instituto Adolfo Lutz, Interdiciplinary Procedures Center, Strategic Laboratory | Claudio Tavares Sacchi, Claudia Regina Gonçalves, Erica Valessa Ramos Gomes, Karoline Rodrigues Campos, Caio Vinicius Dias Lopes |
| EPI_ISL_1303542, EPI_ISL_1303543 | Hospital Estadual de Campanha Barradas | Instituto Adolfo Lutz, Interdiciplinary Procedures Center, Strategic Laboratory | Claudio Tavares Sacchi, Claudia Regina Gonçalves, Erica Valessa Ramos Gomes, Karoline Rodrigues Campos, Caio Vinicius Dias Lopes |
| EPI_ISL_1303545 | Hospital Municipal Cidade Tiradentes Carmen Prudente | Instituto Adolfo Lutz, Interdiciplinary Procedures Center, Strategic Laboratory | Claudio Tavares Sacchi, Claudia Regina Gonçalves, Erica Valessa Ramos Gomes, Karoline Rodrigues Campos, Caio Vinicius Dias Lopes |
| EPI_ISL_1303546, EPI_ISL_1303547, EPI_ISL_1303548, EPI_ISL_1303549 | UPA Vila Santa Catarina | Instituto Adolfo Lutz, Interdiciplinary Procedures Center, Strategic Laboratory | Claudio Tavares Sacchi, Claudia Regina Gonçalves, Erica Valessa Ramos Gomes, Karoline Rodrigues Campos, Caio Vinicius Dias Lopes |
| EPI_ISL_1321468, EPI_ISL_1321469, EPI_ISL_1321470, EPI_ISL_1321471, EPI_ISL_1321472, EPI_ISL_1321473, EPI_ISL_1321493, EPI_ISL_1321506, EPI_ISL_1321524, EPI_ISL_1321528, EPI_ISL_1321557, EPI_ISL_1321574, EPI_ISL_1321575 | | | |
| see above | Genetica Molecular and Subdepartamento de Virologia ISP Chile | Instituto de Salud Publica de Chile | Javier Tognarelli, Karen Orostica, Barbara Parra, Loredana Arata, Jaime Lagos, Gisselle Barra, Patricia Bustos, Rodrigo Fasce, Andres Castillo, Jorge Fernandez |
| EPI_ISL_1324142, EPI_ISL_1324145 | UW Virology Lab | UW Virology Lab | Pavitra Roychoudhury, Hong Xie, Lasata Shrestha, Shah Mohamed Bakhash, Michelle Lin, Margaret Mills, Noah Baker, Sean Ellis, Saraswathi Sathees, Meei-Li Huang, Keith R Jerome, Alexander Greninger |
| EPI_ISL_1336180 | Virology Center Adolfo Lutz Institute Sao Paulo Brazil | Retrovirus Laboratory Adolfo Lutz Institute | Gabriela Bastos Cabral, Giselle I S Lopez-Lopes, Cintia Ahagon, Audrey Cilli, Paula Morena Guimaraes, Igor Mohamed Hussein,Junior Pereira de Souza , Roberta Shiavon, Luis Brigido |
| EPI_ISL_1336181 | Virology Center Adolfo Lutz Institute Sao Paulo Brazil | Retrovirus Laboratory Adolfo Lutz Institute | Gabriela Bastos Cabral, Giselle I S Lopez-Lopes, Cintia Ahagon, Audrey Cilli, Paula Morena Guimaraes, Igor Mohamed Hussein, Junior Pereira de Souza , Roberta Shiavon, Luis Brigido |
| EPI_ISL_1358285 | Centro de Treinamento e Referencia DST AIDS | Instituto Adolfo Lutz, Interdiciplinary Procedures Center, Strategic Laboratory | Claudio Tavares Sacchi, Claudia Regina Gonçalves, Erica Valessa Ramos Gomes, Karoline Rodrigues Campos, Caio Vinicius Dias Lopes |
| EPI_ISL_1358286 | Hospital Municipal Josanias Castanha Braga | Instituto Adolfo Lutz, Interdiciplinary Procedures Center, Strategic Laboratory | Claudio Tavares Sacchi, Claudia Regina Gonçalves, Erica Valessa Ramos Gomes, Karoline Rodrigues Campos, Caio Vinicius Dias Lopes |
| EPI_ISL_1358287 | Hospital Nipo Brasileiro | Instituto Adolfo Lutz, Interdiciplinary Procedures Center, Strategic Laboratory | Claudio Tavares Sacchi, Claudia Regina Gonçalves, Erica Valessa Ramos Gomes, Karoline Rodrigues Campos, Caio Vinicius Dias Lopes |
| EPI_ISL_1358288, EPI_ISL_1358289, EPI_ISL_1358290 | IAL Regional de Aracatuba | Instituto Adolfo Lutz, Interdiciplinary Procedures Center, Strategic Laboratory | Claudio Tavares Sacchi, Claudia Regina Gonçalves, Erica Valessa Ramos Gomes, Karoline Rodrigues Campos, Caio Vinicius Dias Lopes |
| EPI_ISL_1358291, EPI_ISL_1358292, EPI_ISL_1358293, EPI_ISL_1358294, EPI_ISL_1358295, EPI_ISL_1358298, EPI_ISL_1358299 | IAL Regional de Santo Andre | Instituto Adolfo Lutz, Interdiciplinary Procedures Center, Strategic Laboratory | Claudio Tavares Sacchi, Claudia Regina Gonçalves, Erica Valessa Ramos Gomes, Karoline Rodrigues Campos, Caio Vinicius Dias Lopes |
| EPI_ISL_1358300, EPI_ISL_1358301, EPI_ISL_1358303 | Lacen de Tocantins | Instituto Adolfo Lutz, Interdiciplinary Procedures Center, Strategic Laboratory | Claudio Tavares Sacchi, Claudia Regina Gonçalves, Erica Valessa Ramos Gomes, Karoline Rodrigues Campos, Caio Vinicius Dias Lopes |
| EPI_ISL_1358304, EPI_ISL_1358306, EPI_ISL_1358310, EPI_ISL_1358312, EPI_ISL_1358313, EPI_ISL_1358314, EPI_ISL_1358315, EPI_ISL_1358316, EPI_ISL_1358317 | LACEN do Mato Grosso do Sul | Instituto Adolfo Lutz, Interdiciplinary Procedures Center, Strategic Laboratory | Claudio Tavares Sacchi, Claudia Regina Gonçalves, Erica Valessa Ramos Gomes, Karoline Rodrigues Campos, Caio Vinicius Dias Lopes |
| EPI_ISL_1358318, EPI_ISL_1358319, EPI_ISL_1358320, EPI_ISL_1358321 | UPA Vila Santa Catarina | Instituto Adolfo Lutz, Interdiciplinary Procedures Center, Strategic Laboratory | Claudio Tavares Sacchi, Claudia Regina Gonçalves, Erica Valessa Ramos Gomes, Karoline Rodrigues Campos, Caio Vinicius Dias Lopes |
| EPI_ISL_1365747 | Associação Fundo de Incentivo a Pesquisa | Associação Fundo de Incentivo a Pesquisa | Priscila Farias Tempaku, Juliana Nogueira Martins Rodrigues, Erika Rodrigues de Oliveira, Soraya Sgambatti de Andrade, Debora Ribeiro Ramadan, Sergio Tufik |
| EPI_ISL_1370553, EPI_ISL_1370556, EPI_ISL_1370557, EPI_ISL_1370577, EPI_ISL_1370667, EPI_ISL_1371227, EPI_ISL_1371228, EPI_ISL_1371236, EPI_ISL_1371265 | Dutch COVID-19 response team | National Institute for Public Health and the Environment (RIVM) | Adam Meijer, Harry Vennema, Dirk Eggink, Jeroen Cremer, Sharon van den Brink, Bas van der Veer, AnneMarie van den Brandt, Florian Zwagemaker, Dennis Schmitz, Chantal Reusken, on behalf of the national COVID-19 response team |
| EPI_ISL_1381043, EPI_ISL_1381045, EPI_ISL_1381047, EPI_ISL_1381048, EPI_ISL_1381050, EPI_ISL_1381051, EPI_ISL_1381052, EPI_ISL_1381053, EPI_ISL_1381054, EPI_ISL_1381055, EPI_ISL_1381056, EPI_ISL_1381057, EPI_ISL_1381058, EPI_ISL_1381059, EPI_ISL_1381060, EPI_ISL_1381061, EPI_ISL_1381062, EPI_ISL_1381063, EPI_ISL_1381065 | | | |
| see above | IAL Regional de Santo Andre | Instituto Adolfo Lutz, Interdiciplinary Procedures Center, Strategic Laboratory | Claudio Tavares Sacchi, Claudia Regina Gonçalves, Erica Valessa Ramos Gomes, Karoline Rodrigues Campos, Caio Vinicius Dias Lopes |
| EPI_ISL_1381066 | LACEN do Mato Grosso do Sul | Instituto Adolfo Lutz, Interdiciplinary Procedures Center, Strategic Laboratory | Claudio Tavares Sacchi, Claudia Regina Gonçalves, Erica Valessa Ramos Gomes, Karoline Rodrigues Campos, Caio Vinicius Dias Lopes |
| EPI_ISL_1381068 | Conjunto Hospitalar do Mandaqui de Sao Paulo | Instituto Adolfo Lutz, Interdiciplinary Procedures Center, Strategic Laboratory | Claudio Tavares Sacchi, Claudia Regina Gonçalves, Erica Valessa Ramos Gomes, Karoline Rodrigues Campos, Caio Vinicius Dias Lopes |
| EPI_ISL_1381069 | Hospital Heliopolis | Instituto Adolfo Lutz, Interdiciplinary Procedures Center, Strategic Laboratory | Claudio Tavares Sacchi, Claudia Regina Gonçalves, Erica Valessa Ramos Gomes, Karoline Rodrigues Campos, Caio Vinicius Dias Lopes |
| EPI_ISL_1381070, EPI_ISL_1381071 | Hospital Municipal Cidade Tiradentes Carmem Prudente | Instituto Adolfo Lutz, Interdiciplinary Procedures Center, Strategic Laboratory | Claudio Tavares Sacchi, Claudia Regina Gonçalves, Erica Valessa Ramos Gomes, Karoline Rodrigues Campos, Caio Vinicius Dias Lopes |
| EPI_ISL_1381214 | Sentinelles Paris | National Reference Center for Viruses of Respiratory Infections, Institut Pasteur, Paris | Marion Barbet, Sylvie Behillil, Méline Bizard, Angela Brisebarre, Camille Capel, Louise Lefrançois, Etienne Simon-Lorière, Vincent Enouf, Maud Vanpeene, Sylvie van der Werf,Rousset Dominique |
| EPI_ISL_1381215, EPI_ISL_1381216, EPI_ISL_1381217 | Hospital | National Reference Center for Viruses of Respiratory Infections, Institut Pasteur, Paris | Marion Barbet, Sylvie Behillil, Méline Bizard, Angela Brisebarre, Camille Capel, Louise Lefrançois, Etienne Simon-Lorière, Vincent Enouf, Maud Vanpeene, Sylvie van der Werf,Rousset Dominique |
| EPI_ISL_1381218, EPI_ISL_1381219, | Labo Analyses Med | National Reference Center for Viruses of Respiratory | Marion Barbet, Sylvie Behillil, Méline Bizard, Angela Brisebarre, Camille Capel, Louise Lefrançois, Etienne Simon-Lorière, Vincent Enouf, Maud Vanpeene, |

| | | | |
|---|---|---|---|
| EPI_ISL_1381220, EPI_ISL_1381221, EPI_ISL_1381222, EPI_ISL_1381223 | | Infections, Institut Pasteur, Paris | Sylvie van der Werf,Rousset Dominique |
| EPI_ISL_1381304, EPI_ISL_1381305 | Omics Sciences Laboratory | Omics Sciences Laboratory | Derly Andrade Molina, Rubén Armas González, Gabriel Morey León, Darlyn Amaya, Katheryn Sacheri Viteri, Emily Sulay Saltos Montalvo, Paula Juliana Gavilanes Jarrín, Juan Carlos Fernández Cadena |
| EPI_ISL_568545, EPI_ISL_568546 | Laboratorio de Referencia Nacional de Virus Respiratorios, Instituto Nacional de Salud Peru | Laboratorio de Genómica Microbiana, Universidad Peruana Cayetano Heredia | Pablo Tsukayama, Alejandra Dávila-Barclay, Luis González, Pedro E. Romero, Brenda Ayzanoa, Janet Huancachoque, Pool Marcos, Maribel Huaringa, Camila Castillo-Vilcahuaman, Guillermo Salvatierra |
| EPI_ISL_671989 | UEES BioLab | Omics Sciences Laboratory | Derly Andrade Molina, Rubén Armas González, Gabriel Morey León, Darlyn Amaya, Katheryn Sacheri Viteri, Edith Lopez Montanero, Pedro Barberán, Fernando Espinoza Fuentes, Juan Carlos Fernández Cadena |
| EPI_ISL_717921, EPI_ISL_717922, EPI_ISL_717924, EPI_ISL_717925, EPI_ISL_717926, EPI_ISL_717927, EPI_ISL_717928, EPI_ISL_717929, EPI_ISL_717930, EPI_ISL_717931, EPI_ISL_717932, EPI_ISL_717933, EPI_ISL_717934, EPI_ISL_717935, EPI_ISL_717936, EPI_ISL_717937, EPI_ISL_717938, EPI_ISL_717939, EPI_ISL_717940, EPI_ISL_717941, EPI_ISL_717942, EPI_ISL_717943, EPI_ISL_717944, EPI_ISL_717945, EPI_ISL_717946, EPI_ISL_717947, EPI_ISL_717948, EPI_ISL_717949, EPI_ISL_717950, EPI_ISL_717951, EPI_ISL_717952, EPI_ISL_717953, EPI_ISL_717954, EPI_ISL_717955, EPI_ISL_717956, EPI_ISL_717957 | | | |
| see above | Laboratorio de Virologia Molecular / UFRJ | Bioinformatics Laboratory / LNCC | Carolina M Voloch, Ronaldo da Silva F Jr, Luiz G P de Almeida, Cynthia C Cardoso, Otavio Bustrolini, Alexandra L Gerber, Ana Paula de C Guimarães, Diana Mariani, Andréa Cony Cavalcanti, Claudia dos Santos Rodrigues, Terezinha M P P Castiñeira, Amilcar Tanuri, Ana Tereza R de Vasconcelos |
| EPI_ISL_755642 | Instituto Adolfo Lutz - Central | Instituto Adolfo Lutz, Interdiciplinary Procedures Center, Strategic Laboratory | Claudio Tavares Sacchi, Claudia Regina Gonçalves, Erica Valessa Ramos Gomes, Karoline Rodrigues Campos |
| EPI_ISL_755645 | Lab LOC - Itapecerica da Serra | Instituto Adolfo Lutz, Interdiciplinary Procedures Center, Strategic Laboratory | Claudio Tavares Sacchi, Claudia Regina Gonçalves, Erica Valessa Ramos Gomes, Karoline Rodrigues Campos |
| EPI_ISL_755649 | Instituto Adolfo Lutz - Regional de Santo Andre | Instituto Adolfo Lutz, Interdiciplinary Procedures Center, Strategic Laboratory | Claudio Tavares Sacchi, Claudia Regina Gonçalves, Erica Valessa Ramos Gomes, Karoline Rodrigues Campos |
| EPI_ISL_755651 | Instituto Adolfo Lutz - Central | Instituto Adolfo Lutz, Interdiciplinary Procedures Center, Strategic Laboratory | Claudio Tavares Sacchi, Claudia Regina Gonçalves, Erica Valessa Ramos Gomes, Karoline Rodrigues Campos |
| EPI_ISL_755652 | Lab LOC - Itapecerica da Serra | Instituto Adolfo Lutz, Interdiciplinary Procedures Center, Strategic Laboratory | Claudio Tavares Sacchi, Claudia Regina Gonçalves, Erica Valessa Ramos Gomes, Karoline Rodrigues Campos |
| EPI_ISL_755653 | Instituto Adolfo Lutz - Central | Instituto Adolfo Lutz, Interdiciplinary Procedures Center, Strategic Laboratory | Claudio Tavares Sacchi, Claudia Regina Gonçalves, Erica Valessa Ramos Gomes, Karoline Rodrigues Campos |
| EPI_ISL_756294 | Center for Biotechnology and Cell Therapy, São Rafael Hospital, Salvador, Brazil | Center for Biotechnology and Cell Therapy, São Rafael Hospital, Salvador, Brazil | Carolina Kymie Vasques Nonaka, Marília Miranda Franco, Tiago Gräf, Ana Verena Almeida Mendes, Renato Santana de Aguiar, Marta Giovanetti, Bruno Solano de Freitas Souza |
| EPI_ISL_770552, EPI_ISL_770553, EPI_ISL_770554, EPI_ISL_770556, EPI_ISL_770557, EPI_ISL_770559, EPI_ISL_770560, EPI_ISL_770561, EPI_ISL_770563, EPI_ISL_770564, EPI_ISL_770565, EPI_ISL_770566, EPI_ISL_770568, EPI_ISL_770570, EPI_ISL_770571, EPI_ISL_770578, EPI_ISL_770579, EPI_ISL_770580, EPI_ISL_770581, EPI_ISL_770583, EPI_ISL_770584, EPI_ISL_770587, EPI_ISL_770589, EPI_ISL_770591, EPI_ISL_770592, EPI_ISL_770593, EPI_ISL_770594, EPI_ISL_770595, EPI_ISL_770596, EPI_ISL_770598, EPI_ISL_770602, EPI_ISL_770603, EPI_ISL_770604, EPI_ISL_770605, EPI_ISL_770606, EPI_ISL_770607, EPI_ISL_770616, EPI_ISL_770617, EPI_ISL_770618, EPI_ISL_770619, EPI_ISL_770620, EPI_ISL_770621, EPI_ISL_770622, EPI_ISL_770624, EPI_ISL_770625, EPI_ISL_770628 | | | |
| see above | Laboratório de Microbiologia Molecular - Universidade FEEVALE | Bioinformatics Laboratory / LNCC | Felipe Benites, Fernando Rosado Spilki, Alana Witt Hansen, Juliane Deise Fleck, Juliana Schons, Meriane Demoliner, Ana Karolina Eisen Antunes, Fagner Henrique Heldt, Larissa Mallmann, Bruna Hermann, Ana Luiza Ziulkoski, Vyctoria Goes, Karoline Schallenberger, Matheus Nunes Weber, Paula Rodrigues de Almeida, Alessandra Pavan Lamarca da Silva, Ronaldo da Silva F Jr , Luiz G P de Almeida, Alexandra L Gerber , Ana Paula de C Guimarães,Ana Tereza R de Vasconcelos |
| EPI_ISL_778843 | Servicio Virosis Respiratorias-Departamento Virología-INEI | Instituto Nacional Enfermedades Infecciosas C.G.Malbran | Baumeister E., Avaro M., Benedetti E., Russo M., Dattero ME, Pontoriero A., Cisterna D., Molina V., Perandones C., Tuduri E., Lorenzo F., Poklepovich T., Campos J. |
| EPI_ISL_779155, EPI_ISL_779159 | Laboratório de Microbiologia Molecular - Universidade FEEVALE | Bioinformatics Laboratory / LNCC | Felipe Benites, Fernando Rosado Spilki, Alana Witt Hansen, Juliane Deise Fleck, Juliana Schons, Meriane Demoliner, Ana Karolina Eisen Antunes, Fagner Henrique Heldt, Larissa Mallmann, Bruna Hermann, Ana Luiza Ziulkoski, Vyctoria Goes, Karoline Schallenberger, Matheus Nunes Weber, Paula Rodrigues de Almeida, Alessandra Pavan Lamarca da Silva, Ronaldo da Silva F Jr , Luiz G P de Almeida, Alexandra L Gerber , Ana Paula de C Guimarães,Ana Tereza R de Vasconcelos |
| EPI_ISL_792522, EPI_ISL_792523, EPI_ISL_792524 | Laboratorio de Virología del Hospital de Niños Dr. Ricardo Gutierrez | Grupo de Genómica y Bioinformática del Instituto de Investigación de la Cadena Láctea CONICET-INTA on behalf of 'Proyecto Argentino Interinstitucional de genomica de SARS-CoV-2' (PAIS Consortium) | Amadio, AF, Eberhardt, MF; Irazoqui, M; Torres, C; Aulicino, P; König, G; Acevedo, ME; Alvarez Lopez, C; Alexay, S; Jacques, O; Mistchenko, AS, Goya, S; Nabaes Jodar, MS; Viegas, M. |
| EPI_ISL_792525 | Laboratorio del Hospital Interzonal General de Agudos Evita | Grupo de Genómica y Bioinformática del Instituto de Investigación de la Cadena Láctea CONICET-INTA on behalf of 'Proyecto Argentino Interinstitucional de genomica de SARS-CoV-2' (PAIS Consortium) | Amadio, AF, Eberhardt, MF; Irazoqui, M; Torres, C; Aulicino, P; König, G; Desimone, I; Luczac, E; Serrano, L; Grossi, O; Musto; Alexay, S; Goya, S; Nabaes Jodar, MS; Viegas, M. |
| EPI_ISL_792560 | Laboratorio de Ecologia de Doencas Transmissiveis na Amazonia, Instituto Leonidas e Maria Deane - Fiocruz Amazonia | Laboratorio de Ecologia de Doencas Transmissiveis na Amazonia, Instituto Leonidas e Maria Deane - Fiocruz Amazonia | Valdinete Nascimento, Victor Souza, André Corado, Fernanda Nascimento, George Silva, Ágatha Costa, Karina Pessoa, Debora Duarte, Luciana Gonçalves, Maria Júlia Brandão, Michele Jesus, Felipe Naveca on behalf of the Fiocruz COVID-19 Genomic Surveillance Network |
| EPI_ISL_792562, EPI_ISL_792634, EPI_ISL_792635 | Laboratório Central de Saúde Pública do Estado da Paraíba (LACEN-PB) | Laboratory of Respiratory Viruses and Measles, Oswaldo Cruz Institute, FIOCRUZ | Paola Resende, Luciana Appolinario, Fernando Motta, Anna Carolina Paixao, Ana Carolina Mendonca, João Felipe Bezerra, Romero Henrique Teixeira de Vasconcelos, Dalane Loudal Florentino Teixeira, Thiago Franco de Oliveira Carneiro, Marilda Siqueira on behalf of the Fiocruz COVID-19 Genomic Surveillance Network |
| EPI_ISL_792639, EPI_ISL_792642 | Laboratório Central de Saúde Pública do Estado de Alagoas (LACEN-AL) | Laboratory of Respiratory Viruses and Measles, Oswaldo Cruz Institute, FIOCRUZ | Paola Resende, Luciana Appolinario, Fernando Motta, Anna Carolina Paixao, Ana Carolina Mendonca, Anderson Brandao Leite, Marilda Siqueira on behalf of the Fiocruz COVID-19 Genomic Surveillance Network |
| EPI_ISL_792645, EPI_ISL_792646, EPI_ISL_792650, EPI_ISL_792651, EPI_ISL_792652 | Laboratório Central de Saúde Pública do Estado do Paraná (LACEN-PR) | Laboratory of Respiratory Viruses and Measles, Oswaldo Cruz Institute, FIOCRUZ | Paola Resende, Luciana Appolinario, Fernando Motta, Anna Carolina Paixao, Ana Carolina Mendonca, Maria do Carmo Debur, Irina Nastassja Riediger, Marilda Siqueira on behalf of the Fiocruz COVID-19 Genomic Surveillance Network |
| EPI_ISL_804814, EPI_ISL_804816, EPI_ISL_804819, EPI_ISL_804820, EPI_ISL_804821, EPI_ISL_804823, EPI_ISL_804824, EPI_ISL_804827, EPI_ISL_804828, EPI_ISL_804829 | DB Diagnosticos do Brasil | Laboratório de Parasitologia Médica - Instituto de Medicina Tropical - Universidade de São Paulo | Nuno Faria, Ingra Morales Claro, Darlan Candido, Lucas A. Moyses Franco, Pamela dos Santos Andrade, Thais de Moura Coletti, Camila A. Maia da Silva, Flavia Cristina Sales, Erika Regina Manuli, Renato A. Santana, Nelson Gaburo, Cecília da Cunha Camilo, Nelson Abrahim Fraiji, Myuki Alfaia Esashika Crispim, Maria do Perpétuo Socorro Sampaio Carvalho, Andrew Rambaut, Nick Loman, Oliver G. Pybus, Ester C. Sabino; DB; HEMOAM; CDL; CADDE Genomic Network. |
| EPI_ISL_811149 | Laboratorio de Ecologia de Doencas Transmissiveis na Amazonia, Instituto Leonidas e Maria Deane - Fiocruz Amazonia | Laboratorio de Ecologia de Doencas Transmissiveis na Amazonia, Instituto Leonidas e Maria Deane - Fiocruz Amazonia | Valdinete Nascimento, Victor Souza, André Corado, Fernanda Nascimento, George Silva, Ágatha Costa, Debora Duarte, Luciana Gonçalves, Matilde Mejía, Karina Pessoa, Maria Júlia Brandão, Michele Jesus, Felipe Naveca on behalf of the Fiocruz COVID-19 Genomic Surveillance Network |
| EPI_ISL_832010, EPI_ISL_832013 | Laboratório de Microbiologia Molecular - Universidade FEEVALE | Universidade Federal de Ciências da Saúde de Porto Alegre | Vinícius Bonetti Franceschi, Amanda de Menezes Mayer, Gabriel Dickin Caldana, Carla Andretta Moreira Neves, Patrícia Aline Gröhs Ferrareze, Gabriela Bettella Cybis, Ricardo Ariel Zimerman, Lívia Kmetzsch, Fernando Rosado Spilki, Claudia Elizabeth Thompson |
| EPI_ISL_833136, EPI_ISL_833137, EPI_ISL_833138, EPI_ISL_833139, EPI_ISL_833140 | Laboratorio de Ecologia de Doencas Transmissiveis na Amazonia, Instituto Leonidas e Maria Deane - Fiocruz Amazonia | Laboratorio de Ecologia de Doencas Transmissiveis na Amazonia, Instituto Leonidas e Maria Deane - Fiocruz Amazonia | Valdinete Nascimento, Victor Souza, André Corado, Fernanda Nascimento, George Silva, Ágatha Costa, Debora Duarte, Karina Pessoa, Matilde Mejía, Luciana Gonçalves, Maria Júlia Brandão, Michele Jesus, Felipe Naveca on behalf of the Fiocruz COVID-19 Genomic Surveillance Network |
| EPI_ISL_833158 | Instituto Adolfo Lutz - Regional de Santo Andre | Instituto Adolfo Lutz, Interdiciplinary Procedures Center, Strategic Laboratory | Claudio Tavares Sacchi, Claudia Regina Gonçalves, Erica Valessa Ramos Gomes, Karoline Rodrigues Campos |

| | | | |
|---|---|---|---|
| EPI_ISL_833161 | Instituto Adolfo Lutz - Central | Instituto Adolfo Lutz, Interdiciplinary Procedures Center, Strategic Laboratory | Claudio Tavares Sacchi, Claudia Regina Gonçalves, Erica Valessa Ramos Gomes, Karoline Rodrigues Campos |
| EPI_ISL_833163 | Instituto Adolfo Lutz - Regional de Marilia | Instituto Adolfo Lutz, Interdiciplinary Procedures Center, Strategic Laboratory | Claudio Tavares Sacchi, Claudia Regina Gonçalves, Erica Valessa Ramos Gomes, Karoline Rodrigues Campos |
| EPI_ISL_833167, EPI_ISL_833169, EPI_ISL_833170, EPI_ISL_833171, EPI_ISL_833172, EPI_ISL_833173, EPI_ISL_833174, EPI_ISL_833175, EPI_ISL_833176 | DB Diagnosticos do Brasil | Instituto Adolfo Lutz, Interdiciplinary Procedures Center, Strategic Laboratory | Claudio Tavares Sacchi, Claudia Regina Gonçalves, Erica Valessa Ramos Gomes, Karoline Rodrigues Campos |
| EPI_ISL_836143 | Hospital de Campanha COVID-19 de Mairipora | Instituto Adolfo Lutz, Interdiciplinary Procedures Center, Strategic Laboratory | Claudio Tavares Sacchi, Claudia Regina Gonçalves, Erica Valessa Ramos Gomes, Karoline Rodrigues Campos |
| EPI_ISL_836977 | Hospital Municiplal Dr. Jose de Carvalho Florence | Instituto Adolfo Lutz, Interdiciplinary Procedures Center, Strategic Laboratory | Claudio Tavares Sacchi, Claudia Regina Gonçalves, Erica Valessa Ramos Gomes, Karoline Rodrigues Campos |
| EPI_ISL_848557, EPI_ISL_848558, EPI_ISL_848559, EPI_ISL_848560, EPI_ISL_848606, EPI_ISL_848607, EPI_ISL_848608 | Evandro Chagas Institute | Evandro Chagas Institute | Santos, M.C.; Silva, A.M.; Junior, W.D.C.; Barbagelata, L.S.; Ferreira, J.A.; Sousa, E.M.A.; da Silva, P.S.; Pinheiro, K.C.; L.C.; Sousa Junior, E.C. |
| EPI_ISL_861668 | Centro de Triagem Covid19 | Instituto Adolfo Lutz, Interdiciplinary Procedures Center, Strategic Laboratory | Claudio Tavares Sacchi, Claudia Regina Gonçalves, Erica Valessa Ramos Gomes, Karoline Rodrigues Campos |
| EPI_ISL_861674, EPI_ISL_861675 | UPA Central de Caraguatatuba | Instituto Adolfo Lutz, Interdiciplinary Procedures Center, Strategic Laboratory | Claudio Tavares Sacchi, Claudia Regina Gonçalves, Erica Valessa Ramos Gomes, Karoline Rodrigues Campos |
| EPI_ISL_861676 | UPA Vila Santa Catarina | Instituto Adolfo Lutz, Interdiciplinary Procedures Center, Strategic Laboratory | Claudio Tavares Sacchi, Claudia Regina Gonçalves, Erica Valessa Ramos Gomes, Karoline Rodrigues Campos |
| EPI_ISL_861677 | Instituto Adolfo Lutz - Central | Instituto Adolfo Lutz, Interdiciplinary Procedures Center, Strategic Laboratory | Claudio Tavares Sacchi, Claudia Regina Gonçalves, Erica Valessa Ramos Gomes, Karoline Rodrigues Campos |
| EPI_ISL_861678 | Laboratorio Municipal de Guarulhos | Instituto Adolfo Lutz, Interdiciplinary Procedures Center, Strategic Laboratory | Claudio Tavares Sacchi, Claudia Regina Gonçalves, Erica Valessa Ramos Gomes, Karoline Rodrigues Campos |
| EPI_ISL_861679 | Instituto Adolfo Lutz - Regional de Taubate | Instituto Adolfo Lutz, Interdiciplinary Procedures Center, Strategic Laboratory | Claudio Tavares Sacchi, Claudia Regina Gonçalves, Erica Valessa Ramos Gomes, Karoline Rodrigues Campos |
| EPI_ISL_861683 | Complexo Hospitalar Padre Bento de Guarulhos | Instituto Adolfo Lutz, Interdiciplinary Procedures Center, Strategic Laboratory | Claudio Tavares Sacchi, Claudia Regina Gonçalves, Erica Valessa Ramos Gomes, Karoline Rodrigues Campos |
| EPI_ISL_861684, EPI_ISL_861685 | Hospital Pronto Socorro Itaquera | Instituto Adolfo Lutz, Interdiciplinary Procedures Center, Strategic Laboratory | Claudio Tavares Sacchi, Claudia Regina Gonçalves, Erica Valessa Ramos Gomes, Karoline Rodrigues Campos |
| EPI_ISL_861870, EPI_ISL_861872, EPI_ISL_861877, EPI_ISL_861878, EPI_ISL_861880, EPI_ISL_861882, EPI_ISL_861883, EPI_ISL_861887, EPI_ISL_861898, EPI_ISL_861899, EPI_ISL_861904 | | | |
| see above | LATE - Laboratório de Técnicas Especiais - Hospital Israelita Albert Einstein | LATE - Laboratório de Técnicas Especiais - Hospital Israelita Albert Einstein | Deyvid Amgarten, Fernanda de Mello Malta, Raquel Riyuzo, Ana Paula Moreira Salles, Pedro Henrique Sebe Rodrigues, João Renato Rebello Pinho |
| EPI_ISL_872191, EPI_ISL_872192 | Conjunto Hospitalar do Mandaqui de Sao Paulo | Instituto Adolfo Lutz, Interdiciplinary Procedures Center, Strategic Laboratory | Claudio Tavares Sacchi, Claudia Regina Gonçalves, Erica Valessa Ramos Gomes, Karoline Rodrigues Campos, Katia Correa de Oliveira Santos, Ana Lucia de Carvalho Avelino, Fabiana Cristina Pereira dos Santos |
| EPI_ISL_875688 | National Influenza Center - Instituto Adolfo Lutz | Instituto Adolfo Lutz, Interdiciplinary Procedures Center, Strategic Laboratory | Claudio Tavares Sacchi, Claudia Regina Gonçalves, Erica Valessa Ramos Gomes, Karoline Rodrigues Campos, Katia Correa de Oliveira Santos, Ana Lucia de Carvalho Avelino, Clovis Roberto Abe Constantino |
| EPI_ISL_875689 | Hospital do Servidor Publico | Instituto Adolfo Lutz, Interdiciplinary Procedures Center, Strategic Laboratory | Claudio Tavares Sacchi, Claudia Regina Gonçalves, Erica Valessa Ramos Gomes, Karoline Rodrigues Campos |
| EPI_ISL_882657 | LACEN do Estado do Piaui, Dr. Costa Alvarenga | Instituto Adolfo Lutz, Interdiciplinary Procedures Center, Strategic Laboratory | Claudio Tavares Sacchi, Claudia Regina Gonçalves, Erica Valessa Ramos Gomes, Karoline Rodrigues Campos |
| EPI_ISL_882668 | UMS de Juquitiba | Instituto Adolfo Lutz, Interdiciplinary Procedures Center, Strategic Laboratory | Claudio Tavares Sacchi, Claudia Regina Gonçalves, Erica Valessa Ramos Gomes, Karoline Rodrigues Campos |
| EPI_ISL_882669 | UPA Vila Santa Catarina | Instituto Adolfo Lutz, Interdiciplinary Procedures Center, Strategic Laboratory | Claudio Tavares Sacchi, Claudia Regina Gonçalves, Erica Valessa Ramos Gomes, Karoline Rodrigues Campos |
| EPI_ISL_882671, EPI_ISL_882673 | Hospital Municipal Dr. Guido Guida | Instituto Adolfo Lutz, Interdiciplinary Procedures Center, Strategic Laboratory | Claudio Tavares Sacchi, Claudia Regina Gonçalves, Erica Valessa Ramos Gomes, Karoline Rodrigues Campos |
| EPI_ISL_884002, EPI_ISL_890353 | Labo Analyses Med | National Reference Center for Viruses of Respiratory Infections, Institut Pasteur, Paris | Marion Barbet, Sylvie Behillil, Méline Bizard, Angela Brisebarre, Camille Capel, Etienne Simon-Lorière, Vincent Enouf, Maud Vanpeene, Sylvie van der Werf,Rousset Dominique |
| EPI_ISL_902736 | Dirección Departamental de Salud de Pública Leticia | Instituto Nacional de Salud- Dirección de Investigación en Salud Pública, Universidad de los Andes- Applied genomics research group, Vicerrectoria de Investigación y Creación, Universidad de los Andes- Systems and Computing Engineering Department | Katherine Laiton-Donato, Diego A. Álvarez-Díaz, Carlos Franco-Muñoz, Mauricio Pacheco-Montealegre, Maria T. Herrera-Sepúlveda, Diego Andrés Prada, Jorge Duitama, Laura Natalia Gonzalez, Jorge Ivan Diaz, Silvia Restrepo-Restrepo, Magdalena Wiesner, Martha Lucia Ospina Martinez, Marcela Mercado-Reyes |
| EPI_ISL_904120, EPI_ISL_904121 | LACEN - Laboratório Central de Saúde Pública do Pará | Evandro Chagas Institute | Santos, M.C.; Silva, A.M.; Junior, W.D.C.; Barbagelata, L.S.; Ferreira, J.A.; Sousa, E.M.A.; da Silva, P.S.; Pinheiro, K.C.; L.C.; Sousa Junior, E.C. |
| EPI_ISL_906068, EPI_ISL_906069 | Instituto Adolfo Lutz - Regional de Campinas | Instituto Adolfo Lutz, Interdiciplinary Procedures Center, Strategic Laboratory | Claudio Tavares Sacchi, Claudia Regina Gonçalves, Erica Valessa Ramos Gomes, Karoline Rodrigues Campos |
| EPI_ISL_906070 | UPA Dr. Akira Tada | Instituto Adolfo Lutz, Interdiciplinary Procedures Center, Strategic Laboratory | Claudio Tavares Sacchi, Claudia Regina Gonçalves, Erica Valessa Ramos Gomes, Karoline Rodrigues Campos |
| EPI_ISL_906071 | LACEN-PI DR. Costa Alvarenga | Instituto Adolfo Lutz, Interdiciplinary Procedures Center, Strategic Laboratory | Claudio Tavares Sacchi, Claudia Regina Gonçalves, Erica Valessa Ramos Gomes, Karoline Rodrigues Campos |
| EPI_ISL_906072 | UPA Dr. Akira Tada | Instituto Adolfo Lutz, Interdiciplinary Procedures Center, Strategic Laboratory | Claudio Tavares Sacchi, Claudia Regina Gonçalves, Erica Valessa Ramos Gomes, Karoline Rodrigues Campos |
| EPI_ISL_906075 | Hospital Geral de Vila Penteado Dr Jose Pangella Sao Paulo | Instituto Adolfo Lutz, Interdiciplinary Procedures Center, Strategic Laboratory | Claudio Tavares Sacchi, Claudia Regina Gonçalves, Erica Valessa Ramos Gomes, Karoline Rodrigues Campos |
| EPI_ISL_906076, EPI_ISL_906077 | Hospital Sao Luiz Sao Caetano | Instituto Adolfo Lutz, Interdiciplinary Procedures Center, Strategic Laboratory | Claudio Tavares Sacchi, Claudia Regina Gonçalves, Erica Valessa Ramos Gomes, Karoline Rodrigues Campos |
| EPI_ISL_906080, EPI_ISL_906081 | Hospital Beneficiencia Portuguesa | Instituto Adolfo Lutz, Interdiciplinary Procedures Center, Strategic Laboratory | Claudio Tavares Sacchi, Claudia Regina Gonçalves, Erica Valessa Ramos Gomes, Karoline Rodrigues Campos |

| | | | |
|---|---|---|---|
| EPI_ISL_906144, EPI_ISL_906145 | Laboratorio de Salud Publica de Amazonas | Instituto Nacional de Salud- Dirección de Investigación en Salud Pública, Universidad de los Andes- Applied genomics research group, Vicerrectoria de Investigación y Creación, Universidad de los Andes- Systems and Computing Engineering Department | Katherine Laiton-Donato, Diego A. Álvarez-Díaz, Carlos Franco-Muñoz, Mauricio Pacheco-Montealegre, Héctor Alejandro Ruiz-Moreno, Maria T. Herrera-Sepúlveda, Diego Andrés Prada, Jhonnatan Reales-González, Sheryll Corchuelo, Julian Naizaque, Gerardo Santamaría Jorge Duitama, Laura Natalia Gonzalez, Jorge Ivan Diaz, Silvia Restrepo-Restrepo, Magdalena Wiesner, Martha Lucia Ospina Martinez, Marcela Mercado-Reyes |
| EPI_ISL_918499, EPI_ISL_918500, EPI_ISL_918501, EPI_ISL_918502, EPI_ISL_918503, EPI_ISL_918504, EPI_ISL_918505, EPI_ISL_918506, EPI_ISL_918507, EPI_ISL_918508, EPI_ISL_918509, EPI_ISL_918510, EPI_ISL_918511 | | | |
| see above | LACEN - Laboratório Central de Saúde Pública do Amazonas | Evandro Chagas Institute | Santos, M.C.; Silva, A.M.; Junior, W.D.C.; Barbagelata, L.S.; Ferreira, J.A.; Sousa, E.M.A.; da Silva, P.S.; Pinheiro, K.C.; L.C.; Sousa Junior, E.C. |
| EPI_ISL_918514 | Evandro Chagas Institute | Evandro Chagas Institute | Santos, M.C.; Silva, A.M.; Junior, W.D.C.; Barbagelata, L.S.; Ferreira, J.A.; Sousa, E.M.A.; da Silva, P.S.; Pinheiro, K.C.; L.C.; Sousa Junior, E.C. |
| EPI_ISL_918516, EPI_ISL_918517 | LACEN - Laboratório Central de Saúde Pública do Para | Evandro Chagas Institute | Santos, M.C.; Silva, A.M.; Junior, W.D.C.; Barbagelata, L.S.; Ferreira, J.A.; Sousa, E.M.A.; da Silva, P.S.; Pinheiro, K.C.; L.C.; Sousa Junior, E.C. |
| EPI_ISL_918519, EPI_ISL_918520, EPI_ISL_918521 | Evandro Chagas Institute | Evandro Chagas Institute | Santos, M.C.; Silva, A.M.; Junior, W.D.C.; Barbagelata, L.S.; Ferreira, J.A.; Sousa, E.M.A.; da Silva, P.S.; Pinheiro, K.C.; L.C.; Sousa Junior, E.C. |
| EPI_ISL_918523, EPI_ISL_918524, EPI_ISL_918525, EPI_ISL_918526, EPI_ISL_918527, EPI_ISL_918528, EPI_ISL_918529, EPI_ISL_918530 | LACEN - Laboratório Central de Saúde Pública do Para | Evandro Chagas Institute | Santos, M.C.; Silva, A.M.; Junior, W.D.C.; Barbagelata, L.S.; Ferreira, J.A.; Sousa, E.M.A.; da Silva, P.S.; Pinheiro, K.C.; L.C.; Sousa Junior, E.C. |
| EPI_ISL_918534 | LACEN - Laboratório Central de Saúde Pública do Amazonas | Evandro Chagas Institute | Santos, M.C.; Silva, A.M.; Junior, W.D.C.; Barbagelata, L.S.; Ferreira, J.A.; Sousa, E.M.A.; da Silva, P.S.; Pinheiro, K.C.; L.C.; Sousa Junior, E.C. |
| EPI_ISL_918536, EPI_ISL_918537, EPI_ISL_918538, EPI_ISL_918540, EPI_ISL_918541, EPI_ISL_918542, EPI_ISL_918543, EPI_ISL_918544 | LACEN - Laboratório Central de Saúde Pública do Ceara | Evandro Chagas Institute | Santos, M.C.; Silva, A.M.; Junior, W.D.C.; Barbagelata, L.S.; Ferreira, J.A.; Sousa, E.M.A.; da Silva, P.S.; Pinheiro, K.C.; L.C.; Sousa Junior, E.C. |
| EPI_ISL_918545, EPI_ISL_918546, EPI_ISL_918547, EPI_ISL_918548, EPI_ISL_918549, EPI_ISL_918552 | LACEN - Laboratório Central de Saúde Pública do Para | Evandro Chagas Institute | Santos, M.C.; Silva, A.M.; Junior, W.D.C.; Barbagelata, L.S.; Ferreira, J.A.; Sousa, E.M.A.; da Silva, P.S.; Pinheiro, K.C.; L.C.; Sousa Junior, E.C. |
| EPI_ISL_918553, EPI_ISL_918555, EPI_ISL_918556, EPI_ISL_918557, EPI_ISL_918558, EPI_ISL_918559, EPI_ISL_918560, EPI_ISL_918561 | LACEN - Laboratório Central de Saúde Pública do Amapa | Evandro Chagas Institute | Santos, M.C.; Silva, A.M.; Junior, W.D.C.; Barbagelata, L.S.; Ferreira, J.A.; Sousa, E.M.A.; da Silva, P.S.; Pinheiro, K.C.; L.C.; Sousa Junior, E.C. |
| EPI_ISL_940613, EPI_ISL_940614, EPI_ISL_940615, EPI_ISL_940616, EPI_ISL_940617, EPI_ISL_940618 | LACEN-PI DR. Costa Alvarenga | Instituto Adolfo Lutz, Interdiciplinary Procedures Center, Strategic Laboratory | Claudio Tavares Sacchi, Claudia Regina Gonçalves, Erica Valessa Ramos Gomes, Karoline Rodrigues Campos |
| EPI_ISL_940619, EPI_ISL_940620, EPI_ISL_940621, EPI_ISL_940622, EPI_ISL_940623, EPI_ISL_940624, EPI_ISL_940625 | Hospital Sao Joaquim - Beneficiencia Portuguesa | Instituto Adolfo Lutz, Interdiciplinary Procedures Center, Strategic Laboratory | Claudio Tavares Sacchi, Claudia Regina Gonçalves, Erica Valessa Ramos Gomes, Karoline Rodrigues Campos |
| EPI_ISL_940626, EPI_ISL_940627 | Hospital Central Sao Caetano do Sul | Instituto Adolfo Lutz, Interdiciplinary Procedures Center, Strategic Laboratory | Claudio Tavares Sacchi, Claudia Regina Gonçalves, Erica Valessa Ramos Gomes, Karoline Rodrigues Campos |
| EPI_ISL_940628 | Unidade Mista de Iguape | Instituto Adolfo Lutz, Interdiciplinary Procedures Center, Strategic Laboratory | Claudio Tavares Sacchi, Claudia Regina Gonçalves, Erica Valessa Ramos Gomes, Karoline Rodrigues Campos |
| EPI_ISL_940630 | Hospital Geral de Sao Paulo | Instituto Adolfo Lutz, Interdiciplinary Procedures Center, Strategic Laboratory | Claudio Tavares Sacchi, Claudia Regina Gonçalves, Erica Valessa Ramos Gomes, Karoline Rodrigues Campos |
| EPI_ISL_943570 | Laboratorio de Referencia Nacional de Virus Respiratorio. Instituto Nacional de Salud Perú | Laboratorio de Referencia Nacional de Biotecnología y Biología Molecular. Instituto Nacional de Salud Perú | Carlos Padilla Rojas, Karolyn Vega Chozo, Luis Barcena, Priscila Lope Pari, Omar Caceres Rey, Marco Galarza Perez, Maribel Huaringa Nuñez, Johanna Balbuena Torrez, Henri Bailon Calderon, Nancy Rojas Serrano |
| EPI_ISL_943604, EPI_ISL_943607, EPI_ISL_943608, EPI_ISL_943610, EPI_ISL_943612 | Central Laboratory of Public Health of Rio Grande do Sul (Lacen-RS) | State Center for Health Surveillance of the Health Department of the State of Rio Grande do Sul (CEVS/SES-RS) | Aline Campos, Amanda da Silva, Anelise Schaurich, Claudia Dornelles, Cynthia Molina, Fernanda Godinho, Lara Crescente, Leticia Garay, Regina Barcellos, Richard Salvato, Tatiana Gregianini, Vagner Fonseca |
| EPI_ISL_943967, EPI_ISL_943968, EPI_ISL_943969, EPI_ISL_943970, EPI_ISL_943971, EPI_ISL_943972 | Hospital Geral de Sao Paulo | Instituto Adolfo Lutz, Interdiciplinary Procedures Center, Strategic Laboratory | Claudio Tavares Sacchi, Claudia Regina Gonçalves, Erica Valessa Ramos Gomes, Karoline Rodrigues Campos |
| EPI_ISL_943984, EPI_ISL_943986, EPI_ISL_943987 | LACEN do Estado de Tocantins | Instituto Adolfo Lutz, Interdiciplinary Procedures Center, Strategic Laboratory | Claudio Tavares Sacchi, Claudia Regina Gonçalves, Erica Valessa Ramos Gomes, Karoline Rodrigues Campos |
| EPI_ISL_943989, EPI_ISL_943990 | LACEN do Estado de Goias | Instituto Adolfo Lutz, Interdiciplinary Procedures Center, Strategic Laboratory | Claudio Tavares Sacchi, Claudia Regina Gonçalves, Erica Valessa Ramos Gomes, Karoline Rodrigues Campos |
| EPI_ISL_954058, EPI_ISL_954074, EPI_ISL_954075, EPI_ISL_954086, EPI_ISL_954089, EPI_ISL_954109 | Outre Mer | National Reference Center for Viruses of Respiratory Infections, Institut Pasteur, Paris | Marion Barbet, Sylvie Behillil, Méline Bizard, Angela Brisebarre, Camille Capel, Etienne Simon-Lorière, Vincent Enouf, Maud Vanpeene, Sylvie van der Werf,Rousset Dominique |
| EPI_ISL_956287, EPI_ISL_956289, EPI_ISL_956291, EPI_ISL_956292, EPI_ISL_956293, EPI_ISL_956295, EPI_ISL_956297 | Instituto Nacional de Salud- Dirección de Redes de Laboratorios de Salud Pública | Instituto Nacional de Salud- Dirección de Investigación en Salud Pública | Katherine Laiton-Donato, Diego A. Álvarez-Díaz, Carlos Franco-Muñoz, Mauricio Pacheco-Montealegre, Hector Alejandro Ruiz-Moreno, Maria T. Herrera-Sepúlveda, Diego Andrés Prada, Jhonnatan Reales-González, Sheryll Corchuelo, Julian Naizaque, Gerardo Santamaría, Magdalena Wiesner, Martha Lucia Ospina Martinez, Marcela Mercado-Reyes |
| EPI_ISL_956302 | LABORATORIO IMAT | Instituto Nacional de Salud- Dirección de Investigación en Salud Pública | Katherine Laiton-Donato, Diego A. Álvarez-Díaz, Carlos Franco-Muñoz, Mauricio Pacheco-Montealegre, Hector Alejandro Ruiz-Moreno, Maria T. Herrera-Sepúlveda, Diego Andrés Prada, Jhonnatan Reales-González, Sheryll Corchuelo, Julian Naizaque, Gerardo Santamaría, Magdalena Wiesner, Martha Lucia Ospina Martinez, Marcela Mercado-Reyes |
| EPI_ISL_962492, EPI_ISL_962493 | Omics Sciences Laboratory | Omics Sciences Laboratory | Derly Andrade Molina, Rubén Armas González, Gabriel Morey León, Darlyn Amaya, Katheryn Sacheri Viteri, Emily Sulay Saltos Montalvo, Paula Juliana Gavilanes Jarrín, Juan Carlos Fernández Cadena |
| EPI_ISL_977489 | UPA Dr. Akira Tada | Instituto Adolfo Lutz, Interdiciplinary Procedures Center, Strategic Laboratory | Claudio Tavares Sacchi, Claudia Regina Gonçalves, Erica Valessa Ramos Gomes, Karoline Rodrigues Campos |
| EPI_ISL_977490 | Hospital Municipal Guido Guida | Instituto Adolfo Lutz, Interdiciplinary Procedures Center, Strategic Laboratory | Claudio Tavares Sacchi, Claudia Regina Gonçalves, Erica Valessa Ramos Gomes, Karoline Rodrigues Campos |
| EPI_ISL_978522, EPI_ISL_978532 | Central Public Health Laboratory - LACEN -Bahia, Salvador, Brazil | Central Public Health Laboratory - LACEN -Bahia, Salvador, Brazil | Stephane Tosta, Luciana Oliveira, Vanessa Nardy,Patrícia Cajado,Marcela Gómez, Breno Dominguez, Jaqueline Gomes, Vagner Fonseca,Marta Giovanetti,Luiz Alcantara, Felicidade Pereira, Arabela Leal |
| EPI_ISL_981383, EPI_ISL_981385, EPI_ISL_981387 | IAL Regional de Bauru | Instituto Adolfo Lutz, Interdiciplinary Procedures Center, Strategic Laboratory | Claudio Tavares Sacchi, Claudia Regina Gonçalves, Erica Valessa Ramos Gomes, Karoline Rodrigues Campos |
| EPI_ISL_983863, EPI_ISL_983864, | Central Laboratory of Public Health of Rio Grande do Sul | State Center for Health Surveillance of the Health Department | Aline Campos, Cynthia Molina, Lara Crescente, Leticia Garay, Ludmila Fiorenzano Baethgen, Richard Salvato, Tatiana Gregianini |

| | | | |
|---|---|---|---|
| EPI_ISL_983865, EPI_ISL_983867 | (Lacen-RS) | of the State of Rio Grande do Sul (CEVS/SES-RS) | |
| EPI_ISL_984247, EPI_ISL_984248, EPI_ISL_984249, EPI_ISL_984250, EPI_ISL_984251, EPI_ISL_984252, EPI_ISL_984255, EPI_ISL_984256, EPI_ISL_984257, EPI_ISL_984258, EPI_ISL_984260, EPI_ISL_984261, EPI_ISL_984262 | | | |
| see above | IAL Regional de Marilia | Instituto Adolfo Lutz, Interdiciplinary Procedures Center, Strategic Laboratory | Claudio Tavares Sacchi, Claudia Regina Gonçalves, Erica Valessa Ramos Gomes, Karoline Rodrigues Campos |
| EPI_ISL_984619, EPI_ISL_984620, EPI_ISL_984621 | Central Laboratory of Public Health of Rio Grande do Sul (Lacen-RS) | State Center for Health Surveillance of the Health Department of the State of Rio Grande do Sul (CEVS/SES-RS) | Aline Campos, Cynthia Molina, Lara Crescente, Leticia Garay, Ludmila Fiorenzano Baethgen, Richard Salvato, Tatiana Gregianini |
| EPI_ISL_985303, EPI_ISL_985304, EPI_ISL_985305, EPI_ISL_985306, EPI_ISL_985307, EPI_ISL_985308, EPI_ISL_985309, EPI_ISL_985310, EPI_ISL_985311, EPI_ISL_985312, EPI_ISL_985313, EPI_ISL_985314, EPI_ISL_985315, EPI_ISL_985316, EPI_ISL_985317 | | | |
| see above | LACEN do Estado de Goias | Instituto Adolfo Lutz, Interdiciplinary Procedures Center, Strategic Laboratory | Claudio Tavares Sacchi, Claudia Regina Gonçalves, Erica Valessa Ramos Gomes, Karoline Rodrigues Campos |
| EPI_ISL_985318, EPI_ISL_985319 | LACEN de Santa Catarina | Instituto Adolfo Lutz, Interdiciplinary Procedures Center, Strategic Laboratory | Claudio Tavares Sacchi, Claudia Regina Gonçalves, Erica Valessa Ramos Gomes, Karoline Rodrigues Campos |
